# Full characterization of unresolved structural variation through long-read sequencing and optical genome mapping

**DOI:** 10.1101/2024.07.18.24310562

**Authors:** Griet De Clercq, Lies Vantomme, Barbara Dewaele, Bert Callewaert, Olivier Vanakker, Sandra Janssens, Bart Loeys, Mojca Strazisar, Wouter De Coster, Joris Robert Vermeesch, Annelies Dheedene, Björn Menten

## Abstract

Structural variants (SVs) are important contributors to human disease. Their characterization remains however difficult due to their size and association with repetitive regions. Long-read sequencing (LRS) and optical genome mapping (OGM) can aid as their molecules span multiple kilobases and capture SVs in full. In this study, we selected six individuals who presented with unresolved SVs. We applied LRS onto all individuals and OGM to a subset of three complex cases. LRS detected and fully resolved the interrogated SV in all samples. This enabled a precise molecular diagnosis in two individuals. Overall, LRS identified 100% of the junctions at single-basepair level, providing valuable insights into their formation mechanisms without need for additional data sources. Application of OGM added straighaorward variant phasing, aiding in the unravelment of complex rearrangements. These results highlight the potential of LRS and OGM as follow-up molecular tests for complete SV characterization. We show that they can assess clinically relevant structural variation at unprecedented resolution. Additionally, they detect (complex) cryptic rearrangements missed by conventional methods. This ultimately leads to an increased diagnostic yield, emphasizing their added benefit in a diagnostic sehng. To aid their rapid adoption, we provide detailed laboratory and bioinformatics workflows in this manuscript.

## Introduction

Structural variants (SVs) are genomic rearrangements of 50 base pairs (bp) or larger that are categorized into deletions, duplications, insertions, inversions, and translocations [1]. In addition, more complex genomic rearrangements (CGRs) exist, where multiple SV types are combined in a single event. Recent research indicates that one individual’s genome harbors 26,000 SVs on average, accounting for nearly 30 Mb of reorganized DNA [2]. Besides being part of normal variation between individuals, they also contribute to disorders such as cancer [3], autism [4], and (syndromic) intellectual disability [5].

Routine diagnostic techniques employed to detect structural variation consist of conventional karyotyping, fluorescence in situ hybridization (FISH), microarrays, and more recently (shallow) whole genome sequencing (WGS) [6]. These techniques are however unable to interrogate the full spectrum of structural variation and omen fail to pinpoint exact breakpoints. In a diagnostic sehng, precise identification and full characterization of SVs is however crucial as it allows for a conclusive clinical and molecular diagnosis. This can provide insight into the prognosis of a disease and its possible therapeutic management [7]. Short-read WGS has the potential to cater to these shortcomings, but due to the small read size inherent to this technology, the detection of SVs that are only a few kbs in size or reside within repeat regions is omen challenging [8]. Recently, long-read sequencing (LRS) and optical genome mapping (OGM) have been on the rise for accurate SV detection [9,10]. LRS is a sequencing technology that reads DNA and RNA at the single-base level, while OGM is a non-sequencing-based imaging technique that maps patterns of a fluorescent labelled DNA motif [11,12]. Reads generated by LRS typically measure between 10 to 100 kb, while OGM molecules start at 150 kb. Both techniques can however analyze fragments up to several Mbs in length [13]. Due to these long fragment lengths, SVs can be fully captured in unprecedented detail and resolution.

In this study we selected six individuals with a previously identified SV that remained unresolved through routine molecular techniques. In three cases no clear causal molecular diagnosis could be made through previous tests. We sequenced all individuals with nanopore LRS (Oxford Nanopore Technologies (ONT)), and additionally applied OGM (Bionano Genomics) to (highly) complex cases that benefit from further haplotype phasing. The aim of this study was to investigate the ability of LRS and OGM to fully characterize unresolved structural variation, their potential to identify missed cryptic rearrangements, and to evaluate their added value in a clinical sehng.

## Results

### Simple structural variants are identified at single base-pair level, enabling a new molecular diagnosis

Individual S1 presented with severe ID and dysmorphic facial features. Through karyotyping, an apparently balanced *de novo* reciprocal translocation between chromosome bands 9q21.2 and 10p15.2 was identified (table 1; sup. fig. S1a,b). As all other diagnostic tests came back normal, this translocation was highly suggestive as causal for the observed phenotype. However, exact breakpoint coordinates could not be further determined [14]. Application of LRS identified and characterized this variant at single-basepair level as chr9:g.pter_cen_82209673::chr10:g.11203945_pter (der9) and chr9:g.qter_82209674::chr10:g.11203946_cen_qter (der10) (fig. 1a,b; sup. table S3). Through sequence analysis, a 1 bp guanine insertion at the breakpoint of derivative 10 was found (sup. text S1). Additional confirmation by Sanger sequencing revealed the final variant to be a true event and 100% concordant with the LRS consensus. Microhomology analysis near the breakpoints revealed no microhomology stretches. The breakpoint on chromosome 9 falls within a LINE repeat, while on chromosome 10 the *CELF2* gene is disrupted (fig. 1a, sup. fig. S8a). Itai et al. (2021) [15] recently described deleterious variants in *CELF2* that lead to developmental and epileptic encephalopathy (OMIM #619561), and ID and autistic features through a loss-of-function mechanism. This led to a conclusive molecular diagnosis in this individual.

**Table 1.**
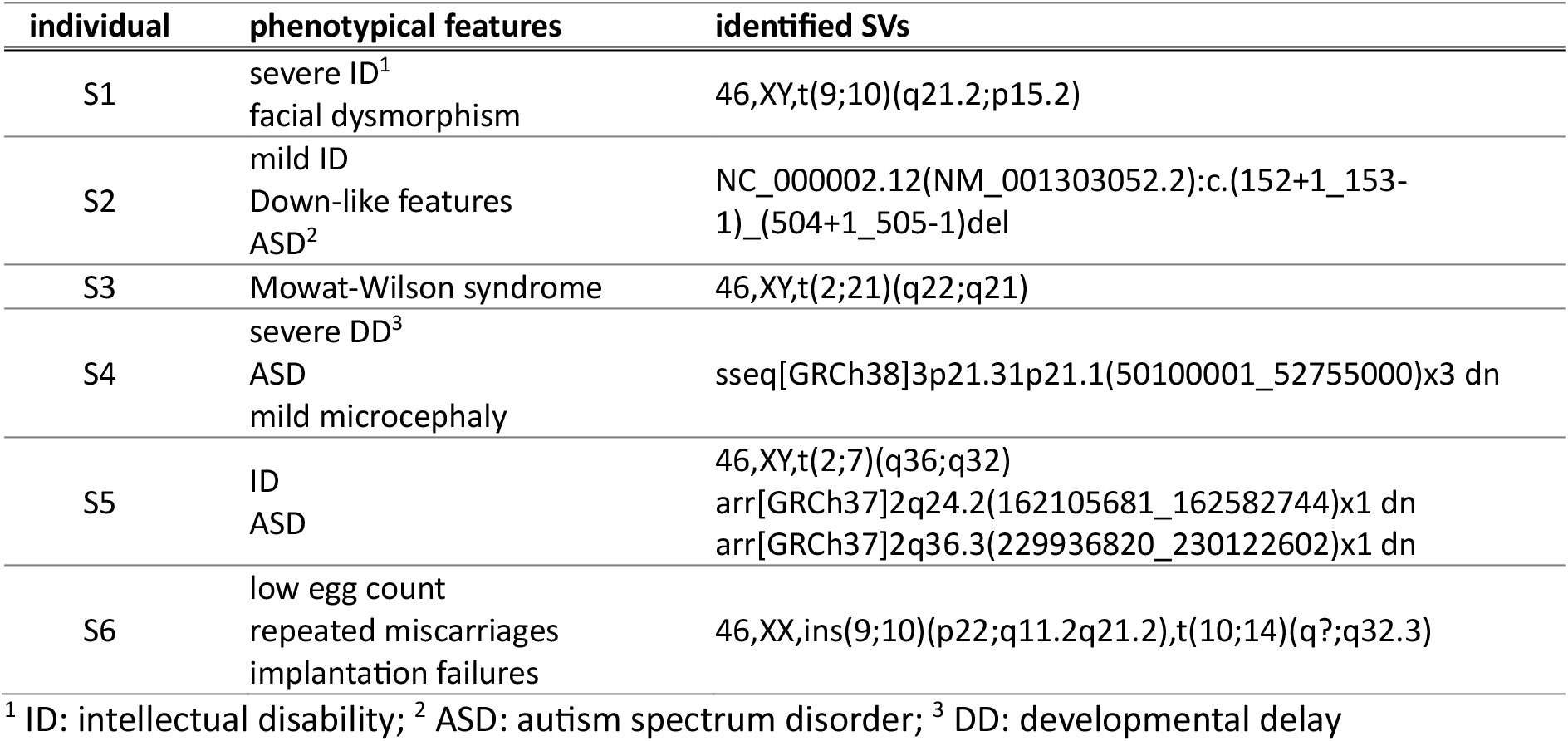
Phenotypical features of the six individuals (S1-6) included in this study, along with their unresolved SVs identified through standard clinical diagnostic testing. Individuals S1-4 presented with a simple structural variation event, while in individuals S5 and S6 a CGR was identified. Molecular diagnoses were made for all participants, except for individuals S1, S3, and S4.

Individual S2 manifested with mild ID, Down-like features, and autism spectrum disorder. Through whole exome sequencing a heterozygous *de novo* deletion was detected of exon 9 of the *MYT1L* gene (table 1). Variations in this gene are associated with autosomal dominant intellectual developmental disorder 39 (OMIM #616521). As this deletion results in an out-of-frame transcript, it was considered causal for the underlying phenotype. LRS was applied to further test its ability to characterize simple SVs. Variants were confined to the *MYT1L* region, which delineated the SV as a 4,574 bp deletion enclosing exon 9 (NC_000002.12:g.1939353-1943926del) (fig. 1c; sup. table S3; sup. fig. S4a,S8b). Microhomology analysis revealed a 1 bp microhomology at the junction breakpoint, and Sanger sequencing confirmed the junction sequence to be 100% concordant to the consensus sequence as determined by LRS (sup. text S2).

**Fig. 1.**
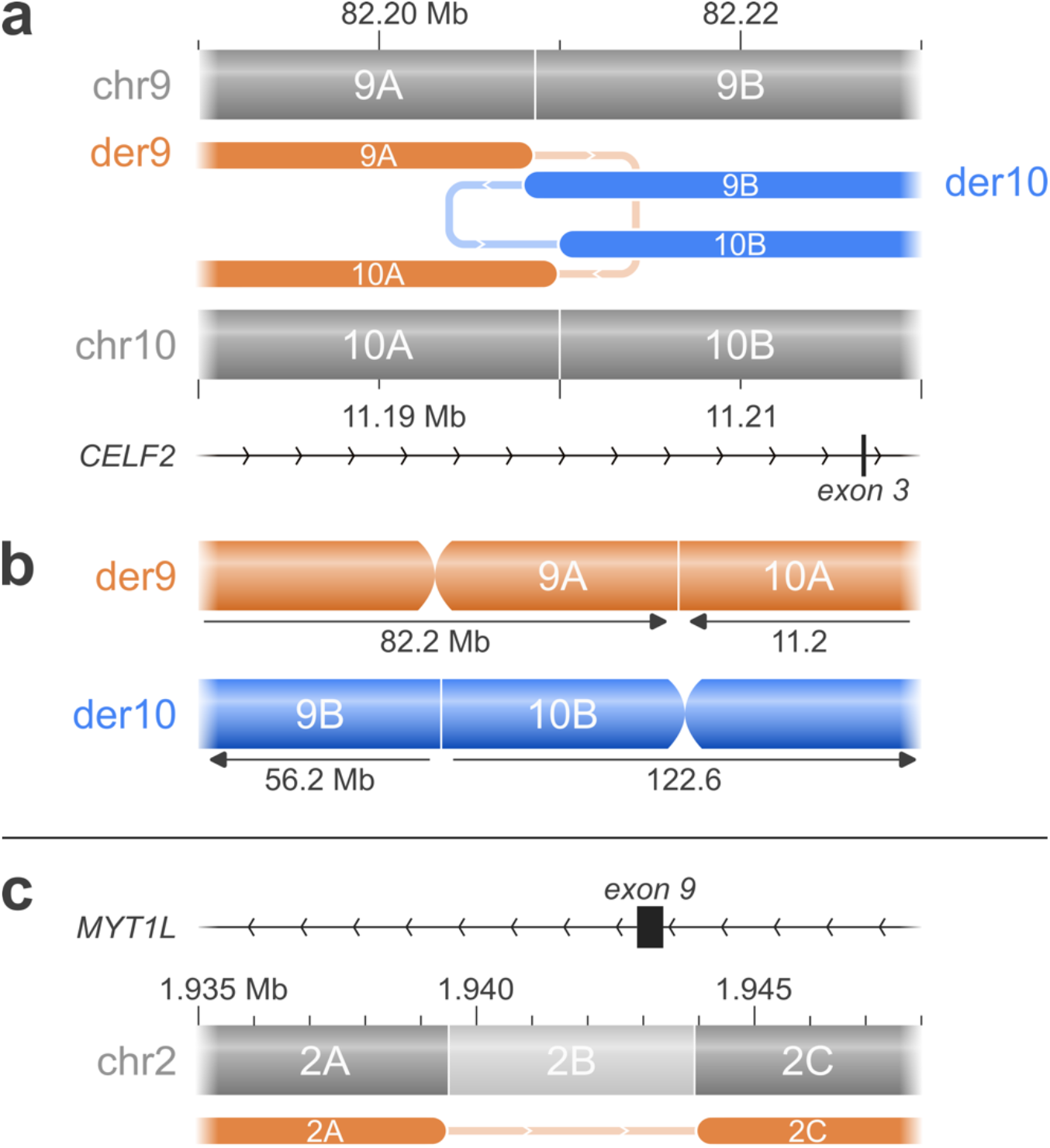
Simple SVs in individuals S1 and S2. **(a)** Exact breakpoint determina:on of the t(9;10) variant in individual S1 through LRS allowed for a novel molecular diagnosis through the disrup:on of the *CELF2* gene. **(b)** Local karyotype of the aberrant chromosomes of individual S1. **(c)** The exon 9 *MYT1L* dele:on in individual S2 was delineated through LRS at single base pair resolu:on as a 4,574 bp dele:on (fragment 2B) enclosing exon 9.

### Conventional techniques underestimate the incidence and complexity of structural variation

Individual S3 was referred to the clinic for multiple phenotypic aberrations corresponding to a clinical reciprocal translocation between chromosome bands 2q22 and 21q21 (table 1; sup. fig. S1c). Haploinsufficiency of *ZEB2*, located on chromosome band 2q22, can lead to Mowat-Wilson syndrome making this translocation likely causal for the observed phenotype. However, no exact breakpoints and thus no precise molecular diagnosis could be established through other diagnostic methods. LRS was applied and the variants filtered to retain translocations between the two long arms of chromosomes 2 and 21. This identified two distinct translocations in bands 2q22.3 and 21q21.2. These translocations however do not disrupt *ZEB2* and their breakpoints on chromosome 2 hit respectively the protein coding gene *GTDC1* and lncRNA gene *TEX41*. These genes are respectively lying up- and downstream of *ZEB2*, suggesting a more complex disruption of the region. Further manual inspection revealed other rearrangements which directly disrupt the *ZEB2* gene and involve chromosome 5 as well (sup. table S3,S8). Reconstruction of the separate fragments led to the discovery of a CGR consisting of 23 breakpoints on chromosomes 2, 5, and 21, affecting 1.4 Mb in total (fig. 2a,b; sup. fig. S7a). Due to its complexity, OGM was additionally applied to verify this reconstructed rearrangement. This confirmed the CGR, although some smaller fragments could not be detected through OGM analysis. Eight breakpoint junctions solely identified through LRS were therefore successfully confirmed through PCR (sup. table S5,S8). The final rearrangement was found to be relatively balanced with no large CNVs, except for a 25 kb deletion affecting exons 5 to 10 of the *ZEB2* gene (fig. 2a,b; sup. fig. S4g). This deletion was picked up by both LRS and OGM SV analysis (sup. fig. S9). LRS CNV analysis however called no copy number changing events in the affected regions (sup. table S2). Additionally, the vast majority of breakpoints are flanked by minor losses of genetic material, identified as deleted fragments in the CGR reconstruction (fig. 2a,b; sup. fig. S7a) and distinct drops in the LRS coverage data (sup. fig. S4b- n). The discovery of this previously concealed complex rearrangement affecting *ZEB2* led to a molecular diagnosis and the confirmation of the clinical diagnosis of Mowat-Wilson syndrome. Apart from *GTDC1* and *ZEB2*, no other protein coding genes were directly disrupted. Investigation for microhomology sequences around the LRS breakpoint junctions revealed 4 to 90 bp insertions in seven variants (sup. text S3). In four out of five junctions with insertions over 20 bp, the insertion sequences originated from regions within the CGR. Other variants presented with no to minimal microhomology (0 to 2 bp), except for one variant at the 2K-2M junction in derivative 5 where a microhomology stretch of 9 bp was detected.

**Fig. 2.**
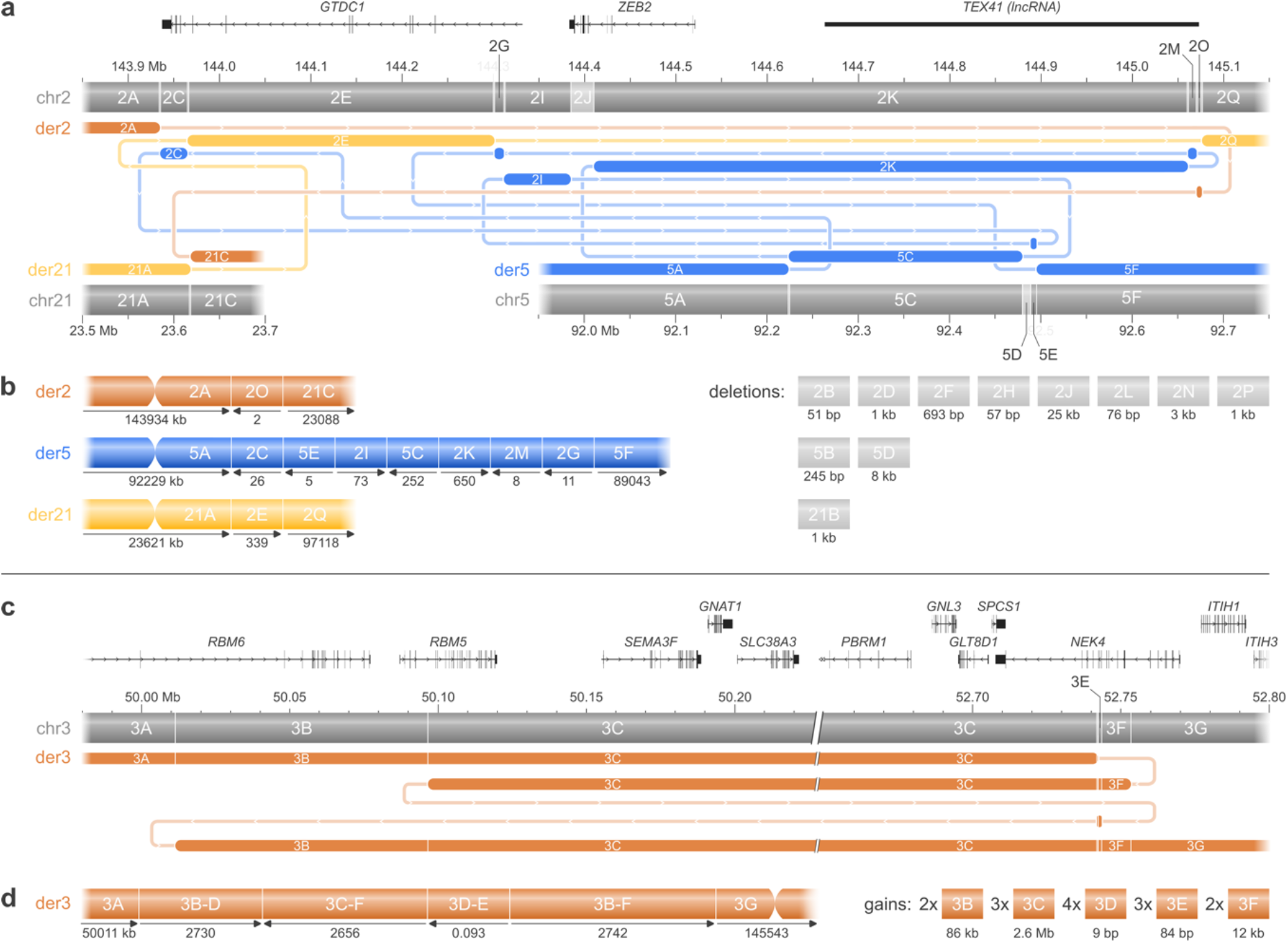
Simple SVs in individuals S3 and S4 turned out to be more complex variants. Apart from the lncRNA gene *TEX41*, only protein coding genes are shown in the figures. **(a)** Through LRS and OGM a seemingly simple t(2;21) variant in individual S3 was delineated as a complex rearrangement additionally involving chr5 and consisting of 23 breakpoints. The rearrangement shows previously missed insertions of chr2 into chr5 (blue fragments), along with the translocation between chr2 and chr21 (yellow and orange fragments). This delineation allowed for a molecular diagnosis through the disruption of *ZEB2* and confirmed the clinical diagnosis of Mowat-Wilson syndrome. **(b)** Local karyotype of the aberrant chromosomes of individual S3. **(c)** LRS revealed that a previously iden:fied triplica:on in individual S4 harbored a complex copy number changing rearrangement affecting 67 protein coding genes. **(d)** Local karyotype of the aberrant chromosome 3 of individual S4.

Individual S4 was diagnosed with severe developmental delay, autism spectrum disorder, microcephaly, and facial dysmorphism. Shallow WGS revealed a *de novo* 2.6 Mb triplication at chromosome bands 3p21.31p21.1, flanked upstream by a *de novo* 90 kb duplication (table 1; sup. fig. S2a). This rearrangement was highly suggestive as causal, due to its size and large number of affected genes and additional diagnostic tests revealing no other molecular aberrations. However, no immediate link could be established between this variant and the observed phenotype, and it was therefore diagnosed as a variant of unknown significance. SV analysis through LRS identified a complex copy-number changing rearrangement on chromosome 3 involving six breakpoints (sup. table S3,S9). This CGR consists of an 86.1 kb duplication, followed by an inverted 2.6 Mb triplication, a 9 bp quadruplication-inversion alongside an 84 bp triplication-inversion, and an inverted 11.6 kb duplication (fig. 2c,d; sup. fig. S5a-e). This rearrangement was fully verified through PCR (sup. table S5). CNV analysis through LRS was able to identify the 2.6 Mb triplication with identical genomic coordinates as previously reported through shallow WGS (sup. table S2). The flanking 86.1 kb duplication was however not called. Microhomology analysis detected no microhomology at the 3D-3F junction, a templated 30 bp inserted sequence at the 3C-3E junction, and a 1 bp microhomology at the 3D-3B junction (sup. text S4). The rearrangement covers 67 protein coding genes of which 17 have an associated OMIM phenotype (sup. table S3,S4). Furthermore, it has breakpoints directly disrupting non-coding regions of *RBM6*, *RBM5*, and *NEK4,* along with a breakpoint falling within exon 13 of *NEK4*. None of these impacted genes can however be unambiguously linked to the observed phenotype. OGM was not further applied as all variants could be easily phased and reconstructed through LRS alone.

### Unresolved complex rearrangements are fully delineated, providing insight into their formation mechanisms

The phenotype for individual S5 consisted of ID and autism spectrum disorder. Chromosome analysis revealed a balanced *de novo* reciprocal translocation between chromosome bands 2q36 and 7q32 (table 1; sup. fig. S1d). In addition, two heterozygous *de novo* CNVs were detected on chromosome 2 through microarray analysis, consisting of a 477 kb deletion in 2q24.2 and a 186 kb deletion in 2q36.3 (sup. fig. S2b,c). The 477 kb deletion in the 2q24.2 region encompasses the *TBR1* gene, described in intellectual developmental disorder with autism and speech delay (OMIM #606053). This variant was therefore considered causal for the associated phenotype. Furthermore, the translocation and 186 kb counseling of a family member, an attempt was made to delineate this region through further research efforts, which however proved fruitless. LRS was therefore applied. Variants were filtered to retain events lying within the long arms of chromosome 2 and 7, manually scanned at the previously defined regions of interest, and further expanded upon finding cryptic variants. This approach generated multiple variants of interest at chromosome band 2q24.2, consisting of one 190 kb inversion, one 839 kb deletion, and several translocations to the short arm of chromosome 7 (sup. table S10). The translocations were identified as a novel 347 kb insertional inversion of chromosome 2 into chromosome 7. One of its breakpoints on chromosome 2 coincides with the 839 kb deletion, and these two variants were therefore considered to be part of the same event. Here, a 347 kb part of the 839 kb deletion was inserted back into chromosome 7, resulting in a 492 kb loss of material of chromosome 2. This is in line with the previous finding of a 477 kb deletion at location 2q24.2. The 190 kb inversion lies 76 kb upstream of this complex variant. Phasing analysis through LRS could not be achieved due to the absence of informative single nucleotide variants, but additional application of OGM confirmed the CGR and identified all variants to be part of the same haplotype (event 1 in fig. 3a; sup. fig. S10a). At location 2q36.3, a 222 kb deletion and several translocations to chromosome band 7q32.2 were found through LRS. The translocations were deemed to be part of the same event that covers the t(2;7)(q36.3;q32) variant identified through karyotyping (event 2 in fig. 3a). The 222 kb deletion present in the same chromosome band 2q36.3 coincides with the previously detected 186 kb deletion at the same location. It was however considered separate from the translocation as it localizes nearly 2 Mb further downstream and phasing information could not be retrieved through LRS nor through OGM (event 3 in fig. 3a). OGM confirmed the variants in event 2 and 3 (fig 3a). Delineation of the SVs previously identified within this sample thus characterized them as three events, with the inversion and inverted insertion at region 2q24.2 as novel finds (fig. 3a,b; sup. fig. S7b). The two deletions at chromosome bands 2q24.2 and 2q36.3 were detected through copy number calling as well (sup. table S2). Microhomology analysis revealed only limited microhomology (0 to 3 bp) at the LRS breakpoint junctions for most variants, except for the 2F-7C junction where a 7 bp microhomology stretch was detected (sup. text S5). At the 2C-2F junction a thymine insertion was identified. The detected SVs disturb four genes (*SLC4A10*, *COL4A3*, *PID1, CPA2*) directly through a breakpoint and two other protein coding genes (*PSMD14*, *TRB1*) through a complete loss of genetic material (sup. table S3).

**Fig. 3.**
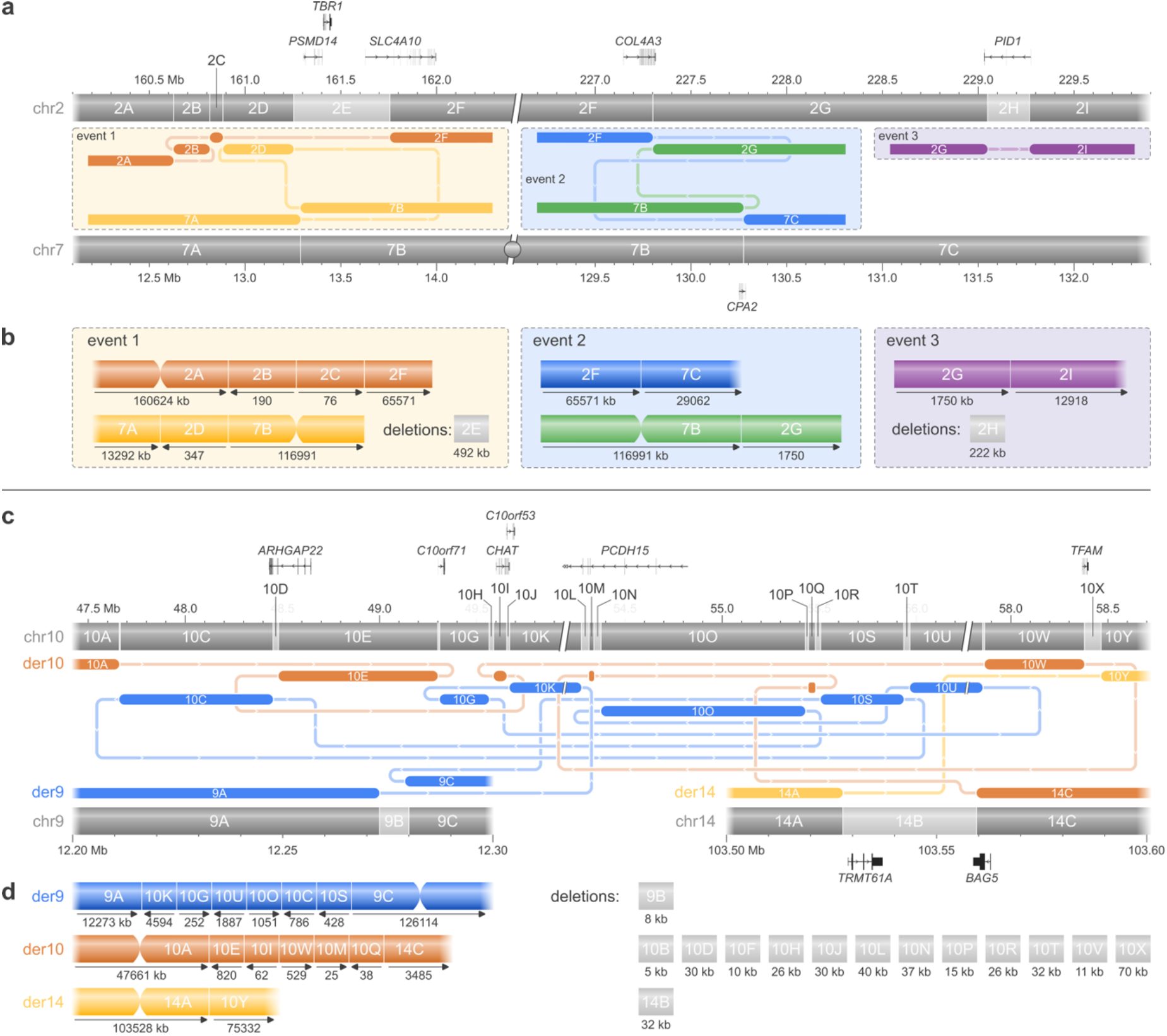
Delinea:on of previously iden:fied CGRs in individuals S5 and S6. Only genes directly affected by a breakpoint or deletion are shown in the figures. **(a)** In individual S5 conven:onal techniques detected two dele:ons at respec:vely chromosome bands 2q24.2 and 2q36.3, along with a t(2;7)(q36;q32) reciprocal translocation. The 2q36.3 dele:on and the transloca:on were thought to be connected to each other and arisen through one single complex event. Application of LRS confirmed all previously detected variants, and iden:fied an addi:onal ins(2;7) variant connected to the 2q24.2 dele:on and an upstream cryp:c inversion (event 1). The t(2;7)(q36;q32) variant and the 2q36.3 deletion were found to be localizing nearly 2 Mb from each other and were thus considered as two dis:nct events (respec:vely events 2 and 3). **(b)** Local karyotypes of the three events identified in individual S5. **(c)** Individual S6 presented with a complex karyotype consisting of an ins(9;10) and a t(10;14) variant. LRS and OGM further delineated this CGR up un:l single base pair resolu:on and iden:fied a much more intricate rearrangement involving 28 breakpoints on chr9, 10, and 14. It features 10 deletions larger than 15 kb and disrupts in total 10.8 Mb of genetic material, causal for the observed fertility problems. **(d)** Local karyotype of the aberrant chromosomes of individual S6.

Individual S6 was referred to the clinic amer repeated miscarriages and recurrent implantation failure. Chromosome analysis showed an aberrant complex karyotype consisting of a translocation between chromosome bands 10q21.2 and 14q32.3, and an insertion of chromosome region 10q11.2 to 10q21.2 into 9p22 (table 1; sup. fig. S1e-g). These variants were considered causal for the observed phenotype as CGRs can lead to unbalanced gametes. Additional shallow WGS identified five deletions on chromosome 10 ranging from 30 to 120 kb in length and one 30 kb deletion on chromosome 14 (sup. fig. S3). LRS was applied to delineate the full situation. SV analysis exposed additional cryptic SVs (sup. table S11), which were reconstructed back into a 10.8 Mb CGR consisting of 28 breakpoints involving chromosomes 9,10, and 14 (fig. 3c,d; sup. table S3; sup. fig. S7c). Through this manual reconstruction, 14 deleted fragments were identified, of which 10 larger than 15 kb (sup. fig. S6). Six of these coincide with the six previously detected deletions. CNV analysis through LRS was however not able to identify any copy number changes in the regions of interest (sup. table S2). Breakpoint junction analysis revealed no inserted sequences between fragments and limited microhomology (1 to 5 bp) in most junctions (sup. text S6). Seven protein coding genes are directly disturbed by a breakpoint (*ARHGAP22, C10orf71, CHAT, C10orf53, PCDH115, TFAM, BAG5*), while an eighth gene (*TRMT61A*) falls entirely within a deleted fragment on chromosome 14 (sup. table S3). OGM was additionally applied to verify the reconstruction of this highly complex rearrangement. This fully confirmed the LRS findings, yet some smaller fragments could not be delineated. Four junctions seen solely through LRS analysis were subsequently successfully confirmed by PCR (sup. table S5).

### Long-read sequencing identifies 100% of breakpoint junctions

The majority of SV junctions in this study were detected through use of the Sniffles2 LRS SV caller (87%, 62/71, n=6), with a mean deviation to the actual breakpoint location of 3 ± 5 bp (n=76) (sup. table S6- 12). The junctions that were not detected through Sniffes2 could still be retrieved from the LRS data through manual curation (sup. fig. S10b), resulting in a 100% pick-up rate through LRS (71/71, n=6). OGM on the other hand, was able to retrieve 67% (41/61) of the junctions directly through automated data analysis, and 74% (45/61) amer manual curation (n=3). The average deviation from the actual breakpoint coordinate through OGM was 5.0 ± 5.5 kb (n=66).

## Discussion

In this study we used LRS as the main method for SV characterization and utilized OGM as a subsequent verification method for more complex cases. Through this, we show their potential as a follow-up diagnostic test to delineate unresolved SVs. This is of importance in a diagnostic sehng as it enables a molecular diagnosis, influences future disease management, and can expose undiscovered disease-associated genes. Through the application of LRS we successfully characterized multiple clinically relevant rearrangements in six individuals up until single basepair level. This led to a conclusive molecular diagnosis in two individuals, which was previously unattainable through standard diagnostic techniques alone. A third individual remained without a precise diagnosis, although application of LRS may give new insights into the underlying disease mechanism. The rearrangement disturbs exon 13 of *NEK4* and several introns of *RBM6*, *RBM6*, and *NEK4*. None of these genes can currently be linked to intellectual disability yet may constitute novel candidate genes. Alternatively, the newly discovered inverted triplication could explain an alternative pathogenic mechanism through disturbed expression patterns of its impacted genes or regulatory regions [16]. Future functional work to investigate this is however outside the scope of this study.

The ability of LRS to characterize SVs at single base pair resolution and reveal their exact sequence provides insight into their formation origins without the need for additional molecular methods. Probands S1 and S2 both manifested with a simple SV. Their respective variants lack considerable microhomology at the breakpoints and are lying outside repeat elements, indicating non-homologous end joining (NHEJ) as the most likely formation mechanism. Slightly more complex cases were seen in individuals S4 and S5. The CGR in participant S4 consists of multiple copy-number changes with several junctions lying within repeat elements, hinting towards a replication defect based on the fork stalling and template switching/microhomology-mediated break-induced replication pathway. Indeed, extensive reports exist within literature describing similar DUP-TRIP/INV-DUP events driven by the 3a,b) can be explained through the same formation mechanism. Non-allelic homologous recombination is typically observed in recurrent SVs yet can also drive non-recurrent events between smaller repetitive sequences [19]. Most fused junctions identified in this case indeed lie within repeat elements of the same family. Event 3 however does not fit this mechanism as only one junction resides within a repetitive region. Microhomology-mediated end joining (MMEJ) could then serve as an alternative explanation, although here again, not all identified junctions adhere to its general characteristics. This may point to two distinct processes driving the formation of the observed variants in this individual instead of one.

Individuals S3 and S6 presented with highly intricate rearrangements which seem to originate from separate mechanisms considering their differing characteristics. The CGR in proband S3 has templated inserted sequences between breakpoints, microhomologies up to 7 bp, and no large CNVs apart for one 25 kb deletion. We here propose chromothripsis through MMEJ as a formation mechanism. MMEJ can act as a repair mechanism for double strand breaks such as those generated in mass during chromothripsis yet is less preferred over the more utilized NHEJ pathway [20]. Most commonly it only produces deletions and features loss of nucleotides at the breakpoints to reveal microhomology necessary for mediating its mechanism. Indeed, flanking deletions of 1 kb or smaller can be observed between junctions (fig. 2b). Case S6 has limited microhomologies at variant junctions, no insertions between breakpoints, and several deletions larger than 15 kb. These observations are consistent with chromothripsis through NHEJ [21].

The complex rearrangements detected in S3-S6 were followed up with OGM to confirm their reconstruction achieved through LRS and to aid with haplotype phasing. Both techniques act complementary to each other and enhance their respective qualities. LRS reads offer single base pair resolution, yet only typically capture one SV or one SV junction per read. On the other hand, OGM molecules omen span multiple variants in one read and as such, can naturally phase variants together.

Due to this difference in technology, LRS and OGM detect rearrangements in a significantly different manner. LRS unveils breakpoints and breaks the affected region up in fragments (sup. fig. S9a) that have to be manually puzzled back together, making CGR reconstruction a feasible yet laborious task. However, OGM detects variants in an aberrant haplotype approach (sup. fig. S9b,c). OGM thus allows for easy phasing and reconstruction of variants, while LRS adds single base pair resolution. As demonstrated in this study, their parallel application is therefore especially powerful for CGR characterization and allows for the easy unraveling of highly complex rearrangements. Note however that OGM glosses over intricate details within the more complex CGRs due to its resolution of 5 kb. While OGM detects 74% of the junctions within its applied complex cases, this was 100% with LRS alone. Application of each technology as a stand-alone method may thus manifest as a trade-off between detailed resolution versus straighaorward variant reconstruction in complex situations.

Historically, CGRs have been considered as rare constitutional events, yet recent studies indicate they are more common than previously anticipated [18]. This is reflected in our study as we find highly complex rearrangements in four out of six included individuals. Especially of interest is case S6. This individual is phenotypically normal apart from reproductive issues, despite the presence of a rearrangement consisting of 28 breakpoints affecting eight protein coding genes. CGRs are indeed occasionally encountered in individuals with fertility problems who are otherwise healthy [22], and might be more common than anticipated around reciprocal translocations [23]. A similar observation was recently made by Eisfeldt *et al*. (2021) [24], who unraveled a CGR involving 137 breakpoints through OGM and LRS, among other technologies. Their application may thus lead to more informed decisions in sehngs where unidentified SVs can result in undesirable outcomes, such as in fertility assistance. Preimplantation embryos have a tendency to form micronuclei, which in turn are susceptive to the formation of complex rearrangements through chromoanagenesis [25]. Current clinical technologies however underestimate both the incidence and complexity of CGRs and may thus fail to identify these events [18], as demonstrated here as well. LRS and OGM can aid in more accurately assessing the implantation success of these embryos and provide more substantiated reproductive decisions. Especially LRS could prove beneficial in this sehng, considering its ability to assess the formation origins of several chromoanagenesis cases in this study without the need for additional data sources.

In this manuscript, we furthermore include detailed laboratory and bioinformatics protocols that can be utilized as an initial framework to implement LRS and OGM in a clinical sehng. These can be applied to identify genome-wide structural variation in LRS and OGM data. We show our protocols can be employed in heterogeneous conditions, as they were successfully applied on divergent coverages, instruments, and input materials. However, the differing experimental states mean we cannot mutually compare the included samples to each other considering their overall quality statistics and variant calls. In addition, separate CNV detection through LRS data failed to pick up a considerable number of verified variants. This inability to detect CNVs through a separate analysis on the same LRS dataset highlights an important hiatus in current SV detection tools. Due to the lack of a dedicated LRS CNV calling algorithm, we used a CNV caller originally designed for short reads. This could fail to detect signals inherent to LRS alone, resulting in missed variants. We therefore recommend cautious result interpretation of these tools and propose a prior benchmarking on a verified set of CNVs to assess their performance before employment in a diagnostic sehng.

A limitation of this study is its small cohort size. While LRS and OGM were able to assess the clinically relevant SV in all cases, the limitations of the isolated use of these techniques could still interfere with successfully making a molecular diagnosis. Studies leveraging larger sample sizes however show similar promising results for the application of both LRS and OGM in a clinical sehng [9],[10]. Furthermore, the current study heavily benefits from a priori knowledge obtained through previous tests. It has to be taken into account that if LRS or OGM were to be applied blind, then stringent filtering needs to be in place to identify the causal variant out of the myriad of SVs present within the human genome. A crucial aspect of implementing a diagnostic test is namely its ability to distinguish between benign and within the Bionano pipeline, yet its impact remains to be evaluated as studies applying this approach in a blind diagnostic sehng are lacking. Especially in the context of LRS, these studies would need to rely on internal and external databases of common SVs present within the population to filter out irrelevant variation. Few such databases exist [26] and ideally need to be extended with data from emerging technologies that excel at SV characterization. We therefore envision that wide-scale employment of these technologies in a diagnostic sehng would further facilitate the sharing of these events and thus their filtering and interpretation.

In conclusion, we show that nanopore LRS coupled with Bionano OGM offers several benefits compared to the currently employed diagnostic techniques used for SV identification. Not only are they able to detect previously identified SVs, they also allow for a complete characterization of currently unresolved SVs up to base-level resolution and the detection of cryptic rearrangements missed by conventional genetic tests. Furthermore, LRS provides valuable insights in the formation mechanisms behind an SV and works in synergy with OGM to easily delineate CGRs, a currently underestimated variant class. Most importantly, the ability of LRS to pinpoint exact breakpoint locations enables molecular diagnoses, resulting in a modest higher diagnostic yield.

## Materials and Methods

### Study design

Six individuals (S1-6) were selected for whom an SV was identified through standard clinical diagnostic methods, yet exact breakpoint coordinates could not be established. The structural variation events were determined to be causal for the underlying phenotype, except for individuals S1, S3, and S4 where no conclusive molecular diagnosis could be made. Standard testing included a selection of G- banded chromosome analysis, FISH, whole exome sequencing, shallow WGS, qPCR, and microarray analysis. For four individuals a simple SV event consisting of maximum two breakends was identified, including two translocations (S1,3), a deletion (S2), and a triplication (S4). For two individuals (S5,6) a rearrangement consisting of three or more breakends, referred to as a CGR, was detected. All individuals presented with ID or DD, except for S6 who came to the clinic due to reduced fertility. All SVs occurred *de novo*, except for S6 for which the *de novo* status could not be further evaluated through segregation analysis. All clinical diagnoses and SVs identified through standard clinical testing can be consulted in table 1. More detailed information on the observed phenotypes and specific conducted genetic tests per individual can be found in supplementary methods.

### Long-read whole genome sequencing

#### Sample preparation and sequencing

High molecular weight (HMW) DNA was extracted from blood or lymphoblastoid cell lines in an automated manner using respectively the Genomic DNA Large Volume Whole Blood Kit (1.200µl; RBC Bioscience) and the Genomic DNA Cultured Cells Kit (RBC Bioscience). Quantification was done with a Qubit fluorometer (Thermo Fisher) and the dsDNA HS Assay kit (Thermo Fisher). DNA was subsequently sheared to 20-30 kb lengths by placing g-TUBES (Covaris) in a centrifuge at 3200 rpm for four minutes, or size selected to enrich for sizes of 10 kb or greater using the Short Read Eliminator XS kit (PacBio) (sup. table S1). Approximately 7 µg of processed DNA was used as input for the EXP-

BND104, SQK-LSK109, or the SQK-LSK114 library preparation kits (ONT) following the manufacturer’s protocol with minor adjustments. Briefly, modifications consisted of longer incubation times and elution in higher volumes to maximize the retention of HMW DNA fragments. Libraries were then sequenced on a MinION or PromethION 24 device (ONT) for 80 hours, with flushing and reloading amer 24 and 48 hours to boost the final yield. Flow cells were of type R9.4.1 or R10.4.1 depending on the used library preparation kit. Loading quantities in fmol were calculated based on size profiles obtained from a TapeStation 4150 device (Agilent) using the Genomic DNA ScreenTape kit (Agilent). Additional library preparation and sequencing was done for CGR samples where the full rearrangement could not be reconstructed and the coverage was below 15x (coverage cut-off determined through internal data, paper in preparation). More detailed information on the processing and quality statistics of each sample can be found in supplementary table S1.

#### Sequencing analysis and structural variant detection

Raw sequencing data were basecalled with Guppy v6.3.7 (ONT) using the super accuracy model without q-score filtering. Demultiplexing was done using the Guppy v6.3.7 barcoder (ONT). Reads were aligned to the hg38 reference genome with minimap2 v2.24 [27], and split libraries were merged into one alignment file using samtools v1.15 [28]. Coverage depth was calculated using Mosdepth v0.3.3 [29]. Other quality control metrics were checked with NanoPlot v1.40.0 [30] and PycoQC v2.5.2 [31]. SVs were called using Sniffles2 v2.0.7 [32] with a read support parameter of one, and further processed to only retain variants with a read support of three or higher and a length of at least 50 bp. Resulting alignment and variant files were filtered to relevant genomic regions based on previous diagnostic tests using respectively samtools v1.15 [28] and vcmools v0.1.16 [33]. Both were then scanned manually using IGV v2.13.2 [34] for SVs of interest, with special attention paid near identified breakpoints to allow for potentially undetected CGRs. If no variants of interest were found or a CGR was suspected, the filter criteria were loosened to allow for SVs at lower read supports and analysis was repeated. The total number of SVs in a sample was calculated as all variants outputted by Sniffes2 with a length of at least 50 bp and with an aberrant read support of three or higher. Detailed commands regarding this workflow can be found in supplementary methods.

#### Copy number variant detection

Large CNVs (≥ 15 kb) were called from the LRS data using WisecondorX v1.2.5 [35]. A custom reference set was built from 36 LRS samples which were sequenced in-house on a PromethION 24 instrument (ONT) on R9.4.1 flow cells. Reference samples were size selected using the Short Read Eliminator XS kit (PacBio) and prepared for sequencing with the SQK-LSK109 kit (ONT). Reference samples were further processed according to the workflow described in this study and had an average coverage of 25.8x ± 6.0x and an average N50 of 20.8 kb ± 5.1 kb. The bin size of the reference was set to 15 kb. WisecondorX was then executed according to the manual instructions to call CNVs on the LRS data of individuals S1-6. Further filtering was applied to only retain CNV events with a log2 ratio lower than - 0.50 and higher than 0.35. Subsequently, events lying within centromeric regions were removed. CNVs were further evaluated and solely reported if present within the region of interest as defined by previous diagnostic tests. Exact commands can be found in supplementary methods. Intermediate filtering results are listed in supplementary table S2.

#### Phasing through single nucleotide detection

Phasing was applied to samples where the SVs identified through LRS could not be accurately reconstructed into one haplotype. Basecalled fastq files were filtered on a q-score of 10 or higher with NanoFilt v2.8.0 [30] and subsequently mapped with minimap2 v2.24 [27] to the hg38 reference genome. SNVs were then called through Clair3 v1.0.2 [36]. The resulting variant file was used as input for WhatsHap v1.7 [37] to phase the long reads into haplotype groups. Aligned reads were haplotagged with the same algorithm to allow visualization in IGV v2.13.2 [34]. Detailed information on the used commands can be found in supplementary methods.

### Optical genome mapping

#### Sample preparation and molecule imaging

OGM was applied to a subset of individuals (S3,5,6) to verify the arrangement detected through LRS and to provide further phasing information. Ultra HMW DNA was extracted from frozen cell pellets (- 80°C) gathered from lymphoblastoid cell lines according to manufacturers’ instructions in the SP Frozen Cell Pellet Isolation Protocol and the SP Blood and Cell Culture DNA isolation kit (Bionano Genomics). In short, frozen samples were thawed in a 37°C warm water bath and approximately 1.5 million cells were collected. Cells were centrifuged at 2200 g for 2 minutes at 4°C and subsequently washed with cold DNA stabilizing buffer (Bionano Genomics). Cells were then digested in the presence of proteinase K, and lysate was transferred to a nanobind disk. Amer washing, HMW DNA was eluted and incubated overnight at 25°C to homogenize. For each sample 750 ng of extracted DNA was labelled using the Direct Labeling Enzyme (DLE-1) following the Direct Label and Stain kit protocol (Bionano Genomics). This DLE-1 enzyme tags the 6 bp CTTAAG sequence that occurs approximately every 5 kb in the human genome with a green fluorophore. The sample was further stained for backbone for molecule imaging. The instrument had a run time between 8 to 16 hours, depending on if the manufacturer’s minimum aim of 320 Gb throughput was obtained, except for sample S5 which ran for 72 hours. DNA was quantified amer extraction and labelling using respectively the dsDNA BR and HS Assay kit (Thermo Fisher) on a Qubit fluorometer (Thermo Fisher). Mixing of samples was done using a wide bore pipette tip to maximize the preservation of long DNA fragments. Quality metrics were assessed according to the manufacturer’s guidelines and can be found in supplementary table S1.

#### De novo assembly and structural variant detection

Molecule data from the Saphyr instrument were analyzed using the Bionano Solve v3.5.1 tool (Bionano Genomics). The De Novo Assembly pipeline v10322 was run on all three OGM samples, where a diploid assembly is constructed from the analyzed molecules and subsequently aligned to the hg38 reference optical map. Structural aberrations are then detected by identifying differences between the tagged sequence motifs of the two aligned genomes. Subsequent visualization, filtering, and interpretation of the identified SVs was done with Bionano Access v1.7.2 (Bionano Genomics). Standard filter sehngs were adjusted to select variants only present in the regions of interest as identified through previous standard diagnostic tests and in less than 1% of control samples. The total number of SVs identified in a sample was determined from the variant vcf-files as outputted by the De Novo Assembly Pipeline without any additional data filtering.

### Variant confirmation

The nanopore LRS data of variants of interest were manually inspected in IGV v2.13.2 [34] to determine the exact breakpoint locations of each variant. Genomic coordinates were set to the first nucleotide immediately upstream of where the majority of the LRS reads switched reference genome mapping locations, i.e. from local mapping to the reference genome to mapping to a distal location. Simple structural variation events (S1,2) were then confirmed with Sanger sequencing. For SVs within a CGR and identified through LRS alone or without parallel strong OGM data support, an additional confirmation through PCR and resulting fragment length was executed. SVs within a CGR and identified other and had the same strand orientation. Concordant events were not further confirmed through an additional technique. SVs identified through OGM without parallel LRS data support were not encountered in this study. The aberrant haplotypes in samples with a CGR were reconstructed manually by finding the path that leads to one aberrant chromosome and one structurally normal chromosome. Resulting subway plots were drawn by hand using Inkscape v1.2 [38].

### Assessment of structural variant origins

For each variant a 50 bp consensus sequence surrounding the SV breakpoint was extracted from the nanopore LRS data. Similarly, a 50 bp sequence surrounding the proximal and distal SV breakpoint was selected from the reference genome. These three sequences were aligned to each other using Clustal Omega v1.2.4 [39] to determine microhomology patches. The regions were extended with another 100 to 200 bp if no microhomology between the three sequences could be detected. If insertions larger than 20 bp were found between breakpoint junctions, this sequence was compared against human reference genome hg38 through blastn [40] to find its initial origin. Furthermore, the genomic region around the breakpoints was investigated for potential repeat elements using the UCSC Genome Browser [41].

### Technology evaluation

The accuracy on breakpoint locations of variants of interest was evaluated for both LRS and OGM. For LRS the breakpoint coordinates as identified through Sniffes2 SV calling were compared to the manually verified variant breakpoint coordinates. For OGM data the breakpoint locations as seen in the Bionano Access somware were compared to the nearest manually verified variant breakpoint coordinates with equal strand orientation. Breakpoint locations were manually extracted as an OGM read can span multiple variants, yet only reports two breakpoints per molecule. The manually extracted OGM breakpoints were then set to the genomic position of the last fluorophore tag before the successive reference mapping was disrupted. Comparison between nanopore LRS and Bionano OGM variant data was achieved by comparing the breakpoint coordinates for each aberrant fragment recombination.

## Data Availability

The data that support the findings of this study are not openly available due to reasons of sensitivity and are available from the corresponding author upon reasonable request. Source code regarding the bioinformatics analyses conducted in this study can be consulted in supplementary methods.

## Acknowledgements

We are grateful to all the individuals and their families who participated in this study. Furthermore, we thank Professor S. Vergult and doctor H. Syryn for their fruiaul and insighaul discussions and their support during manuscript preparation.

## References

1. Weischenfeldt, J., Symmons, O., Spitz, F. & Korbel, J. O. Phenotypic impact of genomic structural variation: Insights from and for human disease. Nat. Rev. Genet. 14, 125– 138 (2013).

2. Porubsky, D. & Eichler, E. E. A 25-year odyssey of genomic technology advances and structural variant discovery. Cell 187, 1024–1037 (2024).

3. Gröbner, S. N. et al. The landscape of genomic alterations across childhood cancers. Nature 555, 321–327 (2018).

4. Sebat, J. et al. Strong association of de novo copy number mutations with autism. Science 316, 445–449 (2007).

5. Cooper, G. M. et al. A copy number variation morbidity map of developmental delay. Nat. Genet. 43, 838–846 (2011).

6. Balachandran, P. & Beck, C. R. Structural variant identification and characterization. Chromosome Res. 28, 31–47 (2020).

7. Wright, C. F., FitzPatrick, D. R. & Firth, H. V. Paediatric genomics: Diagnosing rare disease in children. Nat. Rev. Genet. 19, 253–268 (2018).

8. Ho, S. S., Urban, A. E. & Mills, R. E. Structural variation in the sequencing era. Nat. Rev. Genet. 21, 171–189 (2019).

9. Mantere, T. et al. Optical genome mapping enables constitutional chromosomal aberration detection. Am. J. Hum. Genet. 108, 1409–1422 (2021).

10. Miller, D. E. et al. Targeted long-read sequencing identifies missing disease-causing variation. Am. J. Hum. Genet. 108, 1436–1449 (2021).

11. Bocklandt, S., Hastie, A. & Cao, H. Bionano Genome Mapping: High-Throughput, Ultra-Long Molecule Genome Analysis System for Precision Genome Assembly and Haploid- Resolved Structural Variation Discovery. Adv. Exp. Med. Biol. 1129, 97–118 (2019).

12. Logsdon, G. A., Vollger, M. R. & Eichler, E. E. Long-read human genome sequencing and its applications. Nat. Rev. Genet. 21, 597–614 (2020).

13. Wang, Y., Zhao, Y., Bollas, A., Wang, Y. & Au, K. F. Nanopore sequencing technology, bioinformatics and applications. Nat. Biotechnol. 39, 1348–1365 (2021).

14. Vergult, S. et al. Mate pair sequencing for the detection of chromosomal aberrations in patients with intellectual disability and congenital malformations. Eur. J. Hum. Genet. 22, 652–659 (2014).

15. Itai, T. et al. De novo variants in CELF2 that disrupt the nuclear localization signal cause developmental and epileptic encephalopathy. Hum. Mutat. 42, 66–76 (2021).

16. Kosuthova, K. & Solc, R. Inversions on human chromosomes. Am. J. Med. Genet. 191, 672–683 (2023).

17. Carvalho, C. M. B. & Lupski, J. R. Mechanisms underlying structural variant formation in genomic disorders. Nat. Rev. Genet. 17, 224–238 (2016).

18. Schuy, J., Grochowski, C. M., Carvalho, C. M. B. & Lindstrand, A. Complex genomic rearrangements: an underestimated cause of rare diseases. Trends Genet. 38, 1134–1146 (2022).

19. Verdin, H. et al. Microhomology-Mediated Mechanisms Underlie Non-Recurrent Disease-Causing Microdeletions of the FOXL2 Gene or Its Regulatory Domain. PLoS Genet. 9, 1003358; 10.1371/journal.pgen.1003358 (2013).

20. Ottaviani, D., LeCain, M. & Sheer, D. The role of microhomology in genomic structural variation. Trends Genet. 30, 85–94 (2014).

21. Korbel, J. O. & Campbell, P. J. Criteria for inference of chromothripsis in cancer genomes. Cell 152, 1226–1236 (2013).

22. Madan, K., Nieuwint, A. W. M. & Van Bever, Y. Recombination in a balanced complex translocation of a mother leading to a balanced reciprocal translocation in the child. Review of 60 cases of balanced complex translocations. *Hum. Genet.* **99**, 806–815 (1997).

23. Gajecka, M. et al. Unexpected complexity at breakpoint junctions in phenotypically normal individuals and mechanisms involved in generating balanced translocations t(1;22)(p36;q13). Genome Res. 18, 1733–1742 (2008).

24. Eisfeldt, J. et al. Hybrid sequencing resolves two germline ultra-complex chromosomal rearrangements consisting of 137 breakpoint junctions in a single carrier. Hum. Genet. 140, 775–790 (2021).

25. Koltsova, A. S. et al. On the complexity of mechanisms and consequences of chromothripsis: An update. Front. Genet. 10, 446661; 10.3389/fgene.2019.00393 (2019).

26. Collins, R. L. et al. A structural variation reference for medical and population genetics. Nature 581, 444–451 (2020).

27. Li, H. Minimap2: Pairwise alignment for nucleotide sequences. BioinformaTcs 34, 3094–3100 (2018).

28. Danecek, P. et al. Twelve years of SAMtools and BCFtools. GigaScience 10, 33590861; 10.1093/gigascience/giab008 (2021).

29. Pedersen, B. S. & Quinlan, A. R. Mosdepth: quick coverage calculation for genomes and exomes. BioinformaTcs 34, 867–868 (2018).

30. De Coster, W. & Rademakers, R. NanoPack2: population-scale evaluation of long-read sequencing data. BioinformaTcs 39, (2023).

31. Leger, A. & Leonardi, T. pycoQC, interactive quality control for Oxford Nanopore Sequencing. J. Open Source SoVw. 4, 1236 (2019).

32. Smolka, M. et al. Detection of mosaic and population-level structural variants with Sniffes2. Nat. Biotechnol. 2, 38168980; 10.1038/s41587-023-02024-y (2024).

33. Danecek, P. et al. The variant call format and VCFtools. BioinformaTcs 27, 2156–2158 (2011).

34. Robinson, J. T. et al. Integrative genomics viewer. Nat. Biotechnol. 29, 24–26 (2011).

35. Raman, L., Dheedene, A., De Smet, M., Van Dorpe, J. & Menten, B. WisecondorX: improved copy number detection for routine shallow whole-genome sequencing. Nucleic Acids Res. 47, 1605–1614 (2019).

36. Zheng, Z. et al. Symphonizing pileup and full-alignment for deep learning-based long- read variant calling. Nat. Comput. Sci. 2, 797–803 (2022).

37. Martin, M., et al. WhatsHap: fast and accurate read-based phasing. Preprint at: https://www.biorxiv.org/content/10.1101/085050v2 (2016).

38. Inkscape Project. Inkscape. https://inkscape.org.

39. Sievers, F. et al. Fast, scalable generation of high-quality protein multiple sequence alignments using Clustal Omega. Mol. Syst. Biol. 7, 539 (2011).

40. Zhang, Z., Schwartz, S., Wagner, L. & Miller, W. A greedy algorithm for aligning DNA

41. Kent, W. J. et al. The Human Genome Browser at UCSC. Genome Res. 12, 996–1006 (2002).

