## Supplemental tables for "Full characterization of unresolved structural variation through long-read sequencing and optical genome mapping": captions.docx

**Table S1** Information on start materials, processing and quality statistics, and total number of structural variants of the LRS and OGM analyses for individuals S1-6.

**Table S2** Intermediate filtering steps and final results for the LRS CNV calling through WisecondorX in individuals S1-6.

**Table S3** Exact breakpoint coordinates along with relevant genomic annotation in the previously determined regions of interest in individuals S1-6. Regions of interest were broadened upon the validated finding of a cryptic variant through either LRS or OGM.

**Table S4** List of genes implicated by the rearrangements found in individuals S1-6, along with their genomic locations and associated OMIM phenotypes.

**Table S5** PCR primers and expected and actual fragment size of the validated variants in individuals S3-6.

**Table S6** Variants as identified by Sniffles2 through LRS analysis in individual S1.

**Table S7** Variants as identified by Sniffles2 through LRS analysis in individual S2.

**Table S8** Variants as identified by Sniffles2 and Access for respectively LRS and OGM analysis in individual S3.

**Table S9** Variants as identified by Sniffles2 through LRS analysis in individual S4.

**Table S10** Variants as identified by Sniffles2 through LRS analysis in individual S5.

**Table S11** Variants as identified by Sniffles2 and Access for respectively LRS and OGM analysis in individual S6.

**Table S12** Calculation of the number of total breakpoints seen with each technology, and their mean deviation from the actual breakpoint coordinates.
