## Supplemental figures for "Full characterization of unresolved structural variation through long-read sequencing and optical genome mapping"

Griet De Clercq[1,2], Lies Vantomme[1], Barbara Dewaele[3], Bert Callewaert[1,2], Olivier Vanakker[1,2], Sandra Janssens[1,2], Bart Loeys[4], Mojca Strazisar[5.6], Wouter De Coster[5,6], Joris Robert Vermeesch[3,7], Annelies Dheedene[2], Björn Menten*[1,2]

1. Department of Biomolecular Medicine, Ghent University, Ghent, Belgium
2. Center for Medical Genetics Ghent, Ghent University Hospital, Ghent, Belgium
3. Center for Human Genetics Leuven, University Hospital Leuven, Leuven, Belgium.
4. Center for Medical Genetics Antwerp, University of Antwerp, Antwerp University Hospital, Antwerp, Belgium.
5. VIB Center for Molecular Neurology, VIB, Antwerp, Belgium
6. Department of Biomedical Sciences, University of Antwerp, Antwerp, Belgium
7. Department of Human Genetics, KU Leuven, Leuven, Belgium.

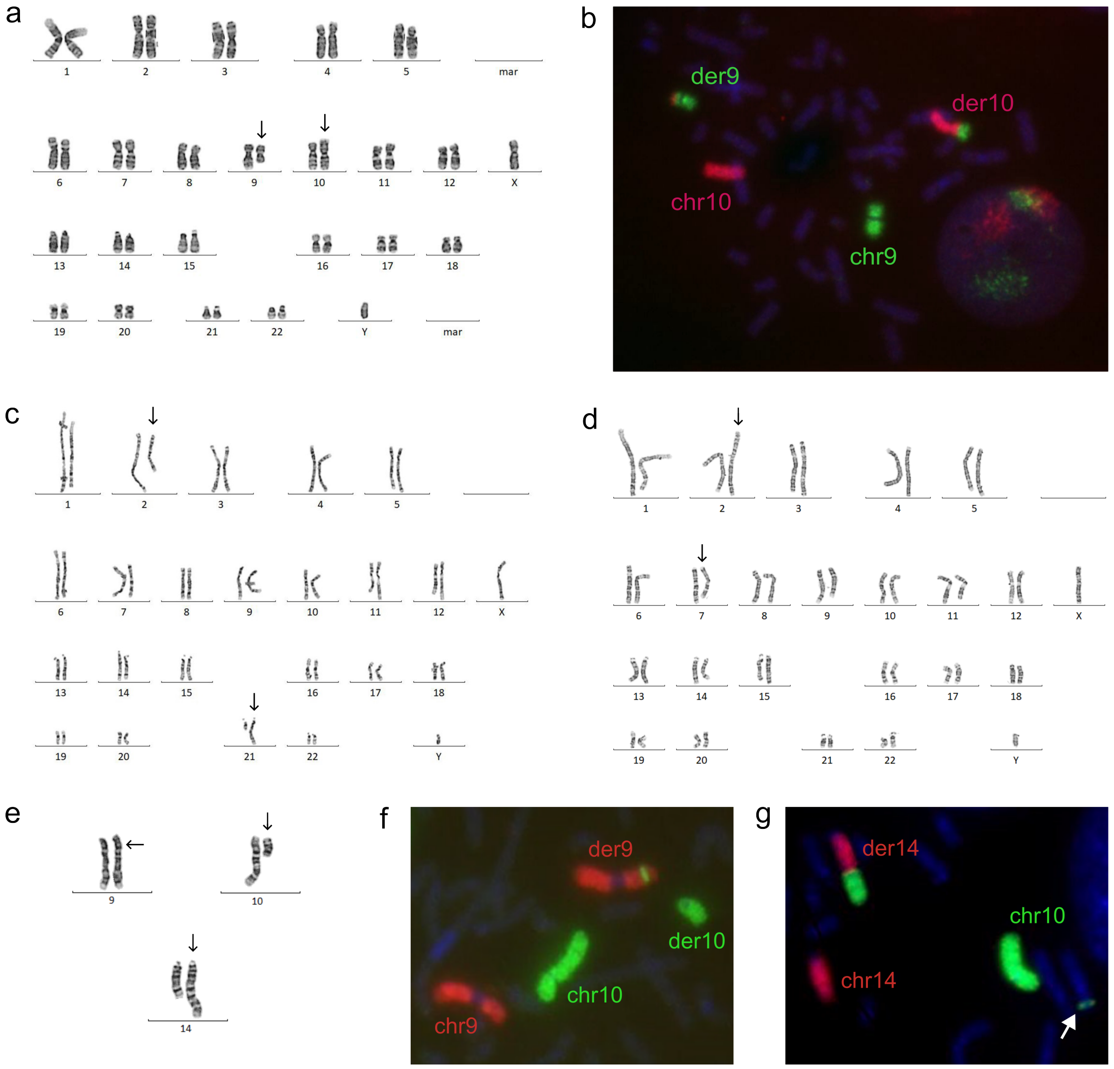


**Fig. S1** Clinically relevant results from previously conducted karyotype and FISH analyses in individuals S1, S3, S5, and S6. Derivative chromosomes in karyotype visualizations are indicated by black arrows. **(a)** Karyotyping in individual S1 identified a t(9;10)(q21.2;p15.2) variant. **(b)** Confirmation of the t(9;10)(q21.2;p15.2) translocation in individual S1 through FISH using whole chromosome probes for chromosome 9 and 10. **(c)** Karyotyping in individual S3 identified a t(2;21)(q22;q21) variant. Chromosome 5, later diagnosed through OGM and LRS as part of a complex rearrangement seen in this individual, displayed no visual aberrations. **(d)** Karyotyping in individual S5 identified a t(2;7)(q36;q32) variant. **(e)** Karyotyping in individual S6 identified the aberrant 46,XX,ins(9;10)(p22;q11.2q21.2),t(10;14)(q?;q32.3) karyotype. **(f)** Confirmation of the ins(9;10)(p22;q11.2q21.2) variant in individual S6 through FISH using whole chromosome probes for chromosome 9 and 10. In der9 the chromosome 10 insertion into chromosome 9 can be seen. The chromosome 10 part of the t(10;14)(q?;q32.3) variant is shown as well in der10. **(g)** Confirmation of the t(10;14)(q?;q32.3) variant in individual S6 through FISH using whole chromosome probes for chromosomes 10 and 14. The translocation is indicated as der14 in the figure. The white arrow indicates the segment of chromosome 10 inserted into chromosome 9 due to the ins(9;10)(p22;q11.2q21.2) variant also identified in this sample.

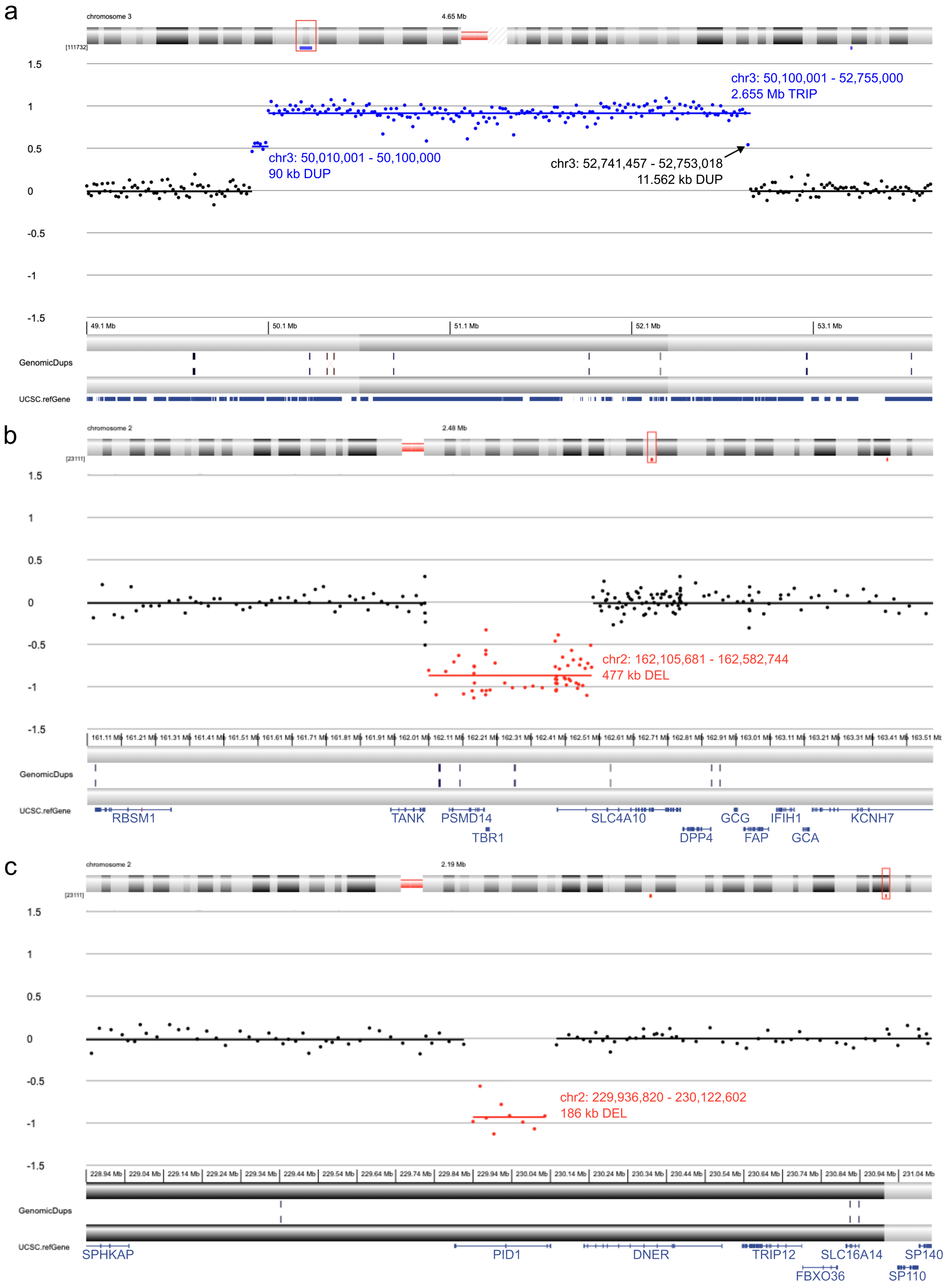


**Fig. S2** Visualization of the clinically relevant results found through previously conducted CNV analyses in individuals S4 (a) and S5 (b,c). Reported coordinates are on genome build hg38 unless stated otherwise. **(a)** Shallow WGS identified a 90 kb duplication and a 2.6 Mb triplication at chromosome bands 3p21.31p21.1 in individual S4. Proximal of the 2.6 Mb triplication a single reporter (indicated by a black arrow) can be seen that indicates the presence of the 11.6 kb duplication found through LRS reconstruction. **(b)** Microarray analysis identified a 477 kb deletion at chromosome band 2q24.2 in individual S5. This deletion encompasses the *TBR1* gene responsible for the observed phenotype. Coordinates are on genome build hg19. **(c)** Microarray analysis identified a 186 kb deletion at chromosome band 2q36.3 in individual S5. This deletion was suspected to be linked to the t(2;7)(q36;q32) variant identified through karyotyping in the same individual. Coordinates are on genome build hg19.

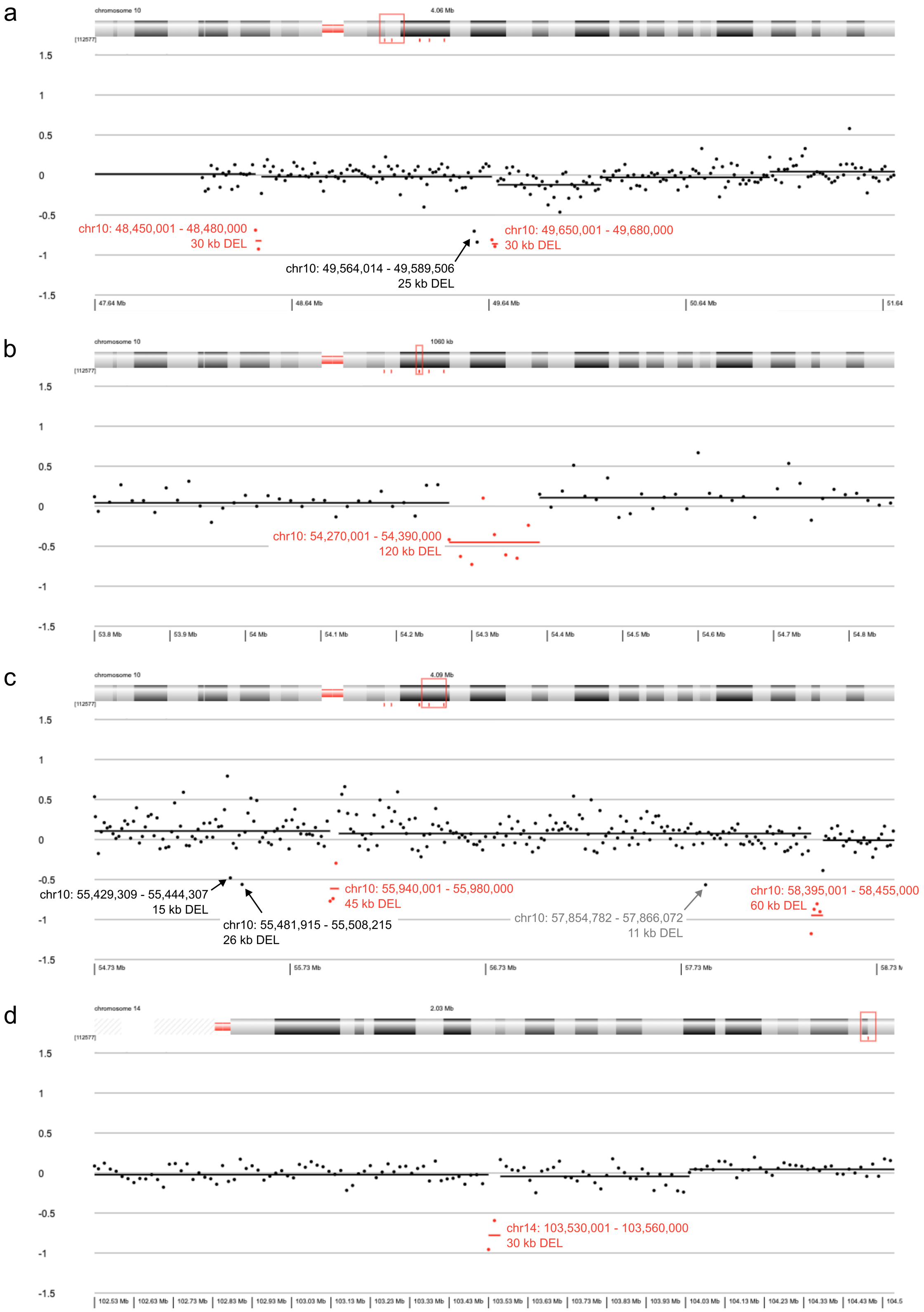


**Fig. S3** Visualization of the clinically relevant results found through shallow WGS analysis previously conducted on individual S6. In total six deletions larger than 15 kb have been identified in the regions of interest (depicted red). Reported coordinates are on genome build hg38. **(a)** Region chr10:47640000-51640000. Two deletions have been called through shallow WGS (red), while LRS discovered three deletions (red + black). **(b)** Region chr10:53800000-54800000. One large 120 kb deletion was called through shallow WGS (red) which LRS eventually identified as two smaller deletions of respectively 40 and 37 kb. **(c)** Region chr10:54730000-58730000. Shallow WGS identified two deletions in this region (red) while four deletions larger than 15 kb have been detected through LRS (red + black). A third, smaller deletion (gray; 11 kb) can also be seen in the shallow WGS data that was identified through CGR reconstruction. **(d)** Region chr14:102530000-104500000. One deletion has been detected through both LRS and shallow WGS (red).

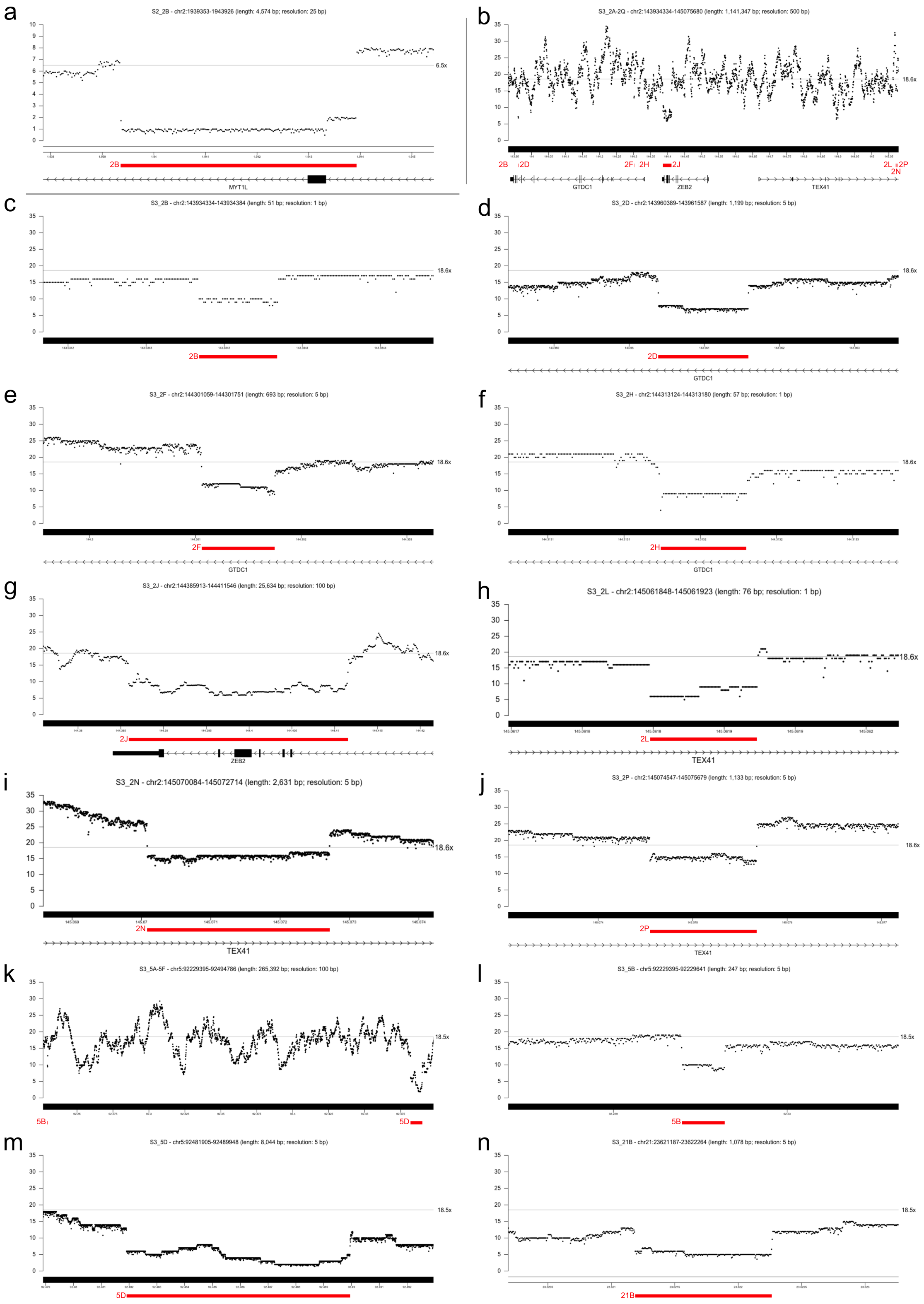


**Fig. S4** Coverage plots from the LRS data at the CNV regions detected in individuals S2 (a) and S3 (b-n). Average chromosome depths are indicated by a horizontal gray line, with the corresponding coverage on the right.

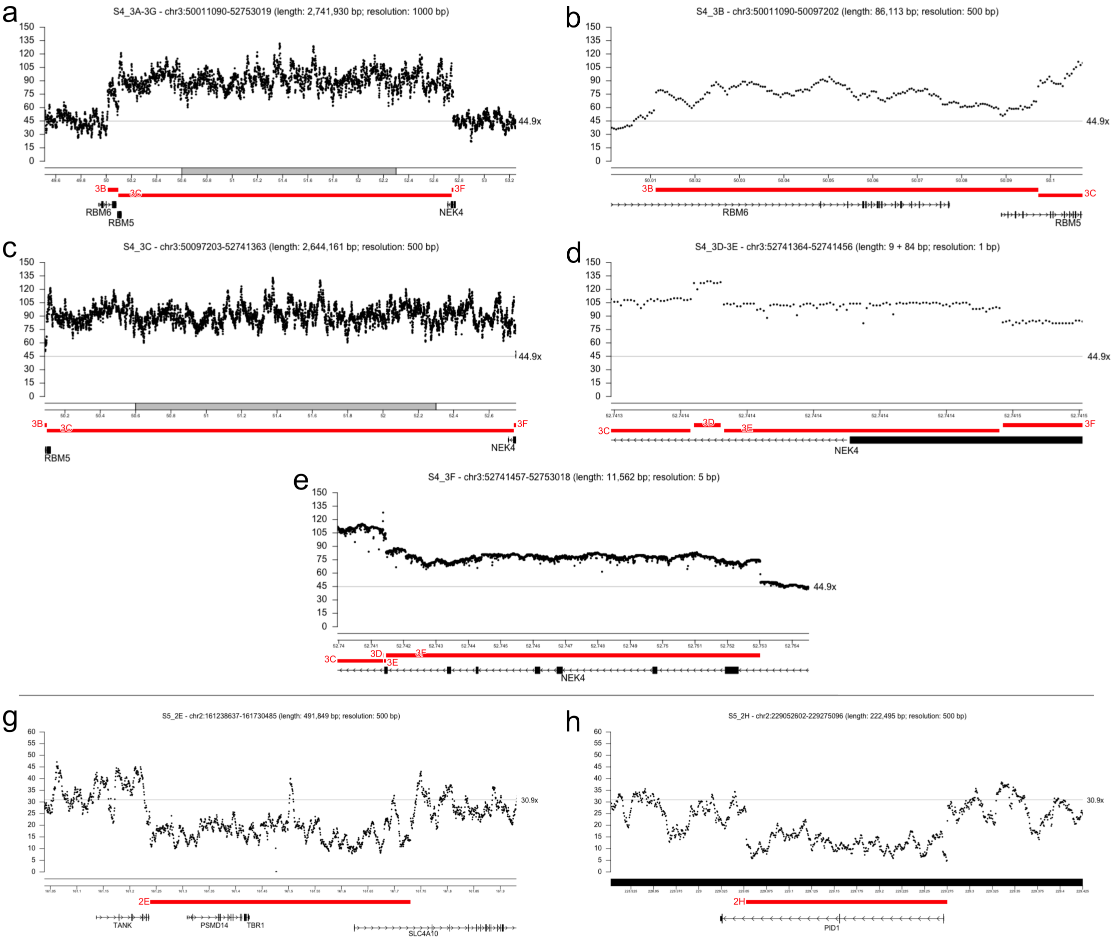


**Fig. S5** Coverage plots from the LRS data at the CNV regions detected in individuals S4 (a-e) and S5 (g,h). Average chromosome depths are indicated by a horizontal gray line, with the corresponding coverage on the right.

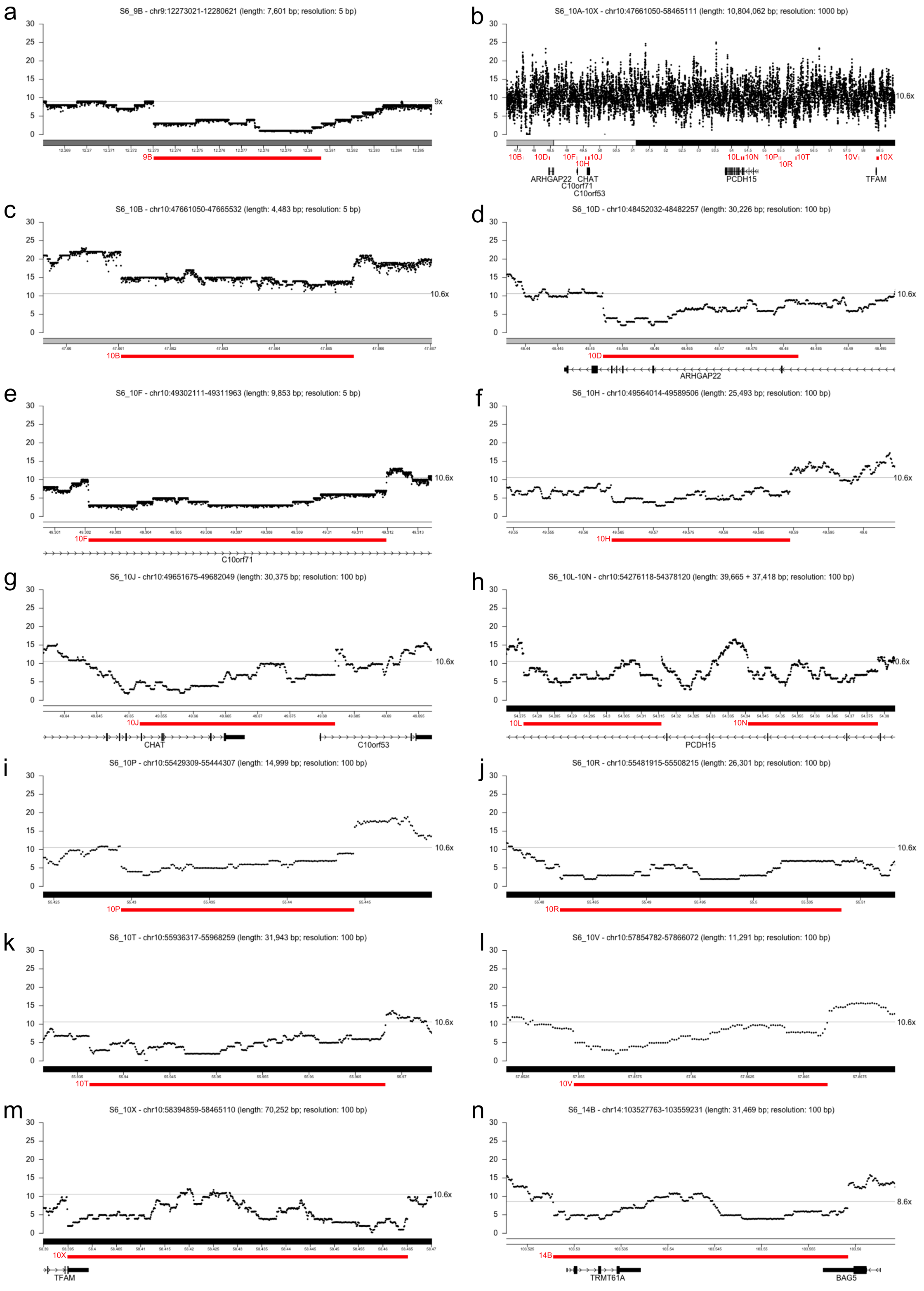


**Fig. S6** Coverage plots from the LRS data at the CNV regions detected in individual S6. Average chromosome depths are indicated by a horizontal gray line, with the corresponding coverage on the right.

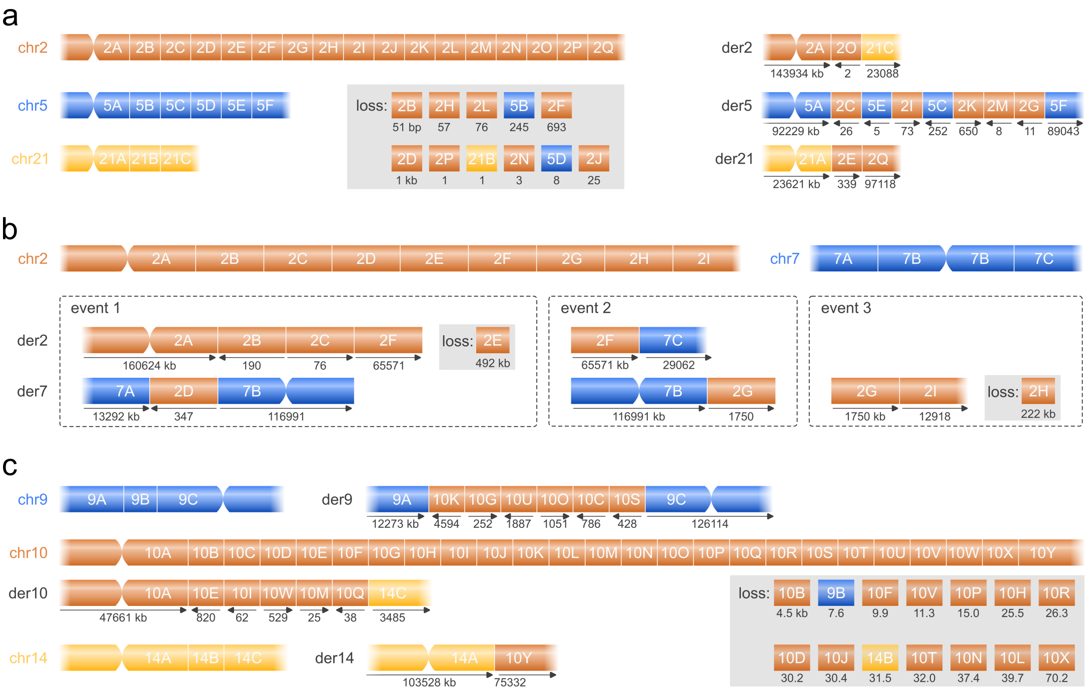


**Fig. S7** Reconstructed haplotypes of the CGRs found in individuals S3, S5, and S6. The normal chromosomes show the different fragments created by the CGR, while the derivatives show how the separate fragments have been reconstructed back into the aberrant haplotypes along with their orientation. Deleted fragments are shown in gray boxes. **(a)** Aberrant haplotypes in individual S3. This shows the translocation between chromosomes 2 and 21, along with the disorganized insertions of chromosomes 2 into chromosome 5. Eleven fragments have been deleted, although the losses are minor covering only a few hundreds of bp in length, and only fragment 2J measuring over 15 kb. **(b)** Aberrant haplotypes in individual S5. The different events could not be further phased and are therefore visualized as separate events. The der2 and der7 however show the potential derivative chromosomes if all events are lying in *cis*. **(c)** Aberrant haplotypes in individual S6. This shows the translocation between chromosomes 10 and 14, along with the complex insertion of multiple reshuffled fragments of chromosome 10 into chromosome 14. Fourteen fragments have been deleted, of which ten measure over 15 kb.

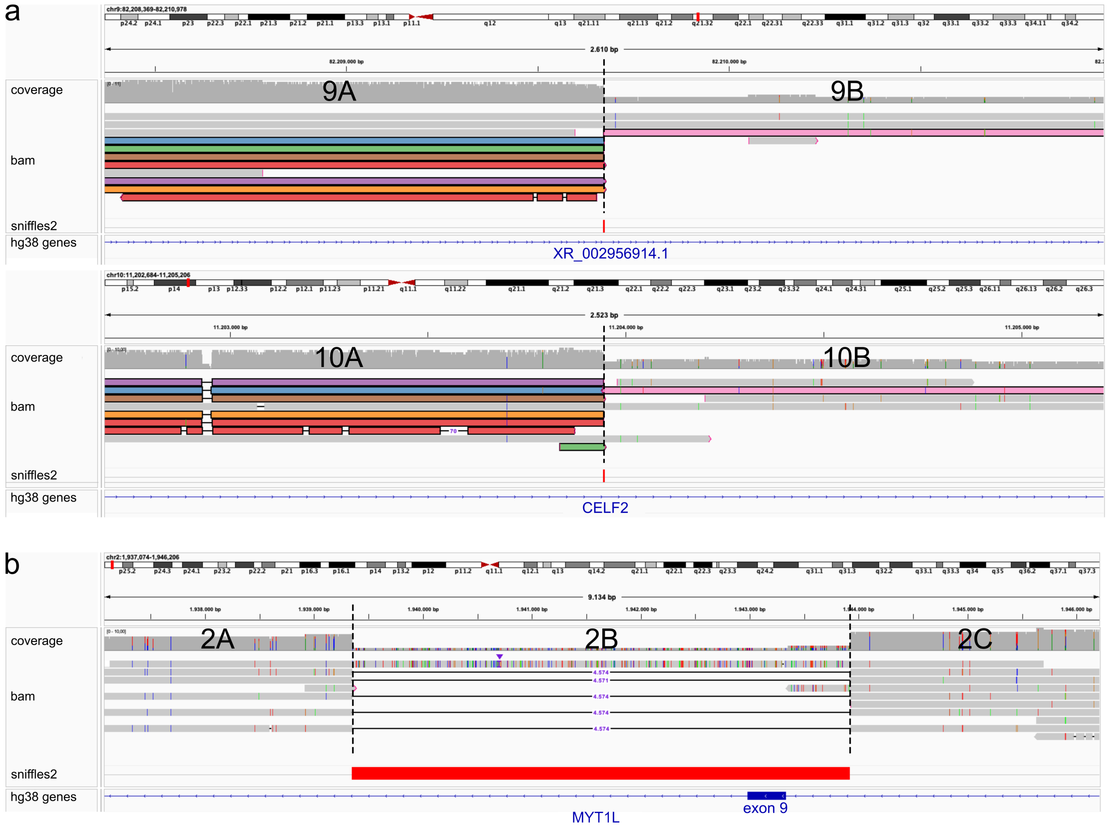


**Fig. S8** Clinically relevant SVs in individual S1 (a) and S2 (b) identified through LRS. **(a)** LRS data from individual S1 at the translocation breakpoints in chromosome 9 (top) and chromosome 10 (bottom). Reads with the same identifier are visualized in the same color, showing how reads from the aberrant chromosomes map to two different locations in the genome. The dashed lines show the breakpoint locations, which fall within the *CELF2* gene on chromosome 10. Sniffles2 called the breakpoints as two 'BND' variants, which can be seen as the red rectangles at the 'sniffles2' track. **(b)** LRS data from individual S2 spanning the 4,574 bp deletion (fragment 2B) in the *MYT1L* gene enclosing exon 9. The dashed lines show the breakpoint locations. Sniffles2 called a deletion over the 2B fragment, seen as a large red rectangle at the 'sniffles2' track. Due to low coverage of this sample only limited normal reads are seen.

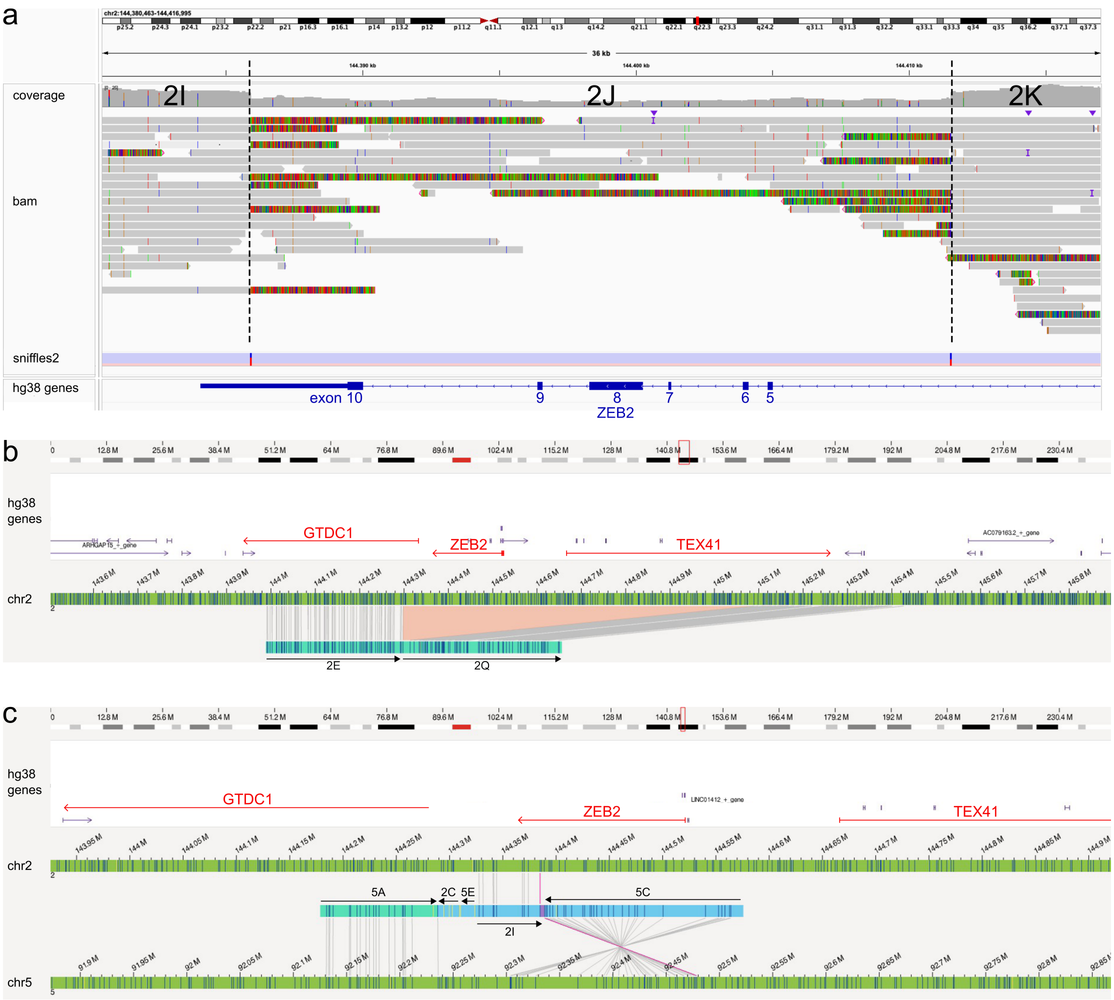


**Fig. S9** Disruption of the *ZEB2* region in individual S3 seen through LRS and OGM. **(a)** IGV screenshot of the LRS data featuring the 25 kb deletion in *ZEB2* (fragment 2J). The breakpoints are illustrated by the dashed lines. Green fluorescent fragments (soft-clipped bases) indicate that this part of the read maps to elsewhere in the genome, and coincide with the identified breakpoints. Sniffles2 calls the deletion breakpoints as two BND variants, seen as the red/blue rectangles at the 'sniffles2' track. In the 'hg38 genes' track you can see how the 2J deletion removes exon 5-10 of the *ZEB2* gene. **(b)** OGM molecule covering the 2E-2Q fragments (indicated by black arrows) of derivative 21. This shows how *GTDC1* and *TEX41* are directly disrupted, and how *ZEB2* is completely deleted in this fragment. **(c)** OGM molecule covering the 5A-2C-5E-2I-5C fragments (indicated by black arrows) of derivative 5. The 5A, 2I and 5C fragments can be located back to the human reference optical map as indicated by the connecting lines. The 2C and 5E fragments however lack sufficient fluorescent tags to unambiguously align back, and as such are seen as unmapped parts in the OGM molecule. This molecule shows how *ZEB2* is directly affected by a breakpoint

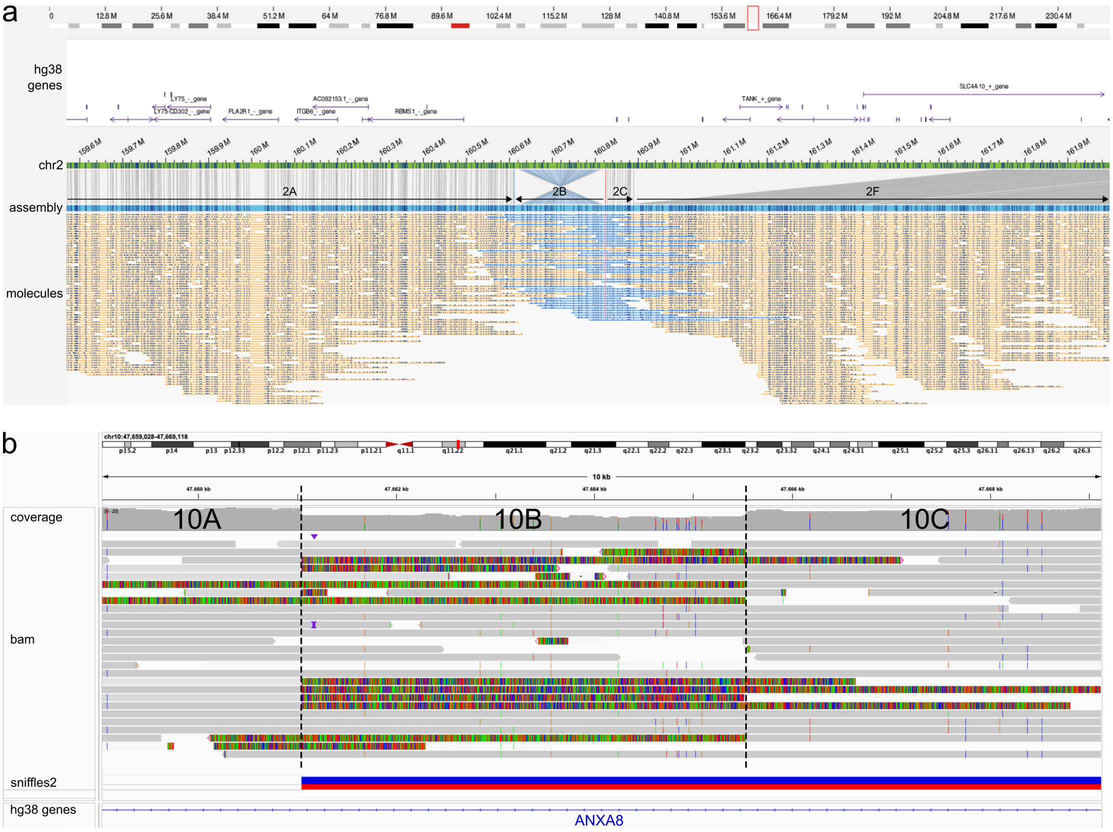


**Fig. S10** OGM and LRS data from individual S5 (a) and individual S6 (b). **(a)** OGM data from individual S5 centered around the 2B inversion fragment in event 1 (fig. 3a,b). This shows the reconstructed assembly, with underneath the separate OGM molecules on which the assembly is based. Through this assembly the 2B-2C, and 2F fragments could be identified as lying within the same haplotype, something which was not feasible through LRS data due to lack of informative SNVs in this region. The blue highlighted molecules all align to the red highlighted tag. Here you can see how most of these molecules map to all three 2B, 2C, and 2F fragments at once and as such these fragments can be phased as one large event. **(b)** LRS data from individual S6 centered around the 10B fragment in chromosome 10 shows how some variants stay unreported by Sniffles2, yet can still be picked up manually from the data. In the aberrant haplotypes this region features the 5 kb deleted 10B fragment, along with flanking translocations of 10A and 10C to elsewhere in the genome. As a read will cover such a junction, it will switch mapping locations on the genome. This can be clearly seen in the data through the fluorescent green parts (soft-clipped bases) of the reads. Breakpoint locations are indicated by the dashed lines. Sniffles2 indeed calls a variant at the first breakpoint, seen as a blue/red rectangle at the 'sniffles2' track. However, sniffles2 does not identify the variant at the second dashed line nor the 5 kb deletion in between. The second breakpoint can however still be manually extracted due to the green fluorescent signatures seen in the LRS data and the accompanying coverage drop.
