## Supplemental text for "Full characterization of unresolved structural variation through long-read sequencing and optical genome mapping"

Griet De Clercq[1,2], Lies Vantomme[1], Barbara Dewaele[3], Bert Callewaert[1,2], Olivier Vanakker[1,2], Sandra Janssens[1,2], Bart Loeys[4], Mojca Strazisar[5.6], Wouter De Coster[5,6], Joris Robert Vermeesch[3,7], Annelies Dheedene[2], Björn Menten*[1,2]

1. Department of Biomolecular Medicine, Ghent University, Ghent, Belgium
2. Center for Medical Genetics Ghent, Ghent University Hospital, Ghent, Belgium
3. Center for Human Genetics Leuven, University Hospital Leuven, Leuven, Belgium.
4. Center for Medical Genetics Antwerp, University of Antwerp, Antwerp University Hospital, Antwerp, Belgium.
5. VIB Center for Molecular Neurology, VIB, Antwerp, Belgium
6. Department of Biomedical Sciences, University of Antwerp, Antwerp, Belgium
7. Department of Human Genetics, KU Leuven, Leuven, Belgium.

#### Table of contents


**Individual S1** **2**

**Individual S2** **5**

**Individual S3** **6**

**Individual S4** **20**

**Individual S5** **24**

**Individual S6** **30**

### Individual S1

#### Reference sequences

9A-9B – chr9:82209673-82209674
>hg38_dna range=chr9:82209598-82209749 5'pad=75 3'pad=75 strand=+
repeatMasking=lower
catttcatttattgtacttctcagtttcagaatttttttttaaattcaat
ctctgttaaatttctcaatttgttcc|atgtattattttcctgattttatt
gaattgtttctctatattttcttaatgttcactgaacttccttaaaacag
at

>hg38_dna range=chr9:82209598-82209749 5'pad=75 3'pad=75 strand=-
repeatMasking=lower
atctgttttaaggaagttcagtgaacattaagaaaatatagagaaacaat
tcaataaaatcaggaaaataatacat|ggaacaaattgagaaatttaacag
agattgaatttaaaaaaaaattctgaaactgagaagtacaataaatgaaa
tg

10A-10B – chr10:11203945-11203946
>hg38_dna range=chr10:11203870-11204021 5'pad=75 3'pad=75 strand=+
repeatMasking=lower
GATACTACAGCGTAAATGGTTGGGAAACAGATTGCGGTGTTGTGTCTTCT
GAGGCATTTTTATTTTAATATGAATC|TGAGCCTCCAACTGAAAGGATGAA
GCTCAGAGGCTGAGTGGAGAGACTGGGGGAGAGGACAGAAGAGGAGGGGC
GC

>hg38_dna range=chr10:11203870-11204021 5'pad=75 3'pad=75 strand=-
repeatMasking=lower
GCGCCCCTCCTCTTCTGTCCTCTCCCCCAGTCTCTCCACTCAGCCTCTGA
GCTTCATCCTTTCAGTTGGAGGCTCA|GATTCATATTAAAATAAAAATGCC
TCAGAAGACACAACACCGCAATCTGTTTCCCAACCATTTACGCTGTAGTA
TC

#### Variants

### 9A-10A

-> <-

Expected:
catttcatttattgtacttctcagtttcagaatttttttttaaattcaat
ctctgttaaatttctcaatttgttcc|GATTCATATTAAAATAAAAATGCC
TCAGAAGACACAACACCGCAATCTGTTTCCCAACCATTTACGCTGTAGTA
TC

LRS:
CATTTCATTTATTGTACTTCTCAGTTTCAGAATTTTTTTTTAAATTCAAT
CTCTGTTAAATTTCTCAATTTGTTCC|GATTCATATTAAAATAAAAATGCC
TCAGAAGACACAACACCGCAATCTGTTTCAACCATTTACGCTGTAGTATC
AA

Sanger:
CATTTCATTTATTGTACTTCTCAGTTTCAGAATTTTTTTTTAAATTCAAT
CTCTGTTAAATTTCTCAATTTGTTCC|GATTCATATTAAAATAAAAATGCC

TCAGAAGACACAACACCGCAATCTGTTTCCCAACCATTTACGCTGTAGTA
TC

Microhomology:

```
9A-9B                tctgttaaatttctcaa---tttgttccatgtattattttcctgattttattg
LRS                  TCTGTTAAATTTCTCAA---TTTGTTCCGATTCATATTAAAATAAAAATGCCT
sanger               TCTGTTAAATTTCTCAA---TTTGTTCCGATTCATATTAAAATAAAAATGCCT
10A-10B_reverse      ---CTTCATCCTTTCAGTTGGAGGCTCAGATTCATATTAAAATAAAAATGCCT
                         ** *   * ***       * **    *  ****    * *   *
```

# 9B-10B

<- ->

Expected:

atctgttttaaggaagttcagtgaacattaagaaaatatagagaaacaat
tcaataaaatcaggaaaataatacat|TGAGCCTCCAACTGAAAGGATGAA
GCTCAGAGGCTGAGTGGAGAGACTGGGGGAGAGGACAGAAGAGGAGGGGC
GC

LRS:

TCTGTTTTAAGGAAGTTCAGTGAACATTAAGAAAATATAGAGAAACAATT
CAATAAAATCAGGAAAATAATACATG|TGAGCCTCCAACTGAAAGGATGAA
GCTCAGAGGCTGAGTGGAAAGACTGGGGAGAGGACAGAAGAGGAGGGGCG
CA

Sanger:

TCTGTTTTAAGGAAGTTCAGTGAACATTAAGAAAATATAGAGAAACAATT
CAATAAAATCAGGAAAATAATACATG|TGAGCCTCCAACTGAAAGGATGAA
GCTCAGAGGCTGAGTGGAAAGACTGGGGGAGAGGACAGAAGAGGAGGGGC
GC

Microhomology:

```
9A-9B_reverse      caataaaatcaggaaaataatacatggaa----caaattgagaaatttaacaga
LRS                -AATAAAATCAGGAAAATAATACATGTGAGCCTCCAACTGAAAGGATGAAG---
sanger             -AATAAAATCAGGAAAATAATACATGTGAGCCTCCAACTGAAAGGATGAAG---
10A-10B            -AGGCATTTTTATTTTAATATGAATCTGAGCCTCCAACTGAAAGGATGAAG---
                    *   *  *       *  **  **   *    * ** *** *   * **
```

### Individual S2

#### Reference sequences

2A-2B – chr2:1939352-1939353

```
>hg38_dna range=chr2:1939277-1939428 5'pad=75 3'pad=75 strand=+
repeatMasking=lower
GAATCGGTTGCTGGCCCAGGGACATAGCAGTCACTAAGAGTGTCACCCAA
GGAACATGAAAGGCCAGCCTCGGGAT|GCTGACCCAGAACTCCAGCGATCA
GAGGAATGGGACCCCCCAGCTTTTAGGGAGTGCATGGTTTCTGCCAAGCC
AC
```

2B-2C – chr2:1943926-1943927

```
>hg38_dna range=chr2:1943851-1944002 5'pad=75 3'pad=75 strand=+
repeatMasking=lower
AGACTTGATAACCTCCTGAAACAAGATGAGCCCTAGGGCTTGTCAATGTT
GTATGAAAGCAACTGCATCATTCAAA|GTTCTGCAGAACTTCCTCAGTCTT
AGATGTTATTAGTCCCACGCCAGCATTCCAGAAAACAACTTCAGTGTTAG
CG
```

#### Variants

### 2A-2C

```
-> ->
```

Expected:

```
GAATCGGTTGCTGGCCCAGGGACATAGCAGTCACTAAGAGTGTCACCCAA
GGAACATGAAAGGCCAGCCTCGGGAT|GTTCTGCAGAACTTCCTCAGTCTT
AGATGTTATTAGTCCCACGCCAGCATTCCAGAAAACAACTTCAGTGTTAG
CG
```

LRS:

```
AATCGGCTGCTGGGCCCAGGGACATAGCAGTCACTAAGAGTGTCACCCAA
GGAACATGAAAGGCCAGCCTCGGGAT|GTTCTGCAGAACTTCCTCAGTCTT
AGATGTTATTAGTCCCACGCCAGCATTCCAGAAAACAACTTCAGTGTTAG
CG
```

Sanger:

```
AGATCGGTTGCTGGCCCAGGGACATAGCAGTCACTAAGAGTGTCACCCAA
GGAACATGAAAGGCCAGCCTCGGGAT|GTTCTGCAGAACTTCCTCAGTCTT
AGATGTTATTAGTCCCACGCCAGCATTCCAGAAAACCCAGA
```

Microhomology:

```
2A-2B       GAACATGAAAGGCCAGCCTCGGGATGCTGACCCAGAACTCCAGCGATCAG
LRS         GAACATGAAAGGCCAGCCTCGGGATGTTCTGCAGAACTTCCTCAGTCTTA
Sanger      GAACATGAAAGGCCAGCCTCGGGATGTTCTGCAGAACTTCCTCAGTCTTA
2B-2C       TATGAAAGCAACTGCATCATTCAAAGTTCTGCAGAACTTCCTCAGTCTTA
             *  *    *       *     * * *   *   *  ***   *
```

### Individual S3

#### Reference sequences

2A-2B - chr2: 143934333-143934334
>hg38_dna range=chr2:143934258-143934409 5'pad=75 3'pad=75 strand=+ repeatMasking=lower
CAATCAAGCAACCATCATTTGCTAATAAATGTTTTTATCACAGCTTTCAA
ACCTTTTTCAAGACATCTGAGCTCAT|CTTCACTTCCTCCTTCTTGAAAAA
GAGACCCCATGGTGTGACACTCTAAAACCACCTCTGGATTTAAAACTATG
GC

>hg38_dna range=chr2:143934258-143934409 5'pad=75 3'pad=75 strand=- repeatMasking=lower
GCCATAGTTTTAAATCCAGAGGTGGTTTTAGAGTGTCACACCATGGGGTC
TCTTTTTCAAGAAGGAGGAAGTGAAG|ATGAGCTCAGATGTCTTGAAAAAG
GTTTGAAAGCTGTGATAAAAACATTTATTAGCAAATGATGGTTGCTTGAT
TG

2B-2C - chr2: 143934384-143934385
>hg38_dna range=chr2:143934309-143934460 5'pad=75 3'pad=75 strand=+ repeatMasking=lower
CCTTTTTCAAGACATCTGAGCTCATCTTCACTTCCTCCTTCTTGAAAAAG
AGACCCCATGGTGTGACACTCTAAAA|CCACCTCTGGATTTAAAACTATGG
CTGCCTGGAATTGAAGGCCCAGTTGCTGAGATTCCCTGATTTGCAAAGCA
AG

>hg38_dna range=chr2:143934309-143934460 5'pad=75 3'pad=75 strand=- repeatMasking=lower
CTTGCTTTGCAAATCAGGGAATCTCAGCAACTGGGCCTTCAATTCCAGGC
AGCCATAGTTTTAAATCCAGAGGTGG|TTTTAGAGTGTCACACCATGGGGT
CTCTTTTTCAAGAAGGAGGAAGTGAAGATGAGCTCAGATGTCTTGAAAAA
GG

2C-2D – chr2: 143960388-143960389
>hg38_dna range=chr2:143960313-143960464 5'pad=75 3'pad=75 strand=+ repeatMasking=lower
GATGACTCTTGGAAGATGGGTTGAGCTTTGACAAATAAAGGAGGAGGGAG
GGGCACTGGATGGTGAGATATGAGCC|ATCTTCAGGGAACACTGACTAATC
TAGGCTACAGTCCAGGATATAATGTTCAAGGATTGAAGTAAGGCTAGGAA
GG

>hg38_dna range=chr2:143960313-143960464 5'pad=75 3'pad=75 strand=- repeatMasking=lower
CCTTCCTAGCCTTACTTCAATCCTTGAACATTATATCCTGGACTGTAGCC
TAGATTAGTCAGTGTTCCCTGAAGAT|GGCTCATATCTCACCATCCAGTGC
CCCTCCCTCCTCCTTTATTTGTCAAAGCTCAACCCATCTTCCAAGAGTCA
TC

2D-2E – chr2: 143961587-143961588
>hg38_dna range=chr2:143961512-143961663 5'pad=75 3'pad=75 strand=+ repeatMasking=lower
ATTTATTTTCCTGTGTCTTTCCCAAGTACACATTTCTCAAACTTTGAGGA
C
AGCTTATGAAAATGACAGAGCATTT|TCCCTCAGTGATGTTTTCATATTT

ATAATTTTTGAGAACAATTTCTTGAGAGATGTTTTGGGAGCTGTATTCCA
GA

>hg38_dna range=chr2:143961512-143961663 5'pad=75 3'pad=75 strand=-
repeatMasking=lower
TCTGGAATACAGCTCCCAAAACATCTCTCAAGAAATTGTTCTCAAAAATT
ATAAATATGAAAACATCACTGAGGGA|AAATGCTCTGTCATTTTCATAAGC
TGTCCTCAAAGTTTGAGAAATGTGTACTTGGGAAAGACACAGGAAAATAA
AT

2E-2F – chr2: 144301058-144301059
>hg38_dna range=chr2:144300983-144301134 5'pad=75 3'pad=75 strand=+
repeatMasking=lower
TAGCATTTAATCTGACCATCTGTTTACCAAAGGCTAGTACATTGAAAAAA
A
GAAAGAAAATGGGTTTTTTTCCTAG|CCAATTGCAATGCCAATGTTTTAA
TCTTTATTTAAAAAAATCTGATTAAAACCAAAAATCCTTCCAATAAACCA
TT

>hg38_dna range=chr2:144300983-144301134 5'pad=75 3'pad=75 strand=-
repeatMasking=lower
AATGGTTTATTGGAAGGATTTTTGGTTTTAATCAGATTTTTTTAAATAAA
GATTAAAACATTGGCATTGCAATTGG|CTAGGAAAAAAACCCATTTTCTTT
CTTTTTTTCAATGTACTAGCCTTTGGTAAACAGATGGTCAGATTAAATGC
TA

2F-2G – chr2: 144301751-144301752
>hg38_dna range=chr2:144301676-144301827 5'pad=75 3'pad=75 strand=+
repeatMasking=lower
CCATAAGAGGCTCTTTTTAAAAGCACATCCACTACCAATGCCCACATGCT
TTCTGGAGGAAATGCAAGTATTAAAC|GCCAGGTTCCACAAACAGTGGCCT
CAAAGCACTAATCAAGACCTCTGAAGGCCACTGCCACTCCTCcaggaggc
at

>hg38_dna range=chr2:144301676-144301827 5'pad=75 3'pad=75 strand=-
repeatMasking=lower
atgcctcctgGAGGAGTGGCAGTGGCCTTCAGAGGTCTTGATTAGTGCTT
T
GAGGCCACTGTTTGTGGAACCTGGC|GTTTAATACTTGCATTTCCTCCAG
AAAGCATGTGGGCATTGGTAGTGGATGTGCTTTTAAAAAGAGCCTCTTAT
GG

2G-2H – chr2: 144313123-144313124
>hg38_dna range=chr2:144313048-144313199 5'pad=75 3'pad=75 strand=+
repeatMasking=lower
CTGCAGCTCATTAAACATCCAGACAAACTACATATCAATTTACACTTCTG
GCAGACAAATCCAAATCCTGGGCTGG|AAAAAAAAGGTCCATTTGGGGAGA
AACAGGGAAATGTCACAAAACTAACTCCCCACTAGATTAACAGCTCCATC
GG

>hg38_dna range=chr2:144313048-144313199 5'pad=75 3'pad=75 strand=-
repeatMasking=lower
CCGATGGAGCTGTTAATCTAGTGGGGAGTTAGTTTTGTGACATTTCCCTG
T
TTCTCCCCAAATGGACCTTTTTTTT|CCAGCCCAGGATTTGGATTTGTCT
GCCAGAAGTGTAAATTGATATGTAGTTTGTCTGGATGTTTAATGAGCTGC
AG

2H-2I – chr2: 144313180-144313181
>hg38_dna range=chr2:144313105-144313256 5'pad=75 3'pad=75 strand=+
repeatMasking=lower
AATCCAAATCCTGGGCTGGAAAAAAAAGGTCCATTTGGGGAGAAACAGGG
AAATGTCACAAAACTAACTCCCCACT|AGATTAACAGCTCCATCGGAATGA
CACTTTTAatctctcttctaatttctcctatatgcgggaaacaggtgttg
ag

>hg38_dna range=chr2:144313105-144313256 5'pad=75 3'pad=75 strand=-
repeatMasking=lower
ctcaacacctgtttcccgcatataggagaaattagaagagagatTAAAAG
TGTCATTCCGATGGAGCTGTTAATCT|AGTGGGGAGTTAGTTTTGTGACAT
TTCCCTGTTTCTCCCCAAATGGACCTTTTTTTTCCAGCCCAGGATTTGGA
TT

2I-2J – chr2: 144385912-144385913
>hg38_dna range=chr2:144385837-144385988 5'pad=75 3'pad=75 strand=+
repeatMasking=lower
TTATCTGATTGTGAAAAGTATCTAATACATTTATTCATATTTATTCAAAT
ATAGTCCCTTCTTTCCCCTTTCCTCC|AAGCCCTCCCACTCCGCCCCCAGT
CCAATTCATGCTAAGGAAGATGTATGTTTTGTTTAGCTCTTGCGGAGAAA
TT

>hg38_dna range=chr2:144385837-144385988 5'pad=75 3'pad=75 strand=-
repeatMasking=lower
AATTTCTCCGCAAGAGCTAAACAAAACATACATCTTCCTTAGCATGAATT
GGACTGGGGGCGGAGTGGGAGGGCTT|GGAGGAAAGGGGAAAGAAGGGACT
ATATTTGAATAAATATGAATAAATGTATTAGATACTTTTCACAATCAGAT
AA

2J-2K – chr2: 144411546-144411547
>hg38_dna range=chr2:144411471-144411622 5'pad=75 3'pad=75 strand=+
repeatMasking=lower
CATTAAAAGTGCATGTATGCGTTTTTGCAGAAGGTTTCCAAAAGACATAT
TGGCACTAGTTACCTAAAAACAACCG|TGAGGGAAGAGGATGATCTTTTTC
TCATTACACAGCTACATGGGCGGCCTCGCAGAGCCTCCAGCTGGTACAGA
GG

>hg38_dna range=chr2:144411471-144411622 5'pad=75 3'pad=75 strand=-
repeatMasking=lower
CCTCTGTACCAGCTGGAGGCTCTGCGAGGCCGCCCATGTAGCTGTGTAAT
GAGAAAAAGATCATCCTCTTCCCTCA|CGGTTGTTTTTAGGTAACTAGTGC
CAATATGTCTTTTGGAAACCTTCTGCAAAAACGCATACATGCACTTTTAA
TG

2K-2L – chr2: 145061847-145061848
>hg38_dna range=chr2:145061772-145061923 5'pad=75 3'pad=75 strand=+
repeatMasking=lower
ACATGTAGCTCAAGTCTTTCAATGGTACACAAGCAGAACCTGAAAGTGTA
T
GATCTCAGAGTTAAACATGAAAAGT|AGACAAGATACTTTGATAAGAAAT
AGATAAACCCAAAGACAGTCAATGCATTACCTGGTATTTGGGGGGTAGGT
AT

>hg38_dna range=chr2:145061772-145061923 5'pad=75 3'pad=75 strand=-
repeatMasking=lower
ATACCTACCCCCCAAATACCAGGTAATGCATTGACTGTCTTTGGGTTTAT
CTATTTCTTATCAAAGTATCTTGTCT|ACTTTTCATGTTTAACTCTGAGAT

CATACACTTTCAGGTTCTGCTTGTGTACCATTGAAAGACTTGAGCTACAT
GT

2L-2M – chr2: 145061923-145061924
>hg38_dna range=chr2:145061848-145061999 5'pad=75 3'pad=75 strand=+
repeatMasking=lower
AGACAAGATACTTTGATAAGAAATAGATAAACCCAAAGACAGTCAATGCA
TTACCTGGTATTTGGGGGGTAGGTAT|GGAGTTAAGGACAATACAAGGTAT
TACCTTCAGGTGGGTTCATCAGTTTTTTAGATAACGATTGAATACACATT
TA

>hg38_dna range=chr2:145061848-145061999 5'pad=75 3'pad=75 strand=-
repeatMasking=lower
TAAATGTGTATTCAATCGTTATCTAAAAAACTGATGAACCCACCTGAAGG
T
AATACCTTGTATTGTCCTTAACTCC|ATACCTACCCCCCAAATACCAGGT
AATGCATTGACTGTCTTTGGGTTTATCTATTTCTTATCAAAGTATCTTGT
CT

2M-2N – chr2: 145070083-145070084
>hg38_dna range=chr2:145070008-145070159 5'pad=75 3'pad=75 strand=+
repeatMasking=lower
AACCCAAAACACAAATGAAGATGCTGtcacacctctccacaaaagggaca
caaaaagtctcatcttcacttttcat|tctcaaagttgaaggtggtcaagt
gatacaatgtcctcttcatcaggtcttgatgcagtttctcagagtgcagt
ga

>hg38_dna range=chr2:145070008-145070159 5'pad=75 3'pad=75 strand=-
repeatMasking=lower
tcactgcactctgagaaactgcatcaagacctgatgaagaggacattgta
t
cacttgaccaccttcaactttgaga|atgaaaagtgaagatgagactttt
tgtgtcccttttgtggagaggtgtgaCAGCATCTTCATTTGTGTTTTGGG
TT

2N-2O – chr2: 145072714-145072715
>hg38_dna range=chr2:145072639-145072790 5'pad=75 3'pad=75 strand=+
repeatMasking=lower
agtttactgtgagccagagaaccaggctgggggctgtatcacttagaat
gagatccaaactcacaccatagttcc|atccttgaggttgccatgttgctg
ccacccctCCTGCCACCCCACCACTGAAGCTTTGGATGCTGGATTCTGGG
CA

>hg38_dna range=chr2:145072639-145072790 5'pad=75 3'pad=75 strand=-
repeatMasking=lower
TGCCCAGAATCCAGCATCCAAAGCTTCAGTGGTGGGGTGGCAGGaggggt
g
gcagcaacatggcaacctcaaggat|ggaactatggtgtgagtttggatc
tcattctaagtgatacagccccccagcctggttctctggctcacagtaaa
ct

2O-2P – chr2: 145074546-145074547
>hg38_dna range=chr2:145074471-145074622 5'pad=75 3'pad=75 strand=+
repeatMasking=lower
ATAGAGTTCAGAATGTGAGTGAAATGTAACTGAAAAGGAAAGCCTTGATT
TTACATATAAAATATCTTTACCAGAT|GGGTGAAGTAAGGTAACTTAAGAC
AATACTTAAAATTCAATAACAATGATGAAACTATGAAATACAAAACTTCT
GG

```
>hg38_dna range=chr2:145074471-145074622 5'pad=75 3'pad=75 strand=-
repeatMasking=lower
CCAGAAGTTTTGTATTTCATAGTTTCATCATTGTTATTGAATTTTAAGTA
TTGTCTTAAGTTACCTTACTTCACCC|ATCTGGTAAAGATATTTTATATGT
AAAATCAAGGCTTTCCTTTTCAGTTACATTTCACTCACATTCTGAACTCT
AT
```

2P-2Q – chr2: 145075679-145075680

```
>hg38_dna range=chr2:145075604-145075755 5'pad=75 3'pad=75 strand=+
repeatMasking=lower
GGTAGAATTGATTGCAAAGTTGCTTCATATTCTGCAATACTGTTGCTGAA
ACTATTTTAGGTATGTCCTTGGTATT|TCTCAAATATTAGAATTCTAATGT
AGTAAAATCAAATTTAATTTGCAACTGAATCATGCACAAACTAAACAGCA
GC
```

```
>hg38_dna range=chr2:145075604-145075755 5'pad=75 3'pad=75 strand=-
repeatMasking=lower
GCTGCTGTTTAGTTTGTGCATGATTCAGTTGCAAATTAAATTTGATTTTA
CTACATTAGAATTCTAATATTTGAGA|AATACCAAGGACATACCTAAAATA
GTTTCAGCAACAGTATTGCAGAATATGAAGCAACTTTGCAATCAATTCTA
CC
```

5A-5B – chr5: 92229394-92229395

```
>hg38_dna range=chr5:92229319-92229470 5'pad=75 3'pad=75 strand=+
repeatMasking=lower
AATTGTTTCAAAGAGGTCAAGGTTTGGACTTATGTGAAGTCAACATTTGA
AAACATTTGTAAATATTCTGTCAATC|ATTAGTATATGCTCTTTTTAGGTA
TTACTTCTCATAATTATCATAATTATCTTAATAATTTATTGATTATTTAT
AA
```

```
>hg38_dna range=chr5:92229319-92229470 5'pad=75 3'pad=75 strand=-
repeatMasking=lower
TTATAAATAATCAATAAATTATTAAGATAATTATGATAATTATGAGAAGT
AATACCTAAAAAGAGCATATACTAAT|GATTGACAGAATATTTACAAATGT
TTTCAAATGTTGACTTCACATAAGTCCAAACCTTGACCTCTTTGAAACAA
TT
```

5B-5C – chr5: 92229641-92229642

```
>hg38_dna range=chr5:92229566-92229717 5'pad=75 3'pad=75 strand=+
repeatMasking=lower
ACAATTACAGAGGCGTATCAAAAAGGCTTTTAATTGAAGAGTATTTACAA
TTAAATTATAACAGAAAGAATACTGT|AAAAGTAAACAAGTAAGTTCTTTT
TGAAAAACAGGAATGTGTAACATTCAAAAAGACCACAAAAATTAATTTCT
GT
```

```
>hg38_dna range=chr5:92229566-92229717 5'pad=75 3'pad=75 strand=-
repeatMasking=lower
ACAGAAATTAATTTTTGTGGTCTTTTTGAATGTTACACATTCCTGTTTTT
CAAAAAGAACTTACTTGTTTACTTTT|ACAGTATTCTTTCTGTTATAATTT
AATTGTAAATACTCTTCAATTAAAAGCCTTTTTGATACGCCTCTGTAATT
GT
```

5C-5D – chr5: 92481904-92481905

```
>hg38_dna range=chr5:92481829-92481980 5'pad=75 3'pad=75 strand=+
repeatMasking=lower
tggcataaaaacacataaacaaatggaacaagatagaaagcccagaaata
tacccacacatataaagtcaactaat|ctttgttaaaggtgccaggaatgc
```

acaatgacggaaggacattgtctttaataaatgaccttgggaaagctggc
ta

>hg38_dna range=chr5:92481829-92481980 5'pad=75 3'pad=75 strand=-
repeatMasking=lower
tagccagctttcccaaggtcatttattaaagacaatgtccttccgtcatt
gtgcattcctggcacctttaacaaag|attagttgactttatatgtgtggg
tatatttctgggctttctatcttgttccatttgtttatgtgtttttatgc
ca

5D-5E – chr5: 92489948-92489949
>hg38_dna range=chr5:92489873-92490024 5'pad=75 3'pad=75 strand=+
repeatMasking=lower
AGGGAAGAAAATTGATGTTTTGGTTTTGTAAAAACCTATGCTTTTGTATT
TCATCTGGGCTCTTACCTGCCTACTC|ACATTTTAAAAATGTACTTCATAT
TGACCCTTAGTACATTTTCCACAGGATTGTTCCAAGCTTTCTGCTCTCTC
AG

>hg38_dna range=chr5:92489873-92490024 5'pad=75 3'pad=75 strand=-
repeatMasking=lower
CTGAGAGAGCAGAAAGCTTGGAACAATCCTGTGGAAAATGTACTAAGGGT
CAATATGAAGTACATTTTTAAAATGT|GAGTAGGCAGGTAAGAGCCCAGAT
GAAATACAAAAGCATAGGTTTTTACAAAACCAAAACATCAATTTTCTTCC
CT

5E-5F – chr5: 92494785-92494786
>hg38_dna range=chr5:92494710-92494861 5'pad=75 3'pad=75 strand=+
repeatMasking=lower
aaatatgtggagcacatacacatctgagttgagcattgtgcttgggccaa
g
gatgtggaagtaagaggcagagtcc|ttgcctttatgtggaacacaatGA
CCGCGTTTTCTCCAAAAGACCTATGAACTGCTTAACACCAATCAATATTT
TA

>hg38_dna range=chr5:92494710-92494861 5'pad=75 3'pad=75 strand=-
repeatMasking=lower
TAAAATATTGATTGGTGTTAAGCAGTTCATAGGTCTTTTGGAGAAAACGC
GGTCattgtgttccacataaaggcaa|ggactctgcctcttacttccacat
ccttggcccaagcacaatgctcaactcagatgtgtatgtgctccacatat
tt

21A-21B – chr21: 23621186-23621187
>hg38_dna range=chr21:23621111-23621262 5'pad=75 3'pad=75 strand=+
repeatMasking=lower
TTAAAAGTATACTACTTTAGTGTGTGAAAAAGTATAAACGTGTATATATA
T
AAGAACATCTAtactttttttaacac|ttacctgtctgcaccactaatctt
taagtaccatgcaactgggtctgtttttgttcaccattgtatttccatct
cc

>hg38_dna range=chr21:23621111-23621262 5'pad=75 3'pad=75 strand=-
repeatMasking=lower
ggagatggaaatacaatggtgaacaaaaacagacccagttgcatggtact
taaagattagtggtgcagacaggtaa|gtgttaaaaaagtaTAGATGTTCT
TATATATATACACGTTTATACTTTTTCACACACTAAAGTAGTATACTTTT
AA

21B-21C – chr21: 23622264-23622265

>hg38_dna range=chr21:23622189-23622340 5'pad=75 3'pad=75 strand=+ repeatMasking=lower
gactgacccctgtggagtctcctgaggcttctcaattatctgacctctag
t
aattgcCATAACAATATATAATATC|AGCCCTTTTACTTGTGAAATGTAA
ATGCTTGTGTGTCTATAAGAATTAAATACAAATAAAATTGATACAGTTTT
TA

>hg38_dna range=chr21:23622189-23622340 5'pad=75 3'pad=75 strand=- repeatMasking=lower
TAAAAACTGTATCAATTTTATTTGTATTTAATTCTTATAGACACACAAGC
ATTTACATTTCACAAGTAAAAGGGCT|GATATTATATATTGTTATGgcaat
tactagaggtcagataattgagaagcctcaggagactccacaggggtcag
tc

### Variants

## 2A-2O

-> <-

Expected:
CAATCAAGCAACCATCATTTGCTAATAAATGTTTTTATCACAGCTTTCAA
ACCTTTTTCAAGACATCTGAGCTCAT|ATCTGGTAAAGATATTTTATATGT
AAAATCAAGGCTTTCCTTTTCAGTTACATTTCACTCACATTCTGAACTCT
AT

LRS:
CAATCAAGCAACCATCATTTGCTAATAAATGTTTTTATCACAGCTTTCAA
ACCTTTTTCAAGACATCTGAGCTCAT|ATCTGGTAAAGATATTTTATATGT
AAAATCAAGGCTTTCCTTTTCAGTTACATTTCACTCACATTCTGAACTCT
AT

Microhomology (50bp):
2A-2B CCTTTTTCAAGACATCTGAGCTCATCTTCACTTCCTCCTTCTTGAAAAAG
LRS CCTTTTTCAAGACATCTGAGCTCATATCTGGTAAAGATATTTTATATGTA
2O-2P_reverse TGTCTTAAGTTACCTTACTTCACCCATCTGGTAAAGATATTTTATATGTA

* ** ** * * * * * * ** *

## 2O-21C

<- ->

Expected:
TGCCCAGAATCCAGCATCCAAAGCTTCAGTGGTGGGGTGGCAGGaggggt
ggcagcaacatggcaacctcaaggat|AGCCCTTTTACTTGTGAAATGTAA
ATGCTTGTGTGTCTATAAGAATTAAATACAAATAAAATTGATACAGTTTT
TA

LRS:
GCCCAGAATCCAGCATCCAAAGCTTCAGTGTGGTGGGTGGCAGGAGGGGT
GGCAGCAACATGGCAACCTCAAGGAT|AGCCCTTAATACTTGTGAAATGTA
AATGCTTGTGTGTCTATAAGAATTAAATACAAATAAAATTGATACAGTTT
TT

Microhomology (50bp):
2N-2O_reverse gcagcaacatggcaacctcaaggatggaactatggtgtgagtttggatct-

```
LRS                GCAGCAACATGGCAACCTCAAGGATAGCCCTTAATACTTGTGAAATGTAA-
21B-21C            -aattgcCATAACAATATATAATATCAGCCCTTTTACTTGTGAAATGTAAA
                     *    ***  ***  *  *  **   *       *         *
```

## 5A-2C

-> <-

Expected:

AATTGTTTCAAAGAGGTCAAGGTTTGGACTTATGTGAAGTCAACATTTGA
AAACATTTGTAAATATTCTGTCAATC|GGCTCATATCTCACCATCCAGTGC
CCCTCCCTCCTCCTTTATTTGTCAAAGCTCAACCCATCTTCCAAGAGTCA
TC

LRS:

TGTTTCAAAGAGGTCAAGGTTTGGACTTCTATGTGAAGTCAACATTTGGA
AAACATTTGTAAATATTCTGTCAATC|CACCTCTGATATAGCCTAACTCCC
ACCTCTGGCTCATATCTCACCATCCAGTGCCCTCCTCCTTTACTTGTCTC
AA

Microhomology (50bp):

```
5A-5B              AACATTTGTAAATATTCTGTCAATCATTAGTATATGCTCTTTTTAGGTAT-
LRS                AACATTTGTAAATATTCTGTCAATCCACCTCTGATATAGCCTAACTCCCA-
2C-2D_reverse      -AGATTAGTCAGTGTTCCCTGAAGATGGCTCATATCTCACCATCCAGTGCC
                    * *** ** * * ***  * **         **
```

Microhomology (150bp):

```
5A-5B              AATTGTTTCAAAGAGGTCAAGGTTTGGACTTATGTGAA--GTCAACAT-TTGAAAACATT
LRS                ---TGTTTCAAAGAGGTCAAGGTTTGGACTTCTATGTGAAGTCAACATTTGGAAAACATT
2C-2D_reverse      CCTTCCTAGCCTTACTTCAATCCTTGAACATTATA----TCCTGGACTGTAGCCTAGATT
                      *  *      *  ****   *** ** *                * * *   * ***

5A-5B              TGTAAATATTCTGTCAATCATTAGTATATGCTCTTTTTAGGTATTACTTCTCATAATTAT
LRS                TGTAAATATTCTGTCAATCCACCTCTGATATAGCCTAACTCCCACCTCTGGCTCATATCT
2C-2D_reverse      AGTCAGTGTTCCCTGAAG-----------------------------ATGGCTCATATCT
                    ** * * ***  * **                               *  *  *  * *

5A-5B              CATAATTATCTTAAT-----AATTTATTGATTATTTATAA--------------------
LRS                CACCATCCAGTGCCC-----TCCTCCTTTACTTGTCTCAA--------------------
2C-2D_reverse      CACCATCCAGTGCCCCTCCCTCCTCCTTTATTTGTCAAAGCTCAACCCATCTTCCAAGAG
                   **  **    *            *  ** * *  *   *

5A-5B              -----
LRS                -----
2C-2D_reverse      TCATC
```

Blasting of LRS sequence (150bp) yielded no significant results associated for black insertion sequence.
(significant defined as in vicinity of previously identified breakpoints)

## 2C-5E

<- <-

Expected:

CTTGCTTTGCAAATCAGGGAATCTCAGCAACTGGGCCTTCAATTCCAGGC
AGCCATAGTTTTAAATCCAGAGGTGG|ggactctgcctcttacttccacat
ccttggcccaagcacaatgctcaactcagatgtgtatgtgctccacatat
tt

LRS:

CCTTGCTTTGCAAATCAGGGAATCTCAGCAACTGGGCCTTCAATTCCAGG
CAGCCATAGTTTTAAATCAGAGGTGG|GGAGTTAGGCCTCTGCCTCTTGGC

TACATCCTTGGCCCAAGCACAATGCTCAACTCAGATGTGTATGTGCTCCA
CA

Microhomology (50bp):

```
2B-2C_reverse      -GCCATAGTTTTAAATCCAGAGGTGGTTTTAGAGTGTCACACCATGGGGTC
LRS                AGCCATAGTTTTAAATCAGAGGTGGGGAGTTAGGCCTCTGCCTCTTGGCT-
5E-5F_reverse      -GTCattgtgttccacataaaggcaaggactctgcctcttacttccacatc
                    * *** ** **  *      *           *  **   *       *
```

Microhomology (150bp):

```
2B-2C_reverse      -CTTGCTTTGCAAATCAGGGAATCTCAGCAACTGG-----GCCTTCAATTCCAGGCAGCC
LRS                CCTTGCTTTGCAAATCAGGGAATCTCAGCAACTGG-----GCCTTCAATTCCAGGCAGCC
5E-5F_reverse      ------TAAAATATTGATTGGTGTTAAGCAGTTCATAGGTCTTTTGGAGAAAACGCGGTC
                         *     * * *  *    * ****  *          **  *    * ** * *

2B-2C_reverse      ATAGTTTTAAATCCAGAGGTGGTTTTAGAGTGTCACACCATGGGGTCTCTTTTTCAAGAA
LRS                ATAGTTTTAAATCAGAGGTGGGGAGTTAGGCCTCTGCCTCTTGGCTACATCCTTGGCCCA
5E-5F_reverse      attgtgttccacataaaggcaaggactc----tgcctcttacttccacatccttggccca
                   ** ** **  *      *              *    *           *  **     *

2B-2C_reverse      GGAGGAAGTGAAGATGAGCTCAGATGTCTTGAAAAAGG--------
LRS                AGCACAAT----GCTCAACTCAGATGTGTATGTGCTCCACA-----
5E-5F_reverse      agcaca----atgctcaactcagatgtgtatgtgctccacatattt
                    *   *      * * * ********* *
```

Blasting of LRS sequence (150bp) yielded a 92% identity to chr5:92494786-92494715 of the underlined sequence + blue sequence

```
Query  7         AGGCCTCTGCCTCTTGGCT--ACATCCTTGGCCCAAGCACAATGCTCAACTCAGATGTGT  64
                 ||| |||||||||||   |  |||||||||||||||||||||||||||||||||||||
Sbjct  92494786  AGGACTCTGCCTCTTACTTCCACATCCTTGGCCCAAGCACAATGCTCAACTCAGATGTGT
92494727

Query  65        ATGTGCTCCACA  76
                 ||||||||||||
Sbjct  92494726  ATGTGCTCCACA  92494715
```

# 5E-2I

<- ->

Expected:
CTGAGAGAGCAGAAAGCTTGGAACAATCCTGTGGAAAATGTACTAAGGGT
CAATATGAAGTACATTTTTAAAATGT|AGATTAACAGCTCCATCGGAATGA
CACTTTTAatctctcttctaatttctcctatatgcgggaaacaggtgttg
ag

LRS:
GCCTAGATTAGTAGCCTAACTCCATACCTACCCCCCAAATACCAGGTAAT
GCATTGACTGTCTTCAGTGTTCCTGA|AAGATTAACAGCTCCATCGGAATG
ACACTTTTTTAATCTCTCTTCTAAATTTCTCTATATGCAGGGAAACAGGT
GT

Microhomology (50bp):

```
5D-5E_reverse      AATAT-GAAGTACATTTTTAAAATGTGAGTAGGCAGGTAAGAGCCCAGATG-
LRS                ATTGACTGTCTTCAGTGTTCCCTGAAGATTAAGCAGCTCCCATCGGAATG--
2H-2I              AATGTCACAAAACTAACTCCCCACTAGATTA-AC-AGCTCCATCGGAATGAC
                   * *         *    *        ** **  *       * *  *
```

Microhomology (150bp):

```
5D-5E_reverse      CTGAGAGAGCAGAAAGCTTGGAACAATCCTGTGGAAAATGTACTAAGGGTCAATATGAAG
LRS                --CCTAGATTAGTAGCC-----TAACTCCATA-CC-TACCCCCCA-------AATACCAG
```

```
2H-2I              -------------AATC-----CAAATCCTGG-GC-TGGAAAAAAAAGGTCCATTTGGGG
                                *  *       * ***               *       *      *

5D-5E_reverse      TACATTTTTAAAATGTGAGTAGGCAGGTAAGAGCCCAGATGAAATACAAAAGCATAGGTT
LRS                GTAATGCATTGACTGTCT--TCAGTGTTC---CCTGAAGATTAAGCAGCTCCCATCG-GA
2H-2I              AGAAACAGGGAAATGTCACAAAACTAACT---CCCCACTAGATTAACAGCTCCATCG-GA
                      *       * ***                 *  *               *** *

5D-5E_reverse      TTTACAAAACC-----AAAACATCAATTTTCTTCCCT---------------------
LRS                ATGACACTTTTAATCTCTCTCTTCTAATTTCTCCTATATGCGGGAAACAGGTGT----
2H-2I              ATGACACTTTTA--atctctcttctaatttctcctatatgcgggaaacaggtgttgag
                    * ***              * ** * ***** *  *
```

Microhomology (300bp):

```
5D-5E_reverse      TGAGTTCCAGAATCTCAATCTTGAGAAGCAGAAAATAAGATCAAGTGATAAATGTTTTGG
LRS                ------------------------------------------------------------
2H-2I              ------------------------------------------------------------


5D-5E_reverse      AATAAAATATGACCTCTGAGAGAGCAGAAAGCTTGGAACAATCCTGTGGAAAATGTACTA
LRS                ---------------------------CAAGCTTGGAACAATCCTGTGGAAAATGTACTA
2H-2I              ------------------------GCACAAGGAAACAAGATTCTGCAGCTCATTAAACAT
                                               ***      ** * **     *    * *   **

5D-5E_reverse      AGGGTCAATATGAAGTACATTTTTAAAATGTGAGTAGGCAGGTAA---------------
LRS                AGGGTCAATATGAAGTACATTTTAAATGTATCCTGGACTGTAGCCTAGATTAGTAGCCTA
2H-2I              CCAGACAAACTACATATCAATTTACACTTCTGGCAGACAAATCCAAA---TCCTGGGCTG
                      * ***  *  *   ** ***  *  * *

5D-5E_reverse      --------------------------------------------------GAGCCCAGAT
LRS                ACTCCATACCTACCCCCCAAATACCAGGTAATGCATTGACTGTCTTCAGTGTTCCTGAAA
2H-2I              GAAAAAA-AAGGTCCATTTGGGGAGAAACAGGGAAATGTCACAAAACTAACTCCCCACTA
                                                                        **

5D-5E_reverse      GAAATACAAAAGCATAGGTTTTTAC----AAAACCAAAACATCAATTTTCTTCCCTTTTC
LRS                GATTAACAGCTCCATCGGAATGACACTTTTTTAATCTCTCTTCTAAATTTCTCTATATGC
2H-2I              GATTAACAGCTCCATCGGAATGACACTTTTAatc--tctcttctaatt-tctcctatatg
                   **   ***    *** **  *                  * ** *  *   **

5D-5E_reverse      CTGGAAAGATGTAATTGATAGTGCA-------TACTACCTGATGCTCGGCTTGACATGGA
LRS                AGGGAAACAG-GTGTTGAGGGTGAACCAAGTCTATTCTCTTTTACTAAGCC-CACATCCC
2H-2I              cgggaaacag-gtgttgagggtgaaccaagtctattctcttttactaagccccacatccc
                     ***** *     ****  *** *       ** *  **  * **  **   ****

5D-5E_reverse      GTATGATAAAGTATAATT----------------
LRS                TCTTTAGAAAGCAGGGTTACTTATTACCCTT---
2H-2I              tctttagaaagcagggttacttattaccctttgg
                      * * **** *   **
```

Blasting of LRS sequence (300bp) yielded a 100% identity to chr2:145061930-145061883 of the second underlined sequence. No further results for other black insertion sequences.
Blasting of black insertion sequence alone yielded no results, except for the one reported above.
Blasting of insertion sequence ATCCTGGACTGTAGCCTAGATTAGTAGCC yielded a 100% identity to chr2:143960430-143960406:

```
Query  1          ATCCTGGACTGTAGCCTAGATTAGT  25
                  |||||||||||||||||||||||||
Sbjct  143960430  ATCCTGGACTGTAGCCTAGATTAGT  143960406
```

# 2I-5C

-> <-

Expected:

```
TTATCTGATTGTGAAAAGTATCTAATACATTTATTCATATTTATTCAAAT
ATAGTCCCTTCTTTCCCCTTTCCTCC|attagttgactttatatgtgtggg
tatatttctgggctttctatcttgttccatttgtttatgtgtttttatgc
ca
```

LRS:

```
TTATCTGATTGTGAAAAGTATCTAATACATTATTCATATTTATTCAAATA
TAGTCCCTTCTTTCCCCTTTCCTCCA|TTTGGGGAGAAACAGGGAAATGTC
ACAAAACTAGTATTGACATCTAGTAGCCTAGATTAGTCAGTGTAGCCTAA
AT
```

Microhomology (50bp):

```
2I-2J              TAGTCCCTTCTTTCCCCTTTCCTCCAAGCCCTCCCACTCCGCCCCCAGTC---
LRS                -AGTCCCTTCTTTCCCCTTTCCTCCATTTGGGGAGAAACAGGGAAATGTCA--
5C-5D_reverse      ---tgcattcctggcacctttaacaaagattagttgactttatatgtgtgggt
                      * * *** *  * * **   * *                     **
```

Microhomology (150bp):

```
2I-2J              TTATCTGATTGTGAAAAGTATCTAATACATTTATTCATATTTATTCAAATATAGTCCCTT
LRS                TTATCTGATTGTGAAAAGTATCTAATACA-TTATTCATATTTATTCAAATATAGTCCCTT
5C-5D_reverse      --------tagccagctttcccaaggtcatt------tattaaagacaatgtccttccgt
                           * *  *    *  * *   ** *      **** *    *** *  * ** *

2I-2J              CTTTCCCCTTTCCTCCAAGCCCTCCCACTCCGCCCCCAGTCCAATTCATGC-TAAGG-AA
LRS                CTTTCCCCTTTCCTCCATTTGGGGAGAAACAGGGAAATGTCACAAAACTAG-TATTG-AC
5C-5D_reverse      cattgtgcattcctggcacctttta----acaaagattagttgactttatatgtgtgggta
                   * **   * *****               *        **        *   *   *

2I-2J              GATGTAT-GTTTTGTT------------TAGCTCTTGCGGAGAAATT---
LRS                ATCTAGTAGCCTAGAT------------TAGTCAGTGTAGCCTAAAT---
5C-5D_reverse      tatttctgggctttctatcttgttccatttgtttatgtgtttttatgcca
                         * *  *   *            * *    **       *
```

Microhomology (300bp):

```
2I-2J              -CTCAAATTTGTGACTAAGATTTGCTTTTATTAAGGCTTACATTATTAGAAAGATAAAAA
LRS                CCTCAAATTTGTGACTAAGATTTGCTTTTATTAAGGCTTACATTATTAGAAAGATAAAAA
5C-5D_reverse      --------------gtccatttgagttgatttttgtgtgtaggataaggaatacaataa
                                  *    ****  ** ***   *  *  *  ** ** ** * ** **

2I-2J              ATGACCTTTTTAAAAGTTATCTGATTGTGAAAAGTATC-TAATACATTTATTCATATTTA
LRS                ATGACCTTTTTA-AAGTTATCTGATTGTGAAAAGTATC-TAATACAT-TATTCATATTTA
5C-5D_reverse      gagtccaatcttagtgtt--tttcatgtgaatagccagctttcccaaggtcatttattaa
                     * **  * *    ***   *   ****** **     *    **        **** *

2I-2J              TTCAAATATAGTCCCTTCTTTCCCCTTTCCTCCAAGCCCTCCCACTCCGCCCCCAGTCCA
LRS                TTCAAATATAGTCCCTTCTTTCCCCTTTCCTCCATTTGGGGAGAAACAGGGAAATGTCAC
5C-5D_reverse      agacaatgtccttccgtcattgtgcattcctggca-------------------------
                       *** *  * ** ** **   * *****

2I-2J              ATTCATGCTAAGGAAGATGTAT-GTTTTGTTTAGCTCTTGCGGAGAAATTCTGATACGCA
LRS                AAAACTAGTATTGACATCTAGTAGCCTAGATTAGTCAGTGTAGCCTAAATTAGTTGACTT
5C-5D_reverse      ------------------------------------cctttaacaaagattagttgactt
                                                         *       *  *  * *

2I-2J              TCTCTCTTCCCAGTTATTTCAGGCCTAAGCTTACAGTGTCAT-GT---------------
LRS                TATATGTGTGGGTATATTTCTGGGCTTTCTATCTTGTTTCCATTT---------------
5C-5D_reverse      tatatgtgtgggtatatttctgggctttctatcttgttccatttgtttatgtgtttttat
                   * * * *       ****** ** **     *   **  *

2I-2J              -----------------------------------GAAGAAAAAAGAAAAGATTGC----
LRS                -----------------------------------GTTTATGTGTT--TTTATACC----
5C-5D_reverse      gccattttcaacactgtttcaataactatagttttgtatcaataatataattttgtaatc
                                                      *                *
```

```
2I-2J              -------------------
LRS                -------------------
5C-5D_reverse      aggaagtatgatgtttcca
```

Blasting of LRS sequence (300bp) yielded a 100% identity to chr2:144313134-144313170 of the first underlined sequence. No further results for other black insertion sequences.
Blasting of the black insertion sequence yielded an additional match for the second underlined sequence with a 100% identity match to chr2:143960420-143960400.

# 5C-2K

<- ->

Expected:

ACAGAAATTAATTTTTGTGGTCTTTTTGAATGTTACACATTCCTGTTTTT
CAAAAAGAACTTACTTGTTTACTTTT|TGAGGGAAGAGGATGATCTTTTTC
TCATTACACAGCTACATGGGCGGCCTCGCAGAGCCTCCAGCTGGTACAGA
GG

LRS:

TTCCTGTTTTTCAAAAAAGAACTTTACTTTGTTTACTTTTTTAACACTTA
CCTGTCTGCACCACTAGTTAGTAGCC|TGAGGGAAGAGGATGATCTTTTTC
TCATTACACAGCTACATGGGCGTTCCCGCAGAGCCTCCAGCTGGTACAGA
GG

Microhomology (50bp):

```
5B-5C_reverse      -------AAAAAGAACTTACTTGTTTACTTTTACAGTATTCTTTCTGTTATAATTTA
LRS                CTGTCTGCACCACTAGTTAGTAGCCTGAGGGAAGAGGATGATCTTTTTCT-------
2J-2K              GGCACTAGTTACCTAAAAACAACCGTGAGGGAAGAGGATGATCTTTTTCT-------
                                 *   *      *      * ** **  * * * *
```

Microhomology (150bp):

```
5B-5C_reverse      ACAGAAATTAATTTTTGTGGTCTTTTTGAATGTTACACATTCCTGTTTTTCAAAAAGAAC
LRS                ----------------------------------------TTCCTGTTTTCAAAAAAGAA
2J-2K              -----------------------------------------------CATTAAAGTGCAT
                                                                   ** ***    *

5B-5C_reverse      TTACTTGTTTACTTTTACAGTATTCT-------TTCTG-TTATAATTTAATTGTAAATAC
LRS                CTTTACTTTGTT---TA----CTTTTTTAACACTTACCTGTCTGCACCACTAGTTAGTAG
2J-2K              GTATGCGTTTTTGCAGAAGGTTTCCAAAAGACATATTGGCACTAGTTACCTAAAAACAAC
                    *     **       *     *          *        *       *    *  *

5B-5C_reverse      TCTT----------------------CAATTAAAAGCCTTTTTGATACGCCTCTGTAATT
LRS                CCTGAGGGAAGAGGATGATCTTTTTCTCATTACACAGCTACATGGGCGTTCCCGCAGAGC
2J-2K              CGTGAGGGAAGAGGATGATCTTTTTCTCATTACACAGCTACATGGGCGGCCTCGCAGAGC
                     *                         **** *   **   **      * *    *

5B-5C_reverse      --------GT--------
LRS                CTCCAGCTGGTACAGAGG
2J-2K              CTCCAGCTGGTACAGAGG
                           *
```

Blasting of LRS sequence (150bp) yielded a 100% identity match of underlined sequence to chr21:23621173-23621205.

# 2K-2M

-> <-

Expected:

ACATGTAGCTCAAGTCTTTCAATGGTACACAAGCAGAACCTGAAAGTGTA
TGATCTCAGAGTTAAACATGAAAAGT|atgaaaagtgaagatgagactttt

tgtgtcccttttgtggagaggtgtgaCAGCATCTTCATTTGTGTTTTGGG
TT

LRS:
ACATGTAGCTCAAGTCTTTCAATGGTACACAAGCAGAACCTGAAAGTGTA
TGATCTCAGAGTTAAACATGAAAAGT|GAAGATGAGACTTTTTGTGTCCCT
TTTGTGGAGAGGTGTGACAGCATCTTCATTTGTGTTTTGGGTTGCATTTT
TA

Microhomology:
```
2K-2L              ---------GATCTCAGAGTTAAACATGAAAAGTAGACAAGATACTTTGATAAGAAATA
LRS                ---------GATCTCAGAGTTAAACATGAAAAGTGAAGATGAGACTTTTTGTGTCCCTT
2M-2N_reverse      cacttgaccaccttcaactttgagaatgaaaagtgaagatgagacttttt---------
                              ***   ** *  *********  * * ** *****
```

## 2M-2G

<-  <-

Expected:
TAAATGTGTATTCAATCGTTATCTAAAAAACTGATGAACCCACCTGAAGG
TAATACCTTGTATTGTCCTTAACTCC|CCAGCCCAGGATTTGGATTTGTCT
GCCAGAAGTGTAAATTGATATGTAGTTTGTCTGGATGTTTAATGAGCTGC
AG

LRS:
AAATGTGTATTCAATGCGTTATCTAAAAAACTGATGAACCCACCTGAAGG
TAATACCTTGTATTGTCCTTAACTCC|CAGCCCAGGATTTGGATTTGTCTT
AAATCCCAGCCCAGGATTTGGATTTGTCTGCCAAGTGTAGAATTGATATG
TA

Microhomology:
```
2L-2M_reverse      -AATACCTTGTATTGTCCTTAACTCCATACCTACCCCCCAAATACCAGGTA
LRS                -AATACCTTGTATTGTCCTTAACTCCCAGCCCAGGATTTGGATTTGTCTTA
2G-2H_reverse      TTCTCCCCAAATGGACCTTTTTTTTCCAGCCCAGGATTTGGATTTGTCTG-
                      * **         * **   * *   ** *       **
```

## 2G-5F

<-  ->

Expected:
atgcctcctgGAGGAGTGGCAGTGGCCTTCAGAGGTCTTGATTAGTGCTT
TGAGGCCACTGTTTGTGGAACCTGGC|ttgcctttatgtggaacacaatGA
CCGCGTTTTCTCCAAAAGACCTATGAACTGCTTAACACCAATCAATATTT
TA

LRS:
CATGCCTCCTGGAGGAGTGGCAGGGTTTCTCAGAGGTCTTGATTAGTGCT
TTGAGGCCACTGTTTGTGGAACCTGG|CTTGCCTTTATGTGGAACACAATG
ACCGCGTTTTCTCCAAAAGACCTATGAACTGCTTAACACCAATCAATATT
TT

Microhomology:
```
2F-2G_reverse      -GAGGCCACTGTTTGTGGAACCTGGCGTTTAATACTTGCATTTCCTCCAGA
LRS                TGAGGCCACTGTTTGTGGAACCTGGCTTGCCTTTATGTGGAACACAATGA-
5E-5F              -gatgtggaagtaagaggcagagtccttgcctttatgtggaacacaatGAC
                    ** *     **  * ** *     * *    *  *       *
```

## 21A-2E

-> ->

Expected:

TTAAAAGTATACTACTTTAGTGTGTGAAAAAGTATAAACGTGTATATATA
TAAGAACATCTAtactttttttaacac|TCCCTCAGTGATGTTTTCATATTT
ATAATTTTTGAGAACAATTTCTTGAGAGATGTTTTGGGAGCTGTATTCCA
GA

LRS:

TAAAAGTATACTACTTTAGTGTGTGTGAAAAAGAGTAAACGTGTATATAT
A
TAAGAACATCTATACTTTTTTAACAC|AAAACATCCCTCAGTGATGTTTTC
ATATTTATAATTTTTGAGAACAATTTCTTGAGAGATGTTTTGGGAGCTGT
AT

Microhomology:

```
21A-21B      -AAGAACATCTAtactttttttaacacttacctgtctgcaccactaatcttt-----
LRS          TAAGAACATCTATACTTTTTTAACACAAAACATCCCTCAGTGATGTTTTCA------
2D-2E        ----AGCT--TATGAAAATGACAGAGCATTTTCCCTCAGTGATGTTTTCATATTTA
                 * *   ***     *            *  **     *    *
```

# 2E-2Q

-> ->

Expected:

TAGCATTTAATCTGACCATCTGTTTACCAAAGGCTAGTACATTGAAAAAA
AGAAAGAAAATGGGTTTTTTTCCTAG|TCTCAAATATTAGAATTCTAATGT
AGTAAAATCAAATTTAATTTGCAACTGAATCATGCACAAACTAAACAGCA
GC

LRS:

CATTTAATCTGACCATCTGTTTACCAAAGGAAGCTAGTACATTGAAAAAA
AGAAAGAAAATGGGTTTTTTCCTAGC|TACAGTCTCAAATATTAGAATTCT
AATGTAGTAAAATCAAATTTAATTTGCAACTGAATCATGCACAAACTAAA
CA

Microhomology:

```
2E-2F      GAAAGAAAATGGGTTTTTTTCCTAGCCAATTGCAATGCCAATGTTTTAAT------
LRS        GAAAGAAAATGGGTTTTTT-CCTAGCTACAGTCTCAAATATTAGAATTCTA-----
2P-2Q      ----CTATTTTAGG-TATGTCCTTG-GTATTTCTCAAATATTAGAATTCTAATGTA
                 *  *  *  * *  *** *       *      * *    *  *
```

### Individual S4

#### Reference sequences

3A-3B – chr3: 50011089-50011090

```
>hg38_dna range=chr3:50011014-50011165 5'pad=75 3'pad=75 strand=+
repeatMasking=lower
CCCAAAAAGATTGTCTATCAGTTTGTCAGGAAGTTAGAGTAAAATGGTCT
T
AAAATGCATCAAGAGGgctgggcac|agtggctgatgcctgtagtttcag
ctactcaggaggctgagataggaggatcacttgagcccaggaattcgagt
ga
```

```
>hg38_dna range=chr3:50011014-50011165 5'pad=75 3'pad=75 strand=-
repeatMasking=lower
tcactcgaattcctgggctcaagtgatcctcctatctcagcctcctgagt
agctgaaactacaggcatcagccact|gtgcccagcCCTCTTGATGCATTT
TAAGACCATTTTACTCTAACTTCCTGACAAACTGATAGACAATCTTTTTG
GG
```

3B-3C – chr3: 50097202-50097203

```
>hg38_dna range=chr3:50097127-50097278 5'pad=75 3'pad=75 strand=+
repeatMasking=lower
ggctcacgcctgtaatcccagcactttaggattctgaggcaggcggatca
caaggtcaggagatcgagaccatcct|ggctagcacggtgaaaccctgtct
ctactaaaactacaaaaaaattagccagccgtggtggcgggcgcctgtag
tc
```

```
>hg38_dna range=chr3:50097127-50097278 5'pad=75 3'pad=75 strand=-
repeatMasking=lower
gactacaggcgcccgccaccacggctggctaatttttttgtagttttagt
agagacagggtttcaccgtgctagcc|aggatggtctcgatctcctgacct
tgtgatccgcctgcctcagaatcctaaagtgctgggattacaggcgtgag
cc
```

3C-3D – chr3: 52741363-52741364

```
>hg38_dna range=chr3:52741288-52741439 5'pad=75 3'pad=75 strand=+
repeatMasking=lower
CACAAACCATTTATAATTTTATCAGTGGGAATACTAGAGATCTAAATACT
ACTGAAAATGCACATTTAATATTCAT|TAAAACAGAAATAGTTTATAAAGG
GAGACAGAAAAACAAGAACTTACCCTTCCCCGTAATCCCCATCTGACTTA
TC
```

```
>hg38_dna range=chr3:52741288-52741439 5'pad=75 3'pad=75 strand=-
repeatMasking=lower
GATAAGTCAGATGGGGATTACGGGGAAGGGTAAGTTCTTGTTTTTCTGTC
TCCCTTTATAAACTATTTCTGTTTTA|ATGAATATTAAATGTGCATTTTCA
GTAGTATTTAGATCTCTAGTATTCCCACTGATAAAATTATAAATGGTTTG
TG
```

3D-3E – chr3: 52741372-52741373

```
>hg38_dna range=chr3:52741297-52741448 5'pad=75 3'pad=75 strand=+
repeatMasking=lower
TTTATAATTTTATCAGTGGGAATACTAGAGATCTAAATACTACTGAAAAT
```

GCACATTTAATATTCATTAAAACAGA|AATAGTTTATAAAGGGAGACAGAA
AAACAAGAACTTACCCTTCCCCGTAATCCCCATCTGACTTATCAGTTGAA
CT

>hg38_dna range=chr3:52741297-52741448 5'pad=75 3'pad=75 strand=-
repeatMasking=lower
AGTTCAACTGATAAGTCAGATGGGGATTACGGGGAAGGGTAAGTTCTTGT
TTTTCTGTCTCCCTTTATAAACTATT|TCTGTTTTAATGAATATTAAATGT
GCATTTTCAGTAGTATTTAGATCTCTAGTATTCCCACTGATAAAATTATA
AA

3E-3F – chr3: 52741456-52741457
>hg38_dna range=chr3:52741381-52741532 5'pad=75 3'pad=75 strand=+
repeatMasking=lower
ATAAAGGGAGACAGAAAAACAAGAACTTACCCTTCCCCGTAATCCCCATC
TGACTTATCAGTTGAACTTGTAGAAG|AACTTAACTCATCCTCAGACAGAC
AATGAATCTGTTTCCTTTCCTATTAAATGTTTGGAAGAATTAAAATTCTA
CA

>hg38_dna range=chr3:52741381-52741532 5'pad=75 3'pad=75 strand=-
repeatMasking=lower
TGTAGAATTTTAATTCTTCCAAACATTTAATAGGAAAGGAAACAGATTCA
TTGTCTGTCTGAGGATGAGTTAAGTT|CTTCTACAAGTTCAACTGATAAGT
CAGATGGGGATTACGGGGAAGGGTAAGTTCTTGTTTTTCTGTCTCCCTTT
AT

3F-3G – chr3: 52753018-52753019
>hg38_dna range=chr3:52752943-52753094 5'pad=75 3'pad=75 strand=+
repeatMasking=lower
cacacacacacaatggaatatttctaatctataaaaaggaatgaagttct
gatacatgctaaaacacgaatgaacc|ctgaaaacatgctaagtaaaataa
gccaggccaagtgtggtggctcacacctatcatcccagcacttgagctca
gg

>hg38_dna range=chr3:52752943-52753094 5'pad=75 3'pad=75 strand=-
repeatMasking=lower
cctgagctcaagtgctgggatgataggtgtgagccaccacacttggcctg
gcttattttacttagcatgttttcag|ggttcattcgtgttttagcatgta
tcagaacttcattccttttttatagattagaaatattccattgtgtgtgtg
tg

### Variants

## 3D-3F

-> <-

Expected:
TTTATAATTTTATCAGTGGGAATACTAGAGATCTAAATACTACTGAAAAT
GCACATTTAATATTCATTAAAACAGA|ggttcattcgtgttttagcatgta
tcagaacttcattccttttttatagattagaaatattccattgtgtgtgtg
tg

LRS:
TTTATAATTTTATCAGTGGGAATACTAGAGATCTAAATACTACTGAAAAT

```
GCACATTTAATATTCATTAAAACAGA|CGGTTCATTCGTGTTTTTAGCATG
TATCAGAACTTCATTCCTTTATAGATTAGAAATATTCCATTGTGTGTGTG
TG
```

Microhomology:

```
3D-3E              CACA-TTTAATATTCATTAAAACAGAAATAGTTTATAAAGGGAGACAGAAA--
LRS                CACA-TTTAATATTCATTAAAACAGACGGTTCATTCGTGTTTTTAGCATGT--
3F-3G_reverse      cttattttacttagcatgttttcag--ggttcattcgt-gttttagcatgtat
                   *  * **** *   ***     ***        *          *
```

# 3C-3E

```
<- <-
gactacaggcgcccgccaccacggctggctaatttttttgtagttttagt
agagacagggtttcaccgtgctagcc|CTTCTACAAGTTCAACTGATAAGT
CAGATGGGGATTACGGGGAAGGGTAAGTTCTTGTTTTTCTGTCTCCCTTT
AT
```

LRS:

```
GGACTACAGGCGCCCGCCACCACGGCTGGCTAATTTTTTGTAGTTTTAGT
AGAGACAGGGTTTCACCGTGTTAGCC|TGGCCTTTGTTTGTCTGAAAGTCA
AAGAACCTTCTACAAGTTCAACTGATAAGTCAGATGGGATTACGGGGAAG
CG
```

Microhomology:

```
3B-3C_reverse      -gactacaggcgcccgccaccacggctggctaatttttttgtagttttagtagagacagg
LRS                GGACTACAGGCGCCCGCCACCACGGCTGGCTAAT-TTTTTGTAGTTTTAGTAGAGACAGG
3E-3F_reverse      --------------------TGTAGAATTTTAATTCTTCCAAACATTTAATAGGA--AAG
                                           *     ****  **    *  **** ***    * *

3B-3C_reverse      gtttcaccgtgctagccaggatggtctcgatctcctgaccttgtgatccgcctgcctcag
LRS                GTTTCACCGTGTTAGCCTGGCCTTTGTTTGTCTGAAAGTCAAAGAACCTTCTACAAGTTC
3E-3F_reverse      GA---------AACAGATTCATTGTCTGTCTGAGGATGAGTTAAGTTCTTCTACAAGTTC
                   *                       * *   *                *  *

3B-3C_reverse      aat--cctaaagtgctgggattacaggcgtgagcc-------------------------
LRS                AACTGATAAGTCAGATGGGATTACGGGGAAGCG---------------------------
3E-3F_reverse      AACTGATAAGTCAGATGGGGATTACGGGGAAGGGTAAGTTCTTGTTTTTCTGTCTCCCTT
                   **      *    * ****  *   **     *

3B-3C_reverse      ---
LRS                ---
3E-3F_reverse      TAT
```

Blasting of the black insertion sequence traced it back to chr3:49.989.173-49.989.199 with a 100% identity match.

# 3D-3B

```
<- ->
GATAAGTCAGATGGGGATTACGGGGAAGGGTAAGTTCTTGTTTTTCTGTC
TCCCTTTATAAACTATTTCTGTTTTA|agtggctgatgcctgtagtttcag
ctactcaggaggctgagataggaggatcacttgagcccaggaattcgagt
ga
```

LRS:

```
TAAGTCAGATGGGGATTACTACGGGGAAGGGTAAGTTCTTGTTTTTCTGT
CTCCCTTTATAAACTATTTCTGTTTT|AGTAGCTGATGCCTGTAGTTTCAG
CTACTCAGGAGGCTGAGATAGGAGGATCACTTGAGCTCAGGAATTCGAGT
```

GA

Microhomology:

```
3C-3D_reverse        -CCCTTTATAAACTATTTCTGTTTTAATGAATATTAAATGTGCATTTTCAG
LRS                  TCCCTTTATAAACTATTTCTGTTTTAGTAGCTGATGCCTGTAGTTTCAGC-
3A-3B                AAAATGCATCAAGAGGgctgggcacagtggctgatgcctgtagtttcagc-
                         *  ** **        *    * *   *  *   ***   **
```

### Individual S5

#### Reference sequences

2A-2B: chr2:160624413-160624414

```
>hg38_dna range=chr2:160624339-160624490 5'pad=75 3'pad=75 strand=+
repeatMasking=lower
ttctcttagtagcaattgtaaatgggagttcactcatgatttggctctct
gtttgtctgttactggtatataggaa|tgcttgtgatttttgcacattgat
tttgtatcctgagactttgctgaagttgcttatcagcttaagattttggg
ct
```

```
>hg38_dna range=chr2:160624339-160624490 5'pad=75 3'pad=75 strand=-
repeatMasking=lower
agcccaaaatcttaagctgataagcaacttcagcaaagtctcaggataca
aaatcaatgtgcaaaaatcacaagca|ttcctatataccagtaacagacaa
acagagagccaaatcatgagtgaactcccatttacaattgctactaagag
aa
```

2B-2C: chr2:160814579-160814580

```
>hg38_dna range=chr2:160814504-160814655 5'pad=75 3'pad=75 strand=+
repeatMasking=lower
CAAAACATAATGTTGTTtatctatctagttatctatctatctatctatcc
atctatctatctatctatctatctTT|GATTCTGTTCATAATTCACCTTGT
CAAGTGCCAAGGACTCTCTTGCCTTCAAATTTAGACAAGAGCAAGTTTTC
AT
```

```
>hg38_dna range=chr2:160814504-160814655 5'pad=75 3'pad=75 strand=-
repeatMasking=lower
ATGAAAACTTGCTCTTGTCTAAATTTGAAGGCAAGAGAGTCCTTGGCACT
TGACAAGGTGAATTATGAACAGAATC|AAagatagatagatagatagatag
atggatagatagatagatagataactagatagataAACAACATTATGTTT
TG
```

2C-2D: chr2:160890958-160890959

```
>hg38_dna range=chr2:160890883-160891034 5'pad=75 3'pad=75 strand=+
repeatMasking=lower
cccatgggggagacagatatggCACAAAAGCGCACTCTGAGgaagatatt
agtacagaactcgcaggcctactctg|aggcctgaaagaaaaaatgcagga
aaaagcatttgtttctaggctgcagaaagtactcagcaaatgctagcCAT
TA
```

```
>hg38_dna range=chr2:160890883-160891034 5'pad=75 3'pad=75 strand=-
repeatMasking=lower
TAATGgctagcatttgctgagtactttctgcagcctagaaacaaatgctt
tttcctgcattttttctttcaggcct|cagagtaggcctgcgagttctgta
ctaatatcttcCTCAGAGTGCGCTTTTGTGccatatctgtctcccccatg
gg
```

2D-2E: chr2:161238636-161238637

```
>hg38_dna range=chr2:161238561-161238712 5'pad=75 3'pad=75 strand=+
repeatMasking=lower
AGAGAAAAGCAGCAGGCTGATTACGAGGTGTCAAAACTGCCAGGAGCAAG
AAGGTGATAGCAATCAGGGGTGAGAA|GAGTGCGGCATTCGTGCGGGGCAA
CTAATTATCCGTCTCATTTGAGAAGAGCAGCATTTGAGGCAGCAGCGTTC
```

```
GC
```

```
>hg38_dna range=chr2:161238561-161238712 5'pad=75 3'pad=75 strand=-
repeatMasking=lower
GCGAACGCTGCTGCCTCAAATGCTGCTCTTCTCAAATGAGACGGATAATT
AGTTGCCCCGCACGAATGCCGCACTC|TTCTCACCCCTGATTGCTATCACC
TTCTTGCTCCTGGCAGTTTTGACACCTCGTAATCAGCCTGCTGCTTTTCT
CT
```

2E-2F: chr2:161730485-161730486

```
>hg38_dna range=chr2:161730410-161730561 5'pad=75 3'pad=75 strand=+
repeatMasking=lower
attatctgtctatttatctattatctctcactcataaaacaacctattat
c
tatctatttatctattatctatcaa|ccatcatcattatcttctTTGTGA
TAAATCCTGAGAATGTAAGGTGGTCTCACATTCTAACAATAtttctgaat
at
```

```
>hg38_dna range=chr2:161730410-161730561 5'pad=75 3'pad=75 strand=-
repeatMasking=lower
atattcagaaaTATTGTTAGAATGTGAGACCACCTTACATTCTCAGGATT
TATCACAAagaagataatgatgatgg|ttgatagataatagataaatagat
agataataggttgttttatgagtgagagataatagataaatagacagata
at
```

2F-2G: chr2:227301893-227301894

```
>hg38_dna range=chr2:227301818-227301969 5'pad=75 3'pad=75 strand=+
repeatMasking=lower
ATCAGAAGCCAGCAAGTCCTTGACCAGAtggttctgttttgtctctctcc
tttttaacaacactttatccaagca|atcagatggttccttctttggtct
tcttgctccaaagatagctggttcaaggccagttttatccttggcacttt
tC
```

```
>hg38_dna range=chr2:227301818-227301969 5'pad=75 3'pad=75 strand=-
repeatMasking=lower
Gaaaagtgccaaggataaaactggccttgaaccagctatctttggagcaa
gaagaccaaagaaggaaccatctgat|tgcttggataaagtgttgttaaaa
aaggagagagacaaaacagaaccaTCTGGTCAAGGACTTGCTGGCTTCTG
AT
```

2G-2H: chr2:229052601-229052602

```
>hg38_dna range=chr2:229052526-229052677 5'pad=75 3'pad=75 strand=+
repeatMasking=lower
TGATGGGACTTGGCTTAGAGAGCTTTTTTTTTTCAACAAATGGGGATCAA
TAAAATATTTGCTGCTTTTAAGACA|GATTTATGTGTTGGTTTACTTTCTC
CTCCAGAAGCCCTCATACATGAAATAAATTTACTTGGTAATCTGTTGTCT
AA
```

```
>hg38_dna range=chr2:229052526-229052677 5'pad=75 3'pad=75 strand=-
repeatMasking=lower
TTAGACAACAGATTACCAAGTAAATTTATTTCATGTATGAGGGCTTCTGG
AGGAGAAAGTAAACCAACACATAAAT|CTGTCTTAAAAGCAGCAAATATTT
TATTGATCCCCATTTGTTGAAAAAAAAAAGCTCTCTAAGCCAAGTCCCAT
CA
```

2H-2I: chr2:229275096-229275097

```
>hg38_dna range=chr2:229275021-229275172 5'pad=75 3'pad=75 strand=+
repeatMasking=lower
```

aacctcccagactcaaagcaatccttccacctcagcctcctgaatagctg
ggactacaggtttgtgccaccacacc|cagctaattaaaaaaaaaatttat
ggagatagagtattgctgtgtttcccaggctccctgggctaaagtaattc
ac

>hg38_dna range=chr2:229275021-229275172 5'pad=75 3'pad=75 strand=-
repeatMasking=lower
gtgaattactttagcccagggagcctgggaaacacagcaatactctatct
ccataaattttttttttaattagctg|ggtgtggtggcacaaacctgtagt
cccagctattcaggaggctgaggtggaaggattgctttgagtctgggagg
tt

7A-7B: chr7:13292251-13292252
>hg38_dna range=chr7:13292176-13292327 5'pad=75 3'pad=75 strand=+
repeatMasking=lower
attattaattctcacaatatcttatgaaatggatactgtttttatccctg
tattttatggaaggaaactgaggttc|agaggatataagtaatttgcccaa
gttgtaaagccatctgagtaaagatggattttggaccgcggtcatgtaag
tt

>hg38_dna range=chr7:13292176-13292327 5'pad=75 3'pad=75 strand=-
repeatMasking=lower
aacttacatgaccgcggtccaaaatccatctttactcagatggctttaca
acttgggcaaattacttatatcctct|gaacctcagtttccttccataaaa
tacagggataaaaacagtatccatttcataagatattgtgagaattaata
at

7B-7C: chr7:130283621-130283622
>hg38_dna range=chr7:130283546-130283697 5'pad=75 3'pad=75 strand=+
repeatMasking=lower
tctcctaaagtgccgggattacagacgtgagccaccatgcctggccACCc
attctaacagacttttaaattaagta|tctattatatgttaagcattatgc
caagcactggagacataatgtgcccacacagaactctttaatagaagaga
tg

>hg38_dna range=chr7:130283546-130283697 5'pad=75 3'pad=75 strand=-
repeatMasking=lower
catctcttctattaaagagttctgtgtgggcacattatgtctccagtgct
tggcataatgcttaacatataataga|tacttaatttaaaagtctgttaga
atgGGTggccaggcatggtggctcacgtctgtaatcccggcactttagga
ga

### Variants

## 2A-2B

-> <-

Expected:
ttctcttagtagcaattgtaaatgggagttcactcatgatttggctctct
gtttgtctgttactggtatataggaa|AAagatagatagatagatagatag
atggatagatagatagatagataactagatagataAACAACATTATGTTT
TG

LRS:
TTCTCTTAGTAGCAATTGTAAATGGGAGTTCACTCATGATTTGGCTCTCT

GTTTGTCTGTTACTGGTATATAGGAA|AAGATAGATAGATAGATAGATAGA
TGGATACATAGATAGATAGATAACTAAATAGATAAACAACATTATGTTTT
GG

Microhomology:

```
blauw_onder              -tttgtctgttactggtatataggaatgcttgtg-atttttgcacattgatt
LRS                      -TTTGTCTGTTACTGGTATATAGGAAAAGATAGATAGATAGATAGATAGAT-
blauw_boven_reverse      GACAAGGTGAATTATGAACAGAATCAAagatagatagatagatagataga--
                                **      * * * *   *    *    *  *    * ** **
```

## 2B-2C

<-  ->

Expected:

ccaaaatcttaagctgataagcaacttcagcaaagtctcaggatacaaaa
tcaatgtgcaaaaatcacaagcattc|tctTTGATTCTGTTCATAATTCAC
CTTGTCAAGTGCCAAGGACTCTCTTGCCTTCAAATTTAGACAAGAGCAAG
TT

LRS:

AGCCCAAAATCTAAGCTGATAAGCAACTTCAGCAAAGTCTCAGGATACAA
AATCAATGTGCAAAAATCACAAGCAT|TCTTTGATTATCTTCCATAATTCA
CCTTGTCAAGTGCCAAGGACTCTCTTGCCTTCAAATTTAGACAAGAGCAA
GT

Microhomology:

```
oranje_boven_reverse      --caatgtgcaaaaatcacaagcattcctatataccagtaacagacaaacag
LRS                       ATCAATGTGCAAAAATCACAAGCATTCTTTGATTATCTTCCATAATTCAC--
oranje_onder              atccatctatctatctatctatctatctTTGATTCTGTTCATAATTCACC--
                            * ** *    *  *  * * *  ** *  **     *          *
```

## 2C-2F

-> ->

Expected:

cccatggggagacagatatggCACAAAAGCGCACTCTGAGgaagatatt
agtacagaactcgcaggcctactctg|ccatcatcattatcttctTTGTGA
TAAATCCTGAGAATGTAAGGTGGTCTCACATTCTAACAATAtttctgaat
at

LRS:

CCATGGGGGAGACAGATATGGCACAAAAGCGCACTCTGAGGAAGATATTA
GTACAGAACTCGCAGGCCTACTCTG|TCCATCATCATTATCTTCTTTGTGA
TAAATCCTGAGAATGTAAGGTGGTCTCACATTCTAACAATATTTCTGAAT
AT

Microhomology:

```
geel_links       gtacagaactcgcaggcctactctgaggcctgaaagaaaaaatgcaggaa-
LRS              -TACAGAACTCGCAGGCCTACTCTGTCCATCATCATTATCTTCTTTGTGAT
geel_rechts      -tatctatttatctattatctatcaaccatcatcattatcttctTTGTGAT
                  **   *  *  *     *               *  *      *  *
```

## 7A-2D

-> <-

Expected:

attattaattctcacaatatcttatgaaatggatactgtttttatccctg
tattttatggaaggaaactgaggttc|TTCTCACCCCTGATTGCTATCACC
TTCTTGCTCCTGGCAGTTTTGACACCTCGTAATCAGCCTGCTGCTTTTCT

CT

LRS:
ATTATTAATTCTCACAATATCTTATGAAATGGATACTGTTTTTATCCCTG
TATTTTATGGAAGGAAACTGAGGTTC|TCACCCCTGATTGCTATCACCTTC
TTGCTCCTGGCAGTTTTGACACCTCGTAATCAGCCTGCTGCTTTCTCTTC
TT

Microhomology:
```
lichtgroen_chr7            atttatggaaggaa---actgaggttcagaggatataagtaatttgcccaag
LRS                        ATTTTATGGAAGGAA---ACTGAGGTTCTCACCCCTGATTGCTATCACCTTCT
lichtgroen_chr2_reverse    GTTGCCCCGCACGAATGCCGCACTCTTCTCACCCCTGATTGCTATCACCT---

                            **     * * ***         ***  *      *      *  **
```

## 2D-7B

<-  ->

Expected:
GTAATGgctagcatttgctgagtactttctgcagcctagaaacaaatgct
ttttcctgcatttttttctttcaggcc|aggttcagaggatataagtaattt
gcccaagttgtaaagccatctgagtaaagatggattttggaccgcggtca
tg

LRS:
TAATGGCTAGCATTTGCTGAGTACTTTTCTGCAGCCTAGAAACAAATGCT
TTTTCCTGCATTTTTTCTTTCAGGCC|AGGTTCAGAGGATATAAGTAATTT
GCCCAAGTTGTAAAGCCATCTGAGTAAAGATGGATTTTTCTGGACCGCGG
TC

Microhomology:
```
rood_chr2_reverse     tttcctgcatttttttctttcaggcctcagagtaggcctgcgagttctgta
LRS                   TTTCCTGCATTTTTTCTTTCAGGCCAGGTTCAGAGGATATAAGTAATTTG
rood_chr7             ccctgtattttatggaaggaaactgaggttcagaggatataagtaatttg
                           *   ** *       *             *  *   ***  * *
```

## 7B-2G

->  ->

Expected:
tctcctaaagtgccgggattacagacgtgagccaccatgcctggccACCc
attctaacagacttttaaattaagta|acactttatccaagcaatcagatg
gttccttctttggtcttcttgctccaaagatagctggttcaaggccagtt
tt

LRS:
GTCTCCTAAAGTGCCGGGATTACAGACGTGAGCCACCATGCCTGGCCACC
CATTCTAACAGACTTTTAAATTAAGT|ACACTTTATCCAAGCAATCAGATG
GTTCCTTCTTTGGTCTTCTTGCTCCAAAGATAGCTGGTTCAAGGCCAGTT
TT

Microhomology:
```
roze_chr7     -ttctaacagacttttaaattaagtatctattatatgttaagcattatgcc
LRS           ATTCTAACAGACTTTTAAATTAAGTACACTTTATCCAAGCAATCAGATGG-
roze_chr2     gttttgtctctctccttttttaacaacactttatccaagcaatcagatgg-
               ** *  *   **  *   ****  *    ****      *     ***
```

## 2F-7C

-> ->

Expected:

ATCAGAAGCCAGCAAGTCCTTGACCAGAtggttctgttttgtctctctcc
tttttaacaacactttatccaagca|aagcattatgccaagcactggaga
cataatgtgcccacacagaactctttaatagaagagatgaacaggaaggc
aa

LRS:

ATCAGAAGCCAGCAAGTCCTTGACCAGATGGTTCTGTTTTGTCTCTCTCC
TTTTTAACAAGCATTATGCCAAGCA|CTGGAGACATAATGTGCCCACACA
GAACTCTTTAATAGAAGAGATGAACAGGGAAGGCAAATGAACTTTTGTAT
GA

Microhomology:

```
paars_chr2      -----------------tttttaacaacactttatccaagcaatcagatggttccttctt
LRS             -----------------TTTTTAACAAGCATTATGCCAAGCACTGGAGAC----ATAATG
paars_chr7      ttaaattaagtatctattatatgttaagcattatgccaagcactggagac----------
                                 * * *   **   **   ******* *

paars_chr2      tggtctt----
LRS             TGCCCACACAG
paars_chr7      -----------
```

# 2G-2I

-> ->

Expected:

TGATGGGACTTGGCTTAGAGAGCTTTTTTTTTTCAACAAATGGGGATCAA
TAAAATATTTGCTGCTTTTAAGACA|cagctaattaaaaaaaaaatttat
ggagatagagtattgctgtgtttcccaggctccctgggctaaagtaattc
ac

LRS:

TGATGGGACTTGGCTTAGAGAGCTTTTTTTTTTCAACAAATGGGGATCAA
TAAAATATTTGCTGCTTTTAAGACAG|CTAATTAAAAAAAAAATTTATGGA
GATAGAGTATTGCTGTGTTTCCCAGGCTCCCTGGGCTAAAGTAATTCACT
TG

Microhomology:

```
donkergroen_links       ---AAAATATTTGCTGCTTTTAAGACAGATTTATGTGTTGGTTTACTTTCTCC
LRS                     ---AAAATATTTGCTGCTTTTAAGACAGCTAATTAAAAAAAAAATTTATGGAG
donkergroen_rechts      gactacaggtttgtgccaccacacccagctaattaaaaaaaaaatttatg---
                            * *  ****   *     *  *** *   *            * *
```

### Individual S6

#### Reference sequences

9A-9B – chr9:12273020-12273021

```
>hg38_dna range=chr9:12272945-12273096 5'pad=75 3'pad=75 strand=+
repeatMasking=lower
TTTCTAACATGCCAGAGGCTCTTCTCATTGCCAAAATTTCTCCTTTGTCA
GGCTCCATAGGAAACTGATGTAGTCC|ATGCTCAAAAGACATCCAATAAAA
AGCTATGAAGCTGCCACTGATTCACATCTCCACTGGAGGGGGAAAAAAAA
AA
```

```
>hg38_dna range=chr9:12272945-12273096 5'pad=75 3'pad=75 strand=-
repeatMasking=lower
TTTTTTTTTTCCCCCTCCAGTGGAGATGTGAATCAGTGGCAGCTTCATAG
CTTTTTATTGGATGTCTTTTGAGCAT|GGACTACATCAGTTTCCTATGGAG
CCTGACAAAGGAGAAATTTTGGCAATGAGAAGAGCCTCTGGCATGTTAGA
AA
```

9B-9C – chr9: 12280621-12280622

```
>hg38_dna range=chr9:12280546-12280697 5'pad=75 3'pad=75 strand=+
repeatMasking=lower
ATTATTTCCAATTGGAATATCAGTATTCTTTGTTTTCTTCAACTTCCCTG
ACTCTCTACCTATGCTCCTTTGCACT|TGAATGATTCTTATTAGCCATGAG
GCAATGCTTGCTCTTGTCAACACATTCTATCCATCCCTTTAGTGAagcag
tc
```

```
>hg38_dna range=chr9:12280546-12280697 5'pad=75 3'pad=75 strand=-
repeatMasking=lower
gactgctTCACTAAAGGGATGGATAGAATGTGTTGACAAGAGCAAGCATT
GCCTCATGGCTAATAAGAATCATTCA|AGTGCAAAGGAGCATAGGTAGAGA
GTCAGGGAAGTTGAAGAAAACAAAGAATACTGATATTCCAATTGGAAATA
AT
```

10A-10B – chr10: 47661049-47661050

```
>hg38_dna range=chr10:47660974-47661125 5'pad=75 3'pad=75 strand=+
repeatMasking=lower
TTAACACAATATTGGCAAGCTAAATTTAATCAAATATTCTGCCCATTTTA
AAATTCACTTCCTagcctgtccatcc|ttccttttattttgttgtttgatt
ttgttttattgtatgcttttagcagcctgaagccatggtttttagtttct
gt
```

```
>hg38_dna range=chr10:47660974-47661125 5'pad=75 3'pad=75 strand=-
repeatMasking=lower
acagaaactaaaaaccatggcttcaggctgctaaaagcatacaataaaac
aaaatcaaacaacaaaataaaaggaa|ggatggacaggctAGGAAGTGAAT
TTTAAAATGGGCAGAATATTTGATTAAATTTAGCTTGCCAATATTGTGTT
AA
```

10B-10C – chr10: 47665532-47665533

```
>hg38_dna range=chr10:47665457-47665608 5'pad=75 3'pad=75 strand=+
repeatMasking=lower
CATGATAATGTATTTCTCCATTGAAACAGTATGGTTTGAACAGTGCTATA
TATTCATACTCATTATTATTCCTTAA|ATGAATATTTTAAGCTCCTGATAG
AAGGCAGGTACTGTGTTATATGCTGTTTGTGTTACCAGAATACAAAGATG
```

AA

>hg38_dna range=chr10:47665457-47665608 5'pad=75 3'pad=75 strand=-
repeatMasking=lower
TTCATCTTTGTATTCTGGTAACACAAACAGCATATAACACAGTACCTGCC
TTCTATCAGGAGCTTAAAATATTCAT|TTAAGGAATAATAATGAGTATGAA
TATATAGCACTGTTCAAACCATACTGTTTCAATGGAGAAATACATTATCA
TG

10C-10D – chr10: 48452031-48452032
>hg38_dna range=chr10:48451956-48452107 5'pad=75 3'pad=75 strand=+
repeatMasking=lower
TCCGCCACACACAATTGCACACACAATGCACCCTCCGCAGCCTCCCCAAA
TTCCCCCATACACAACTGCACACATA|GTGCACCCTGCTCACCCCCCAGAA
TCCCACATACATATAATCTCGCACACACACAATCCCCCGTGTCTAGGCCA
AG

>hg38_dna range=chr10:48451956-48452107 5'pad=75 3'pad=75 strand=-
repeatMasking=lower
CTTGGCCTAGACACGGGGGATTGTGTGTGTGCGAGATTATATGTATGTGG
GATTCTGGGGGGTGAGCAGGGTGCAC|TATGTGTGCAGTTGTGTATGGGGG
AATTTGGGGAGGCTGCGGAGGGTGCATTGTGTGTGCAATTGTGTGTGGCG
GA

10D-10E – chr10: 48482257-48482258
>hg38_dna range=chr10:48482182-48482333 5'pad=75 3'pad=75 strand=+
repeatMasking=lower
ttcatctaaggccataaaagttttcttctaggttttctttcaaaagtttc
acagttttaggatttacatgtaggtc|tatgagttacttttttgtatatggg
agaaggtgtaattgaagttcattcttttgcatatgcacattccagcccca
ct

>hg38_dna range=chr10:48482182-48482333 5'pad=75 3'pad=75 strand=-
repeatMasking=lower
agtggggctggaatgtgcatatgcaaaagaatgaacttcaattacacctt
ctcccatatacaaaaagtaactcata|gacctacatgtaaatcctaaaact
gtgaaacttttgaaagaaaacctagaagaaaacttttatggccttagatg
aa

10E-10F – chr10: 49302110-49302111
>hg38_dna range=chr10:49302035-49302186 5'pad=75 3'pad=75 strand=+
repeatMasking=lower
ATGGCTGGAGGGAAGGATTTCATCTGGGATATCTCCCACCTCAAAGCCTT
CACCTGAAGCTTGAGGAGGGCAGGGG|GAAGGGGAGGCTCTCCAATTTGCT
TGGGAAGCCTCTGAAGGAGTTTCTGAGGCTCTCTCCCTACTCCCCACTGT
GT

>hg38_dna range=chr10:49302035-49302186 5'pad=75 3'pad=75 strand=-
repeatMasking=lower
ACACAGTGGGGAGTAGGGAGAGAGCCTCAGAAACTCCTTCAGAGGCTTCC
CAAGCAAATTGGAGAGCCTCCCCTTC|CCCCTGCCCTCCTCAAGCTTCAGG
TGAAGGCTTTGAGGTGGGAGATATCCCAGATGAAATCCTTCCCTCCAGCC
AT

10F-10G – chr10: 49311963-49311964
>hg38_dna range=chr10:49311888-49312039 5'pad=75 3'pad=75 strand=+
repeatMasking=lower

```
AGGATTAGGAGGGGGAGAGAATTGTATCCGCAAAAAGCAGCACATCCCAG
TCAATAATAACAGCATCATGAGGCAT|GGCATGGAAAAAAAGCATCATGTG
AGAAAACATTTTGGAAGAGTTAGAATTTAGCAATGACTGTGCAAAGGGTA
TT

>hg38_dna range=chr10:49311888-49312039 5'pad=75 3'pad=75 strand=-
repeatMasking=lower
AATACCCTTTGCACAGTCATTGCTAAATTCTAACTCTTCCAAAATGTTTT
CTCACATGATGCTTTTTTTCCATGCC|ATGCCTCATGATGCTGTTATTATT
GACTGGGATGTGCTGCTTTTTGCGGATACAATTCTCTCCCCCTCCTAATC
CT
```

10G-10H – chr10: 49564013-49564014

```
>hg38_dna range=chr10:49563938-49564089 5'pad=75 3'pad=75 strand=+
repeatMasking=lower
ttagaatcaccttcagctgcatgagcaaacacactaaatccagaatgtcc
aataagagcacccatattcacaatca|gggctgatctagtgttagattcca
ccctccttggtgtaacgcccttagaatgcacttccacacatgcatcctag
gc

>hg38_dna range=chr10:49563938-49564089 5'pad=75 3'pad=75 strand=-
repeatMasking=lower
gcctaggatgcatgtgtggaagtgcattctaagggcgttacaccaaggag
ggtggaatctaacactagatcagccc|tgattgtgaatatgggtgctctta
ttggacattctggatttagtgtgtttgctcatgcagctgaaggtgattct
aa
```

10H-10I – chr10: 49589506-49589507

```
>hg38_dna range=chr10:49589431-49589582 5'pad=75 3'pad=75 strand=+
repeatMasking=lower
gatagtgaaaatagGCTGTTGTTATGAGGtcatcttgagtcacttttggt
aatttgtatgcatattagtccattct|cacactgctataaagacatacctt
agactgggtaatttatataaagaaaaagatgtttaattggctcactgttc
tg

>hg38_dna range=chr10:49589431-49589582 5'pad=75 3'pad=75 strand=-
repeatMasking=lower
cagaacagtgagccaattaaacatctttttctttatataaattacccagt
ctaaggtatgtctttatagcagtgtg|agaatggactaatatgcatacaaa
ttaccaaaagtgactcaagatgaCCTCATAACAACAGCctattttcacta
tc
```

10I-10J – chr10: 49651674-49651675

```
>hg38_dna range=chr10:49651599-49651750 5'pad=75 3'pad=75 strand=+
repeatMasking=lower
AACACAGCCAGGCCCACCTCCCCACCCACTCTCCAGAGCAGGGCCGACAC
CCCACTCAGCTGAGCTCACCTTGGGT|CATAGTAGTCCTGGCCTGGTTTGT
CTTCTGACACTTTCATAGGTCAAGGCATGTCTCTGACACCTCAGTGCACT
CC

>hg38_dna range=chr10:49651599-49651750 5'pad=75 3'pad=75 strand=-
repeatMasking=lower
GGAGTGCACTGAGGTGTCAGAGACATGCCTTGACCTATGAAAGTGTCAGA
AGACAAACCAGGCCAGGACTACTATG|ACCCAAGGTGAGCTCAGCTGAGTG
GGGTGTCGGCCCTGCTCTGGAGAGTGGGTGGGGAGGTGGGCCTGGCTGTG
TT
```

10J-10K – chr10: 49682049-49682050

>hg38_dna range=chr10:49681974-49682125 5'pad=75 3'pad=75 strand=+
repeatMasking=lower
CTAAGAAGACAGCCTAAATAAGTGACTTTTCTCTTCCTCCCCCTCTGATG
AGCCACACCTACCCACCCTCACCTCA|TAATCTACCTTTAGTTTATGTTCA
CTGTCTCTAAAAATAACGTGggtaaaacatatatttaaaatgttgccatt
tt

>hg38_dna range=chr10:49681974-49682125 5'pad=75 3'pad=75 strand=-
repeatMasking=lower
aaaatggcaacattttaaatatatgttttaccCACGTTATTTTTAGAGAC
AGTGAACATAAACTAAAGGTAGATTA|TGAGGTGAGGGTGGGTAGGTGTGG
CTCATCAGAGGGGGAGGAAGAGAAAAGTCACTTATTTAGGCTGTCTTCTT
AG

10K-10L – chr10: 54276117-54276118
>hg38_dna range=chr10:54276042-54276193 5'pad=75 3'pad=75 strand=+
repeatMasking=lower
ataattaccaaaaatgtaagtcatcatctttatggtatttaaataaaaaa
aagacttctttaaaaagacacaaaga|aagcaaaacccataaagagaaaaa
ctgatactcttgaagatgttcaagtttaaaactcatgcaaaatagcacca
aa

>hg38_dna range=chr10:54276042-54276193 5'pad=75 3'pad=75 strand=-
repeatMasking=lower
tttggtgctattttgcatgagttttaaacttgaacatcttcaagagtatc
agtttttctctttatgggttttgctt|tctttgtgtctttttaaagaagtc
ttttttttatttaaataccataaagatgatgacttacatttttggtaatt
at

10L-10M – chr10: 54315782-54315783
>hg38_dna range=chr10:54315707-54315858 5'pad=75 3'pad=75 strand=+
repeatMasking=lower
atggtataaggaggggggggtccagcttcaatcttcatatggctagccag
ttctcccagcaccatttactgaatag|gaagtctttttgccactacttttg
tcagctttgtcgaagggcagatggcagtaggtgtgcagccttatAAAAAT
TA

>hg38_dna range=chr10:54315707-54315858 5'pad=75 3'pad=75 strand=-
repeatMasking=lower
TAATTTTTataaggctgcacacctactgccatctgcccttcgacaaagct
gacaaaagtagtggcaaaaagacttc|ctattcagtaaatggtgctgggag
aactggctagccatatgaagattgaagctggaccccccccctccttatacc
at

10M-10N – chr10: 54340702-54340703
>hg38_dna range=chr10:54340627-54340778 5'pad=75 3'pad=75 strand=+
repeatMasking=lower
ttagagcaggaatgaaagcaagcaaaatactcttggaagacggctaagtg
ggtgacttgaaagatccaagtgcctt|gttcatcccttcacttggggtttt
tctacattggcatggtttgggggtttgccattctcctcccttgatttttc
tt

>hg38_dna range=chr10:54340627-54340778 5'pad=75 3'pad=75 strand=-
repeatMasking=lower
aagaaaaatcaagggaggagaatggcaaacccccaaaccatgccaatgta
gaaaacccccaagtgaagggatgaac|aaggcacttggatctttcaagtca
cccacttagccgtcttccaagagtattttgcttgctttcattcctgctct
aa

10N-10O – chr10: 54378120-54378121

```
>hg38_dna range=chr10:54378045-54378196 5'pad=75 3'pad=75 strand=+
repeatMasking=lower
ctgcagattagtgggactacaggcacgtgttaccatgcctgcctagtttt
tcaaatttttgtcaagacagagtct|tactctatttgccaggctggactc
aaactcctgggctcaagcagttctccccactgggcctcccaaagtgctgg
ga

>hg38_dna range=chr10:54378045-54378196 5'pad=75 3'pad=75 strand=-
repeatMasking=lower
tcccagcactttgggaggcccagtggggagaactgcttgagcccaggagt
ttgagtccagcctggcaaatagagta|agactctgtcttgacaaaaaattt
gaaaaactaggcaggcatggtaacacgtgcctgtagtcccactaatctgc
ag
```

10O-10P – chr10: 55429308-55429309

```
>hg38_dna range=chr10:55429233-55429384 5'pad=75 3'pad=75 strand=+
repeatMasking=lower
gaaattttaagcacttcttataagatggatttgctggttattaactattt
catatttataggtctaaaacgtcatt|gttccactttgtttttgaaagata
tttttgctaatgataggacactagtaatacaggtttaggctttacctccc
tt

>hg38_dna range=chr10:55429233-55429384 5'pad=75 3'pad=75 strand=-
repeatMasking=lower
aagggaggtaaagcctaaacctgtattactagtgtcctatcattagcaaa
aatatctttcaaaaacaaagtggaac|aatgacgttttagacctataaata
tgaaatagttaataaccagcaaatccatcttataagaagtgcttaaaatt
tc
```

10P-10Q – chr10: 55444307-55444308

```
>hg38_dna range=chr10:55444232-55444383 5'pad=75 3'pad=75 strand=+
repeatMasking=lower
atgtaacaaacctgcacattctgcacatgtaccccagaaagtaaagtgta
attaaaaaaaaaaaGAAAACAAGAG|TGCTAGTTTTATTTTTTTATTTTA
AAGTTTCTATTTTTATAAGCAGACACACAAATACACATGTACATCTTTAT
GT

>hg38_dna range=chr10:55444232-55444383 5'pad=75 3'pad=75 strand=-
repeatMasking=lower
ACATAAAGATGTACATGTGTATTTGTGTGTCTGCTTATAAAAATAGAAAC
TTTAAAATAAAAAAATAAAACTAGCA|CTCTTGTTTTCtttttttttttta
attacactttactttctggggtacatgtgcagaatgtgcaggtttgttac
at
```

10Q-10R – chr10: 55481914-55481915

```
>hg38_dna range=chr10:55481839-55481990 5'pad=75 3'pad=75 strand=+
repeatMasking=lower
tctatcaggtgcatttgatccagcactgagttcaggtcctgaatatcttt
gttaattttctgtctcagtgatctaa|tactgtcagtggggtgttaaattt
ccccactataattgtgtggtagtctatgtatctttgaaggtctttaagaa
ct

>hg38_dna range=chr10:55481839-55481990 5'pad=75 3'pad=75 strand=-
repeatMasking=lower
agttcttaaagaccttcaaagatacatagactaccacacaattatagtgg
ggaaatttaacaccccactgacagta|ttagatcactgagacagaaaatta
```

acaaagatattcaggacctgaactcagtgctggatcaaatgcacctgata
ga

10R-10S – chr10: 55508209-55508210

>hg38_dna range=chr10:55508134-55508285 5'pad=75 3'pad=75 strand=+
repeatMasking=lower
AATTAAAGTGGGATACATTTTACATGTTTTGGCTCTACCATAAATTTTAG
CATGACTTAAGAAAATAACAGAAATG|CATACATGCCTGCATTCTGTAAGG
ACTGTGCTATCCTTTGGTTCAATGAAATTAGCATGGGAAGAGAGCAGTTC
CA

>hg38_dna range=chr10:55508134-55508285 5'pad=75 3'pad=75 strand=-
repeatMasking=lower
TGGAACTGCTCTCTTCCCATGCTAATTTCATTGAACCAAAGGATAGCACA
GTCCTTACAGAATGCAGGCATGTATG|CATTTCTGTTATTTTCTTAAGTCA
TGCTAAAATTTATGGTAGAGCCAAAACATGTAAAATGTATCCCACTTTAA
TT

10S-10T – chr10: 55936316-55936317

>hg38_dna range=chr10:55936241-55936392 5'pad=75 3'pad=75 strand=+
repeatMasking=lower
TATTGGCCTTTATTTTAGTGGTTCCAAAGAAAACAGGGCAAAttcatata
tgaagtctaattaaataaaggcagat|ggctacatattttctcttcctggt
agggacagatcggtgtaagaaggcttaaagtcaacatgaaaggtttagAA
AA

>hg38_dna range=chr10:55936241-55936392 5'pad=75 3'pad=75 strand=-
repeatMasking=lower
TTTTctaaacctttcatgttgactttaagccttcttacaccgatctgtcc
ctaccaggaagagaaaatatgtagcc|atctgcctttatttaattagactt
catatatgaaTTTGCCCTGTTTTCTTTGGAACCACTAAAATAAAGGCCAA
TA

10T-10U – chr10: 55968259-55968260

>hg38_dna range=chr10:55968184-55968335 5'pad=75 3'pad=75 strand=+
repeatMasking=lower
ACTTCCTATTGGATGACTCCACTAAGGCAATCACCAGACTCCGGAGTCTG
GCACACAAGGCCTCCGCCACCTAAGC|ACTTGATTAAGATCTACTATTATT
ATTATAAGTGATATTacctgaccgttgtctcttctcatgccttgtcaatc
tc

>hg38_dna range=chr10:55968184-55968335 5'pad=75 3'pad=75 strand=-
repeatMasking=lower
gagattgacaaggcatgagaagagacaacggtcaggtAATATCACTTATA
ATAATAATAGTAGATCTTAATCAAGT|GCTTAGGTGGCGGAGGCCTTGTGT
GCCAGACTCCGGAGTCTGGTGATTGCCTTAGTGGAGTCATCCAATAGGAA
GT

10U-10V – chr10: 57854781-57854782

>hg38_dna range=chr10:57854706-57854857 5'pad=75 3'pad=75 strand=+
repeatMasking=lower
gttacacaggtgcccgtgggccatagtggtttgctgcacctattgacccg
tcctctaagttccctcctctcacact|ccacccctcaacaggccctggtgt
gtgttgttcccctctctgtgtccctgtgttctcaatgttcaactcccact
ta

>hg38_dna range=chr10:57854706-57854857 5'pad=75 3'pad=75 strand=-
repeatMasking=lower
taagtgggagttgaacattgagaacacagggacacagagaggggaacaac

a
cacaccagggcctgttgaggggtgg|agtgtgagaggagggaacttagag
gacgggtcaataggtgcagcaaaccactatggcccacgggcacctgtgta
ac

10V-10W – chr10: 57866072-57866073
>hg38_dna range=chr10:57865997-57866148 5'pad=75 3'pad=75 strand=+
repeatMasking=lower
gaaaaagtacatctcagaatatgccactgtgaaacagcacatttcatctg
gcaaactgtgggtatctcagcatggt|tggcacattgagcagtatcagaga
atgataagaaataaaactaaagaaagtaagcaggtgccagatGACTAGGG
CC

>hg38_dna range=chr10:57865997-57866148 5'pad=75 3'pad=75 strand=-
repeatMasking=lower
GGCCCTAGTCatctggcacctgcttactttctttagttttatttcttatc
attctctgatactgctcaatgtgcca|accatgctgagatacccacagttt
gccagatgaaatgtgctgtttcacagtggcatattctgagatgtactttt
tc

10W-10X – chr10: 58394858-58394859
>hg38_dna range=chr10:58394783-58394934 5'pad=75 3'pad=75 strand=+
repeatMasking=lower
TCTCCAGTCTGCCTTTATACATGTAGAATGGTAATGTAATTTCTAAAGTA
TATACAGAAGCATTCCAGAAATTAAT|TGCTATTTTAAATAATCATTTTAT
CTCAAAAAATAAGTAAATCATTTTAACAGTTATGCTTTTTCTCAGTTATA
TA

>hg38_dna range=chr10:58394783-58394934 5'pad=75 3'pad=75 strand=-
repeatMasking=lower
TATATAACTGAGAAAAAGCATAACTGTTAAAATGATTTACTTATTTTTTG
AGATAAAATGATTATTTAAAATAGCA|ATTAATTTCTGGAATGCTTCTGTA
TATACTTTAGAAATTACATTACCATTCTACATGTATAAAGGCAGACTGGA
GA

10X-10Y – chr10: 58465105-58465106
>hg38_dna range=chr10:58465030-58465181 5'pad=75 3'pad=75 strand=+
repeatMasking=lower
tggccaggggaagtattctggtttctcaagcaatgggtgggaccataaag
ctcctaagagtttatgtgtttgtctt|cagctaccagggcaggtagagaac
taccattaggtggcggtagggttaggcgggtctgttctcagactctcctt
gg

>hg38_dna range=chr10:58465030-58465181 5'pad=75 3'pad=75 strand=-
repeatMasking=lower
ccaaggagagtctgagaacagacccgcctaaccctaccgccacctaatgg
tagttctctacctgccctggtagctg|aagacaaacacataaactcttagg
agctttatggtcccacccattgcttgagaaaccagaatacttcccctggc
ca

14A-14B – chr14: 103527762-103527763
>hg38_dna range=chr14:103527687-103527838 5'pad=75 3'pad=75 strand=+
repeatMasking=lower
GGAGCTGTGGAGGGGGCTGTTTCAACGTTGAGAGCAGAAGGCAGGGAGGC
TGTAAGGCAGAAGTAGAGGCAGCTGG|TGGCCAGCGTGGTGGCCCCACAGG
GACAGGAGCCCACAGAGAGTGGGAGGCCATGGAAGGCCTCTCACCAGCTT
AC

>hg38_dna range=chr14:103527687-103527838 5'pad=75 3'pad=75 strand=-
repeatMasking=lower
GTAAGCTGGTGAGAGGCCTTCCATGGCCTCCCACTCTCTGTGGGCTCCTG
TCCCTGTGGGGCCACCACGCTGGCCA|CCAGCTGCCTCTACTTCTGCCTTA
CAGCCTCCCTGCCTTCTGCTCTCAACGTTGAAACAGCCCCCTCCACAGCT
CC

14B-14C – chr14: 103559220-103559221
>hg38_dna range=chr14:103559145-103559296 5'pad=75 3'pad=75 strand=+
repeatMasking=lower
AATGCAATCTCAAAGGTTTTTTGGCTATTAGTTTTCATAATTTTCTTATG
TTGCACACAAAAACAAGATTCCTCTC|TAAAACGTAGAGGATGGGGAAAAT
GCAGATGCTGTTTTTCCAACTAAAAATGTTTACAAAAGAACAGACTGTCT
GA

>hg38_dna range=chr14:103559145-103559296 5'pad=75 3'pad=75 strand=-
repeatMasking=lower
TCAGACAGTCTGTTCTTTTGTAAACATTTTTAGTTGGAAAAACAGCATCT
GCATTTTCCCCATCCTCTACGTTTTA|GAGAGGAATCTTGTTTTTGTGTGC
AACATAAGAAAATTATGAAAACTAATAGCCAAAAAACCTTTGAGATTGCA
TT

### Variants

## 9A-10K

-> <-

Expected:
TTTCTAACATGCCAGAGGCTCTTCTCATTGCCAAAATTTCTCCTTTGTCA
GGCTCCATAGGAAACTGATGTAGTCC|tctttgtgtctttttaaagaagtc
tttttttatttaaataccataaagatgatgacttacattttggtaatt
at

LRS:
TTTCTAACATGCCAGAGGCTCTTCTCATTGCCAAAATTTCTCCTTTGTCA
GGCTCCATAGGAAACTGATGTAGTCC|TCTTTGTGTCTTTTTAAAGAAGTC
TTTTTTTATTTAAATACCATAAAGATGATGACTTACATTTTTGTAATTAT
GT

Microhomology:
```
9A-9B                GCTCCATAGGAAACTGATGTAGTCCATGCTCAAAAGACATCCAATAAAAA
LRS                  GCTCCATAGGAAACTGATGTAGTCCTCTTTGTGTCTTTTTAAAGAAGTCT
10K-10L_reverse      gtttttctctttatgggttttgctttctttgtgtctttttaaagaagtct
                     * *         *  * * * *       *         *  *  *
```

## 10K-10G

<-    ->

Expected:
aaaatggcaacattttaaatatatgttttaccCACGTTATTTTTAGAGAC
AGTGAACATAAACTAAAGGTAGATTA|GGCATGGAAAAAAAGCATCATGTG
AGAAAACATTTTGGAAGAGTTAGAATTTAGCAATGACTGTGCAAAGGGTA
TT

LRS:
AAATGGCAACATTTTAAATATATGTTTTACCCACGTTATTTTTAGAGACA
GTGAACATAAACTAAAGGTAGATTA|GGCATGGAAAAAAGCATCATGTGAG
AAAACATTTTGGAAGAGTTAGAATTTAGCAATGACTGTGCAAAGGGTATT
TC

Microhomology:

```
10J-10K_reverse      -GTGAACATAAACTAAAGGTAGATTATGAGGTGAGGGTGGGTAGGTGTGGC-
LRS                  --TGAACATAAACTAAAGGTAGATTAGGCATGGAAAAAAGCATCATGTGAGA
10F-10G              CAATAATAACAGCATCATGAGGCATGGCATGGAAAAAAGCATCATGTGA--
                         ** *  * *   * *  *  *        *     *     ****
```

# 10G-10U

-> <-

Expected:
ttagaatcaccttcagctgcatgagcaaacacactaaatccagaatgtcc
aataagagcacccatattcacaatca|agtgtgagaggagggaacttagag
gacgggtcaataggtgcagcaaaccactatggcccacgggcacctgtgta
ac

LRS:
TTAGAATCACCTTCAGCTGCATGAGCAAACACACTAAATCCAGAATGTCC
AATAAGAGCACCCATATTCACAATCA|GAGTGTGAGAGGAGGAACTTAGAG
GACGGGTCAATAGGTGCAGCAAACCACTATGGCCCACGGGCACCTGTGTA
AC

Microhomology:

```
10G-10H              ataagagcacccatattcacaatcagggctgatctagtgttagattccac-
LRS                  ATAAGAGCACCCATATTCACAATCAGAGTGTGAGAGGAGGAACTTAGAGG-
10U-10V_reverse      -cacaccagggcctgttgaggggtggagtgtgagaggagggaacttagagg
                       *        * * ** *      * *        * *  *  *
```

# 10U-10O

<- ->

Expected:
gagattgacaaggcatgagaagagacaacggtcaggtAATATCACTTATA
ATAATAATAGTAGATCTTAATCAAGT|tactctatttgccaggctggactc
aaactcctgggctcaagcagttctccccactgggcctcccaaagtgctgg
ga

LRS:
GGAGATTGACAAGGCATGAGAAGAGACAACGGTCAGGTAATATCACTTAT
AATAATAATAGTAGATTCAATCAAGT|ACTCTATTTGCCAGGCTGGACTCA
AACTCCTGGGCTCAAGCAGTTCTCCCCACTGGGCCTCCCAAAGTGCTGGG
AT

Microhomology:

```
10T-10U_reverse      --TAATAATAGTAGATCTTAATCAAGTGCTTAGGTGGCGGAGGCCTTGTGTG
LRS                  -ATAATAATAGTAGATTCAAT-CAAGTACTCTATTTGCCAGGCTGGACTCAA
10N-10O              caaattttttgtcaagacaga-gtcttactctatttgccaggctggactca-
                        * *  * **  *           * **    * **   *      *
```

# 10O-10C

-> <-

Expected:
gaaattttaagcacttcttataagatggatttgctggttattaactattt

catatttataggtctaaaacgtcatt|TATGTGTGCAGTTGTGTATGGGGG
AATTTGGGGAGGCTGCGGAGGGTGCATTGTGTGTGCAATTGTGTGTGGCG
GA

LRS:
GACATTTTAAGCACTTCTTATAAGATGGATTTGCTGATTATTAACTATTT
CATATTTATAGGTCTAAAACGTCATT|ATGTGTGCAGTTGTGTATGGGGAA
TTTGGGGAGGCTGCGGAGGGTGCATTGTGTGCAATTGTGTGTGGCGGATT
TT

Microhomology:
```
10O-10P              -atatttataggtctaaaacgtcattgttccactttgtttttgaaagatat
LRS                  -ATATTTATAGGTCTAAAACGTCATTATGTGTGCAGTTGTGTATGGGGAAT
10C-10D_reverse      ATTCTGGGGGGTGAGCAGGGTGCACTATGTGTGCAGTTGTGTATGGGGGA-
                       * *     *     *     ** * *         * * *    *  *
```

# 10C-10S

<-     <-

Expected:
TTCATCTTTGTATTCTGGTAACACAAACAGCATATAACACAGTACCTGCC
TTCTATCAGGAGCTTAAAATATTCAT|atctgcctttatttaattagactt
catatatgaaTTTGCCCTGTTTTCTTTGGAACCACTAAAATAAAGGCCAA
TA

LRS:
TCTTCATATTTTGTATTCTGGTAACACAAACAGCATATAACACAGTACCT
GCCTTCTATCAGGAGCTTAAAATATT|CATCTGCCTTTATTTAATTAGACT
TCATATATGAATTGCCCTGTTTTTCTTTGGAACCACTAAAATAAGAGCCA
AT

Microhomology:
```
10B-10C_reverse      ---TCTATCAGGAGCTTAAAATATTCATTTAAGGAATAATAATGAGTATGAAT
LRS                  CCTTCTATCAGGAGCTTAAAATATTCATCTGCCTTTATTTAATTAGACTT---
10S-10T_reverse      -taccaggaagagaaaatatgtagccatctgcctttatttaattagacttc--
                         *    **       *  **  *** *         **** **  *
```

# 10S-9C

<-   ->

Expected:
TGGAACTGCTCTCTTCCCATGCTAATTTCATTGAACCAAAGGATAGCACA
GTCCTTACAGAATGCAGGCATGTATG|TGAATGATTCTTATTAGCCATGAG
GCAATGCTTGCTCTTGTCAACACATTCTATCCATCCCTTTAGTGAagcag
tc

LRS:
TAAAAAAATGGAACTGCTCTCTTCCCATGCTAATTTCATTAGACAAGGAT
AGCACAGTCCTTACAGAATGCAGGCA|TTGAATGATTCTTATTAGCCATGA
GGCAATGCTTGCTCTTGTCAACACATTCTATCCATCCCTTTAGTGAAGCA

Microhomology:
```
10R-10S_reverse      -------TCCTTACAGAATGCAGGCATGTATGCATTTCTGTTATTTTCTTAAGTCAT
LRS                  AGCACAGTCCTTACAGAATGCAGGCATTGAATGATTCTTATTAGCCATGA-------
9B-9C                --CTCTCTACCTATGCTCCTTTGCACTTGAATGATTCTTATTAGCCATGAGG-----
                            * * **         *   *  *   ***  * ***
```

# 10A-10E

->    <-

Expected:
TTAACACAATATTGGCAAGCTAAATTTAATCAAATATTCTGCCCATTTTA
AAATTCACTTCCTagcctgtccatcc|CCCCTGCCCTCCTCAAGCTTCAGG
TGAAGGCTTTGAGGTGGGAGATATCCCAGATGAAATCCTTCCCTCCAGCC
AT

LRS:
CCAACACAATATTGGCAAGCTAAATTTAATCAAATATTTTGCCCATTTTA
AAATTCACTTCCTAGCCTGTTCATCC|CCCTGCCCTCAAGCTCAGGTGAAG
GTTTTGAGGGTGGAGATATCCCAGATGAAAACTCTTCTCCAGCCATCCCC
AG

Microhomology:

```
10A-10B             -AATTCACTTCCTagcctgtccatccttccttttattttgttgtttgattt
LRS                 AAATTCACTTCCTAGCCTGTTCATCCCCCTGCCCTCAAGCTCAGGTGAAG-
10E-10F_reverse     AAGCAAATTGGAGAGCCTCCCCTTCCCCCTGCCCTCCTC-AAGCTTCAGGT
                     *    * *    *****   * ***  *                * *
```

## 10E-10I

<-   <-

Expected:
agtggggctggaatgtgcatatgcaaaagaatgaacttcaattacacctt
ctcccatatacaaaaagtaactcata|ACCCAAGGTGAGCTCAGCTGAGTG
GGGTGTCGGCCCTGCTCTGGAGAGTGGGTGGGGAGGTGGGCCTGGCTGTG
TT

LRS:
AGTGGGGCTGGAATGTGCATATGCAGAAGAATGAACTTCAATTACACCTT
CTCCCATATACAAAAAGTAACTCATA|CCCAAGGTGAGCTCAGCTGAGTGG
GGTGTCGGCCCTGCTCTGGAGAGTGGTGGGGAGGTGGGCCTGGCTGTGTT
AG

Microhomology:

```
10D-10E_reverse     -tcccatatacaaaaagtaactcatagacctacatgtaaatcctaaaactg
LRS                 -TCCCATATACAAAAAGTAACTCATACCCAAGGTGAGCTCAGCTGAGTGGG
10I-10J_reverse     GACAAACCAGGCCAGGACTACTATGACCCAAGGTGAGCTCAGCTGAGTGG-
                      *  *       *     ***   *  *             ** *
```

## 10I-10W

<-     ->

Expected:
cagaacagtgagccaattaaacatctttttctttatataaattacccagt
ctaaggtatgtctttatagcagtgtg|tggcacattgagcagtatcagaga
atgataagaaataaaactaaagaaagtaagcaggtgccagatGACTAGGG
CC

LRS:
TGCAGAACAGTGAGCCAATTAAACATCTTTTTCTTTTATATAATTACCCA
GTCTAAGGTATGTCTTTATAGCAGTG|TGGCACATTGAGCAGTATCAGAGA
ATGATAAGAAATAAAACTAAAGAAAGTAAGCAGGTGCCAGATGACTAGGG
CC

Microhomology:

```
10H-10I_reverse     --taaggtatgtctttatagcagtgtgagaatggactaatatgcatacaaat
LRS                 TCTAAGGTATGTCTTTATAGCAGTGTGGCACATTGAGCAGTATCAGAGAA-
10V-10W             caaactgtgggtatctcagcatggttggcacattgagcagtatcagagaa--
                       *  **  ** * *      *  **  *        *    ** * **
```

# 10W-10M

->    ->

Expected:

TCTCCAGTCTGCCTTTATACATGTAGAATGGTAATGTAATTTCTAAAGTA
TATACAGAAGCATTCCAGAAATTAAT|gaagtctttttgccactacttttg
tcagctttgtcgaagggcagatggcagtaggtgtgcagccttatAAAAAT
TA

LRS:

TCTCCAGTCTGCCTTTATACATGTAGAATGGTAATGTAATTTCTAAAGTA
TATACAGAAGCATTCCAGAAATTAAT|GAAGTCTTTTTGCCACTACTTTTG
TCAGCTTTGTCGAAGGGCAGATGGCATTAGGTGTGCAACCTTATAAAAAT
TA

Microhomology:

```
10W-10X      --ATACAGAAGCATTCCAGAAATTAATTGCTATTTTAAATAATCATTTTATC
LRS          --ATACAGAAGCATTCCAGAAATTAATGAAGTCTTTTTGCCACTACTTTTGT
10L-10M      tctcccagcaccatttactgaata--ggaagtctttttgccactacttttgt
                  *** * ****     ***          ***     *  * ***
```

# 10M-10Q

->    <-

Expected:

ttagagcaggaatgaaagcaagcaaaatactcttggaagacggctaagtg
ggtgacttgaaagatccaagtgcctt|ttagatcactgagacagaaaatta
acaaagatattcaggacctgaactcagtgctggatcaaatgcacctgata
ga

LRS:

TAGGAGCAGGAATGAAAGCAAGCAAAATACTCTTGGAAGACGGCTAAGTG
GGTGACTTGAAAGATCCAAGTGCCTT|AGATCACTGAGACAGAAAATTAAC
AAAGATATTCAGGACCTGAACTCAGTGCTGGATCAAATGCACCTGATAGA
CA

Microhomology:

```
10M-10N              --gtgacttgaaagatccaagtgccttgttcatcccttcacttgggggtttt--
LRS                  --GTGACTTGAAAGATCCAAGTGCCTTAG--ATCACTGAGACAGAAAATTAACA
10Q-10R_reverse      gaaatttaacaccccactgacagtattag--atcactgagacagaaaattaa--
                               *     *  *  *  **    *** **      *    **
```

# 10Q-14C

<-    ->

Expected:

ACATAAAGATGTACATGTGTATTTGTGTGTCTGCTTATAAAAATAGAAAC
TTTAAAATAAAAAAATAAAACTAGCA|TAAAACGTAGAGGATGGGGAAAAT
GCAGATGCTGTTTTTCCAACTAAAAATGTTTACAAAAGAACAGACTGTCT
GA

LRS:

ATATAAAGATGTGTTTATGTATTTGTGTGTCTGCTTATAAAAATAGAAAC
TTTAAAATAAAAAAATAAAACTAGCA|TAAAACGTAGAGGATGGGGAAAAT
GCAGATGCTGTTTTTCCAACTAAAAATGTTTACAAAAGAACAGACTGTCT

Microhomology:

```
10P-10Q_reverse      -TTAAAATAAAAAAATAAAACTAGCACTCTTGTTTTCttttttttttttaa
```

```
LRS                  TTTAAAATAAAAAAATAAAACTAGCATAAAACGTAGAGGATGGGGAAAAT-
14B-14C              -TGCACACAAAAACAAGATTCCTCTCTAAAACGTAGAGGATGGGGAAAATG
                      *  * * ***** *  *  *            *      *
```

## 14A-10Y

->     ->

Expected:

GGAGCTGTGGAGGGGGCTGTTTCAACGTTGAGAGCAGAAGGCAGGGAGGC
TGTAAGGCAGAAGTAGAGGCAGCTGG|cagctaccagggcaggtagagaac
taccattaggtggcggtagggttaggcgggtctgttctcagactctcctt
gg

LRS:

ACACAGTTCGGGAGCTGTGGAGGGGCTATTTCACTGCGGAGATGAGAACA
GGAGGCTAAAGGGCAGAAGTAGAGG|CAGCTACCAGGGCAGGTAGAGAACT
ACCATTAGGTGGCGGTAGGGTTAGGCGGGTCTGTTCTCAGACTCTCCTTG

Microhomology:

```
14A-14B      -------GTAAGGCAGAAGTAGAGGCAGCTGGTGGCCAGCGTGGTGGCCCCACAGGG
LRS          GGAGGCTAAAGGGCAGAAGTAGAGGCAGCTACCAGGGCAGGTAGAGAACT-------
10X-10Y      tcctaagagtttatgtgtttgtcttcagctaccagggcaggtagagaact-------
                                *     *****    *     ** * *  *
```
