## Supplemental methods for "Full characterization of unresolved structural variation through long-read sequencing and optical genome mapping"


Griet De Clercq[1,2], Lies Vantomme[1], Barbara Dewaele[3], Bert Callewaert[1,2], Olivier Vanakker[1,2], Sandra Janssens[1,2], Bart Loeys[4], Mojca Strazisar[5.6], Wouter De Coster[5,6], Joris Robert Vermeesch[3,7], Annelies Dheedene[2], Björn Menten*[1,2]

1. Department of Biomolecular Medicine, Ghent University, Ghent, Belgium
2. Center for Medical Genetics Ghent, Ghent University Hospital, Ghent, Belgium
3. Center for Human Genetics Leuven, University Hospital Leuven, Leuven, Belgium.
4. Center for Medical Genetics Antwerp, University of Antwerp, Antwerp University Hospital, Antwerp, Belgium.
5. VIB Center for Molecular Neurology, VIB, Antwerp, Belgium
6. Department of Biomedical Sciences, University of Antwerp, Antwerp, Belgium
7. Department of Human Genetics, KU Leuven, Leuven, Belgium.

#### Table of contents


**1 Clinical summaries** **3**

**2 Source code of long-read sequencing analysis** **4**

**3 Interpretation of subway plots** **8**

**4 References** **8**

### 1 Clinical summaries

The clinical summaries have been summarized to be relevant only for the disorders at hand and subsequent conducted LRS and OGM analyses. More detailed clinical information can be requested from the corresponding authors if necessary.

**Individual S1**

A male proband was referred to the clinic for severe ID and dysmorphic facial features. Karyotyping revealed a *de novo* balanced reciprocal translocation between chromosome bands 9q21.2 and 10p15.2 (46,XY,t(9;10)(q21.2;p15.2)) (sup. fig. S1a). This was further confirmed through FISH (sup. fig. S1b). Although the exact breakpoints of the translocation could not be determined, it was considered as likely causal for the underlying phenotype as other tests came back normal.

**Individual S2**

A female proband presented with mild ID, Down-like features, and autism spectrum disorder. Whole exome sequencing analysis indicated a heterozygous deletion of exon 9 of the *MYT1L* (NC_000002.12(NM_001303052.2):c.(152+1_153-1)_(504+1_505-1)) through the ExomeDepth algorithm[1]. Truncating variants in *MYT1L* are described in autosomal dominant intellectual developmental disorder 39 (OMIM #616521), and the deletion was thus characterized as pathogenic.

**Individual S3**

This male individual received a clinical diagnosis of Mowat-Wilson syndrome (OMIM #235730). A balanced reciprocal *de novo* translocation between chromosome bands 2q22 and 21q21 was detected through karyotyping (46,XY,t(2;21)(q22;q21)) (sup. fig. S1c). Mowat-Wilson syndrome is caused by haploinsufficiency due to heterozygous variants in the *ZEB2* gene, which localizes on chromosome band 2q22. The translocation was therefore considered as likely causal for the observed clinical features, although the exact breakpoint location could not be determined. CNV analysis through microarray revealed no further abnormalities.

**Individual S4**

This male individual was seen in the clinic for severe developmental delay, autism spectrum disorder, microcephaly, and dysmorphic facial features. CNV analysis through shallow WGS revealed a *de novo* 2.6 Mb triplication at chromosome bands 3p21.31 to 3p21.1 (sseq[GRCh38]3p21.31p21.1(50100001_52755000)x3 dn) (sup. fig. S2a). This triplication was immediately flanked upstream by a smaller 90 kb *de novo* duplication at chromosome band 3p21.31 (sseq[GRCh38]3p21.31p21.31(50100001_50100000)x2 dn). Other diagnostic tests revealed to further molecular aberrations. Therefore, due to the large size of the triplication and the number of implicated genes this variant was considered as likely causal for the underlying phenotype, although the variant remained classified as a variant of unknown significance.

**Individual S5**

This male was referred to the clinic due to ID and autism spectrum disorder. Karyotyping revealed a *de novo* reciprocal balanced rearrangement between the long arms of chromosomes 2 and 7 (46,XY,t(2;7)(q36;q32)) (sup. fig. S1d). Subsequent microarray analysis further indicated a *de novo* 477 kb deletion in chromosome band 2q24.2 (arr[GRCh37]2q24.2(162105681_162582744)x1 dn) and a *de novo* 186 kb deletion in chromosome band 2q36.3 (arr[GRCh37]2q36.3(229936820_230122602)x1 dn) (sup. fig. S2b,c). Due to the cytogenetic location of the translocation and the 186 kb deletion at chromosome band 2q36, they were believed to be linked to each other through a more complex rearrangement. The 477 kb deletion at location 2q24.2 encompasses the *TBR1* gene, which is involved in intellectual developmental disorder with autism and speech delay (OMIM #606053). This variant was therefore characterized as pathogenic.

##### Individual S6

A woman was referred to the clinic after repeat miscarriages, and failed implantation attempts during in vitro fertilisation treatment. Karyotyping identified a complex rearrangement between chromosomes 9, 10, and 14 (46,XX,ins(9;10)(p22;q11.2q21.2),t(10;14)(q21.2;q32.3)) (sup. fig. S1e). These variants were confirmed with FISH (sup. fig. S1f,g). Shallow WGS revealed six deletions in chromosomes 10 and 14, ranging from 30 to 120 kb (sup. fig. S3; see also table 1 below). The detected complex karyotype was considered causal for the observed clinical features as this can lead to unbalanced gametes and thus reproduction issues. This woman was otherwise phenotypically normal.

**Table 1** Deletions larger than 15 kb found through shallow WGS in individual S6 in the regions implicated by the observed complex karyotype 46,XX,ins(9;10)(p22;q11.2q21.2),t(10;14)(q?;q32.3). Size is listed in kb.

| chr | location | start | end | size | notation |
|---|---|---|---|---|---|
| 10 | 10q11.22q11.22 | 48450001 | 48480000 | 30 | sseq[GRCh38]10q11.22q11.22(48450001_48480000)x1 |
| 10 | 10q11.23q11.23 | 49650001 | 49680000 | 30 | sseq[GRCh38]10q11.23q11.23(49650001_49680000)x1 |
| 10 | 10q21.1q21.1 | 54270001 | 54390000 | 120 | sseq[GRCh38]10q21.1q21.1(54270001_54390000)x1 |
| 10 | 10q21.1q21.1 | 55935001 | 55980000 | 45 | sseq[GRCh38]10q21.1q21.1(55935001_55980000)x1 |
| 10 | 10q21.1q21.1 | 58395001 | 58455000 | 60 | sseq[GRCh38]10q21.1q21.1(58395001_58455000)x1 |
| 14 | 14q32.33q32.33 | 10353001 | 103560000 | 30 | sseq[GRCh38]14q32.33q32.33(103530001_103560000)x1 |

#### 2 Source code of long-read sequencing analysis

##### Guppy v6.3.7 basecaller

```
guppy_basecaller \
   --device cuda:all \
   --input_file_list {fast5.txt} \
   --save_path {save_dir} \
   --recursive \
   --config {config}.cfg \
   --compress_fastq \
   --disable_qscore_filtering \
   --detect_adapter \
   --trim_strategy 'dna' \
   --trim_adapters \
   --gpu_runners_per_device 4 \
   --chunks_per_runner 128 \
   --num_callers 4 \
   --chunk_size 2500
```

with `{fast5.txt}` a text file describing the paths to the FAST5 files to be basecalled (one per line), and `{config}` for samples sequenced with the SQK-LSK109 kit on an R9.4.1 flow cell equal to `dna_r9.4.1_450bps_sup` for MinION runs and `dna_r9.4.1_450bps_sup_prom` for PromethION runs.

For samples sequenced with the SQK-LSK114 kit on an R10.4.1 flow cell only simplex basecalling was performed. The {config} parameter was set to dna_r10.4.1_e8.2_400bps_sup for both MinION and PromethION runs and an additional flag --do_read_splitting was added to split duplex reads into simplex reads.

**Guppy v6.3.7 barcoder**

```
guppy_barcoder \
    --worker_threads {threads} \
    --num_barcoding_threads {threads} \
    --input_path {input_dir} \
    --save_path {save_dir} \
    --barcode_kits "EXP-NBD104" \
    --detect_barcodes
    --records_per_fastq 0 \
    --compress_fastq \
    --fastq_out
```

with {input_dir} the directory containing the FASTQ reads as basecalled by Guppy basecaller.

**Minimap2 v2.24**

```
minimap2 -a -x map-ont --MD -Y -t {threads} --seed 42 {genome.mmi} {
    reads.fastq.gz} | samtools view -@ {threads} -bh - | samtools sort -@
     {threads} -m {memory}GB - > {alignment.sorted.bam}
samtools index -@ {threads} {alignment.sorted.bam}
```

with {genome.mmi} the index to the reference genome as produced by Minimap2 using:

```
minimap2 -t {threads} -x map-ont -d {genome.mmi} {genome.fa}
```

For the {genome.fa} parameter the soft masked hg38 analysis set reference genome without alternative contigs was used (hgdownload.cse.ucsc.edu/goldenPath/hg38/bigZips/analysisSet/).

**Samtools v1.15 merging**

```
samtools merge -@ {threads} -rf {merged.bam} {alignment1.bam} {
    alignment2.bam}
samtools sort -@ {threads} -m {memory}GB {merged.bam} > {merged.sorted.
    bam}
samtools index -@ ${threads} {merged.sorted.bam}
```

**Sniffles2 v2.0.7**

```
sniffles \
    --input {alignment.sorted.bam} \
    --vcf {variants.vcf} \
    --reference {genome.fa} \
    --tandem-repeats {tandem_repeats.bed} \
    --threads {threads} \
    --minsupport 1 \
    --minsvlen 30 \
    --mapq 20 \
    --min-alignment-length 500 \
    --sample-id {sample} \
```

```
    --output-rnames \
    --allow-overwrite
```

with `{genome.fa}` the soft masked hg38 analysis set reference genome without alternative contigs (hgdownload.cse.ucsc.edu/goldenPath/hg38/bigZips/analysisSet/) and `{tandem_repeats.bed}` the associated tandem repeats in `{genome.fa}` as identified through RepeatMasker (https://raw.githubusercontent.com/fritzsedlazeck/Sniffles/master/annotations/human_GRCh38_no_alt_analysis_set.trf.bed).

**Mosdepth v0.3.3**

```
mosdepth -t {threads} {prefix} {alignment.sorted.bam}
```

with `{prefix}` a string describing the sample.

**Samtools v1.15 filtering**

```
samtools view -@ {threads} -bh {alignment.bam} {region_of_interest} |
    samtools sort -@ {threads} -m {memory}GB - > {alignment_filtered.
    sorted.bam}
samtools index -@ {threads} {alignment_filtered.sorted.bam}
```

with `{region_of_interest}` the genomic regions of interest formatted as `chr1:start1-end1 chr2:start2-end2 ... chrN:startN-endN`

**VCFtools v0.1.16**

```
vcftools --vcf {variants.vcf} --stdout --bed {regions.bed} --recode-INFO
    -all --recode > {variants_filtered.vcf}
```

with `{regions.bed}` the bed file listing the relevant genomics regions of interest to filter to.

**WisecondorX v1.2.5**

WisecondorX consists of three separate steps which respectively cover the converting of BAM files of both the test and references samples into an .npz format, the building of the reference set, and the calling of the CNVs.

We first converted the BAM files of each test and reference samples into an .npz format:

```
WisecondorX convert --reference {genome.fa} --normdup {alignment.sorted.
    bam} {alignment.npz}
```

with `{genome.fa}` the soft masked hg38 analysis set reference genome without alternative contigs (hgdownload.cse.ucsc.edu/goldenPath/hg38/bigZips/analysisSet/).

Next, the reference set based upon 36 in-house sequenced LRS samples was built using:

```
WisecondorX newref --binsize 15000 --cpus {threads} {
    reference_alignments/*.npz} reference_set.npz
```

with `{reference_alignments/*.npz}` the directory holding all the to npz converted alignments of the reference samples.

Finally, CNVs were called on each test sample with:

```
WisecondorX predict --bed --plot {test_alignment.npz} reference_set.npz
    {prefix}
```

with `{prefix}` a string describing the test sample.

CNVs in the 'aberrations.bed' file were then filtered to only retain events with a ratio lower than -0.50 or higher than 0.35. This equals respectively to half of $log2(1/2)$ and $log2(3/2)$, which are the ratios for a heterozygous deletion and duplication. Remaining CNVs with one or two of their breakpoints lying within centromeres were then further removed.

In addition, WisecondorX stresses for the use of a reference set where all the samples are exclusively negative controls and subject to the same protocols as the test samples. While this is the case for the majority of the samples within this study, coverage can still differ between samples as this depends on the number of pores available within a flow cell and the sequencing device used. As such, we tested two approaches. In the first one samples were compared to each other without downsampling and thus with differing coverage numbers. In the second approach all reference and test samples were downsampled to the same coverage, i.e. to the sample with the lowest coverage available (sample S1 - 4.77x). While the latter approach yielded far less false positive results, this also resulted in false negative CNV calls (verified by previously performed shallow WGS). As the first approach yields more true positive results, and false positive variants can be filtered out with the technique explained above, we opted to implement the first approach into the study.

**NanoFilt v2.8.0**

```
gunzip -c {reads.fastq.gz} | NanoFilt -q 10 | gzip > {reads_filtered.
    fastq.gz}
```

**Clair3 v1.0.2**

Clair3 was executed through a singularity container available through Docker Hub.

```
singularity exec \
    clair3_v102.sif \
    /opt/bin/run_clair3.sh \
    --ref_fn={genome.fa} \
    --bam_fn={alignment.sorted.bam} \
    --sample_name={sample} \
    --threads={threads} \
    --model_path={model} \
    --platform="ONT"\
    --output={outdir} \
    --min_coverage=3
```

with `{genome.fa}` the soft masked hg38 analysis set reference genome without alternative contigs (`hgdownload.cse.ucsc.edu/goldenPath/hg38/bigZips/analysisSet/`), and `{model}` the `r1041_e82_400bps_sup_g615` model for samples run on R10.4.1 flow cells as downloaded from `https://github.com/nanoporetech/rerio/tree/master/clair3_models`. No phasing was necessary for any sample ran on an R9.4.1 flow cell and as such no SNVs were called in those samples.

**WhatsHap v1.7**

Long reads were phased using:

```
whatshap phase \
    --output {variants_phased.vcf} \
    --reference {genome.fa} \
    --ignore-read-groups \
    {variants.vcf} {alignment.sorted.bam}
```

with {variants.vcf} a vcf-file holding accurate SNVs calls for your interrogated sample, and {genome.fa} the soft masked hg38 analysis set reference genome without alternative contigs (hgdownload.cse.ucsc.edu/goldenPath/hg38/bigZips/analysisSet/).

Reads were haplotagged for further visualization using:

```
whatshap haplotag \
   --output-threads {threads} \
   --output {alignment.phased.bam} \
   --reference {genome.fa} \
   --ignore-read-groups \
   --tag-supplementary \
   {variants_phased.vcf} {alignment.sorted.bam}

samtools sort -@ {threads} -m {memory}GB {alignment.phased.bam} > {
   alignment.phased.sorted.bam}
samtools index -@ {threads} {alignment.phased.sorted.bam}
```

with {genome.fa} the soft masked hg38 analysis set reference genome without alternative contigs (hgdownload.cse.ucsc.edu/goldenPath/hg38/bigZips/analysisSet/). The resulting {alignment.phased.sorted.bam} can be visualized in IGV by right-clicking on its BAM alignment track and selecting *Color Alignments by* $\rightarrow$ *tag* and typing *HP*.

#### 3 Interpretation of subway plots

All variants in this manuscript are visualized as co-called subway plots. Through this manner of visualization you get an immediate overview how the different fragments in a rearrangement connect to each other, what their orientation is, how large the affected regions are and where they are located, how complex the situation is, and which genes are impacted.

In each individual the genome at the region of interest can be divided into different fragments according to the exact breakpoint coordinates of the identified SVs (sup. table S3). In (complex) SVs these fragments are then reshuffled and reorganized, creating a local aberrant haplotype or entire derivative chromosome. In a subway plot the normal chromosome is displayed with its division into fragments. Directly under or above these fragmented normal chromosomes the different fragments are again depicted but now in differently colored blocks according to the aberrant haplotype they belong to (each haplotype is assigned a different color). The orientation and connection of the fragments in the newly created haplotype can then be easily determined by following the same-colored lines that connect the fragment blocks. Deleted fragments are greyed out in the fragmented normal chromosomes. The final aberrant haplotypes are then also visualized along with the deleted or duplicated fragments to offer a quick view of the connection and orientation of the different fragments.

#### 4 References

[1] Plagnol V, Curtis J, Epstein M, Mok KY, Stebbings E, Grigoriadou S, et al. A robust model for read count data in exome sequencing experiments and implications for copy number

variant calling. Bioinformatics. 2012 11;28:2747-54. Available from: https://academic.oup.com/bioinformatics/article/28/21/2747/236565.
